# Design and quality control of large-scale two-sample Mendelian randomisation studies

**DOI:** 10.1101/2021.07.30.21260578

**Authors:** Philip C. Haycock, Maria Carolina Borges, Kimberly Burrows, Rozenn N. Lemaitre, Sean Harrison, Stephen Burgess, Xuling Chang, Jason Westra, Nikhil K. Khankari, Kostas Tsilidis, Tom Gaunt, Gib Hemani, Jie Zheng, Therese Truong, Tracy OMara, Amanda B. Spurdle, Matthew H. Law, Susan L. Slager, Brenda M. Birmann, Fatemeh Saberi Hosnijeh, Daniela Mariosa, ACCC, CCFR-CORECT-GECCO, EPITHYR, InterLymph, ECAC, ILCCO, KIDRISK, PRACTICAL, PanScan, PanC4, Chris I. Amos, Rayjean J. Hung, Wei Zheng, Marc J. Gunter, George Davey Smith, Caroline Relton, Richard M Martin

## Abstract

**Background:** Mendelian randomization studies are susceptible to meta-data errors (e.g. incorrect specification of the effect allele column) and other analytical issues that can introduce substantial bias into analyses. We developed a quality control pipeline for the Fatty Acids in Cancer Mendelian Randomization Collaboration (FAMRC) that can be used to identify and correct for such errors.

**Methods:** We invited cancer GWAS to share summary association statistics with the FAMRC and subjected the collated data to a comprehensive QC pipeline. We identified meta data errors through comparison of study-specific statistics to external reference datasets (the NHGRI-EBI GWAS catalog and 1000 genome super populations) and other analytical issues through comparison of reported to expected genetic effect sizes. Comparisons were based on three sets of genetic variants:

1) GWAS hits for fatty acids, 2) GWAS hits for cancer and 3) a 1000 genomes reference set.

**Results:** We collated summary data from six fatty acid and 49 cancer GWAS. Meta data errors and analytical issues with the potential to introduce substantial bias were identified in seven studies (13%). After resolving analytical issues and excluding unreliable data, we created a dataset of 219,842 genetic associations with 87 cancer types.

**Conclusion:** In this large MR collaboration, 13% of included studies were affected by a substantial meta data error or analytical issue. By increasing the integrity of collated summary data prior to their analysis, our protocol can be used to increase the reliability of post-GWAS analyses. Our pipeline is available to other researchers via the CheckSumStats package (https://github.com/MRCIEU/CheckSumStats).

## Background

Summary data from genome-wide association studies (GWAS) provide a rich resource for two-sample Mendelian randomisation (MR) studies of exposure-disease pathways (see **Box 1** for a general overview of MR). To strengthen causal inference, MR studies evaluate the sensitivity of their results to violations of analytical or instrumental variable assumptions, such as the presence of horizontal pleiotropy, for which an increasingly broad and sophisticated range of methods are available^1–3^. An additional, often overlooked, source of bias in MR studies are errors in the underlying summary or meta data. For example, incorrect specification of the effect allele column may lead to effect estimates that are in the wrong direction^4^. These errors occur because conventions for the inclusion or naming of data fields that avoid ambiguity have not been widely adopted by the GWAS community, increasing the potential for mis-interpretation by data analysts^5^. GWAS summary data can also be obtained from an increasingly diverse range of sources, including online platforms and study-specific websites, but it is not always clear whether such results have been through post-GWAS filtering steps (e.g. with low frequency or poorly imputed SNPs excluded), which increases the potential for unreliable genetic associations. The potential for meta and summary data errors is compounded in relatively complex MR study designs, such as in MR-PheWAS^6–8^ (MR-phenome-wide association study), wide-angled MR^9,10^ and pan-disease MR^11^, in which summary datasets from many different studies, corresponding to many different exposures and/or outcomes, are collated and harmonised into a single analysis.

Within the GWAS field, quality control (QC) procedures have been developed that can detect a wide range of analytical issues and meta data errors, either at the GWAS stage^12^ or at the post-GWAS meta-analysis stage^13^. For example, it is common practice to exclude genetic variants of low genotype or imputation quality or with low minor allele counts, since inclusion of such variants can lead to unstable genetic effect estimates and increase the rate of type I errors^13^. A widely used QC strategy for the identification of meta and summary data errors in GWAS meta-analyses is to compare study-specific statistics to external reference datasets or to results based on theoretical expectations^13^. Some of these QC procedures can also be used in the MR context to identify potential issues with the summary data. For example, effect allele coding errors can be identified by comparing reported allele frequency to allele frequency in a reference population. However, MR studies are subject to a unique set of challenges that often hamper the application of some previously developed QC checks. For example, to reduce the risk of individual re-identification, some consortia do not share allele frequency information with external researchers or replace it with the allele frequency of a reference population. A further hindrance is that metrics of genotype or imputation quality, or of between-study heterogeneity (in the meta-analysis context), are often not made available in GWAS results files. Some QC procedures flag potential issues by comparing study-specific statistics across studies^13^ but under the assumption that all studies employed the same regression models with the same outcomes, covariates and trait transformations, which is unlikely in complex MR study designs. Some studies only make small subsets of GWAS summary data available to researchers, which makes detecting errors harder.

In the present paper we describe a pipeline for the QC of GWAS summary data developed for the Fatty Acids in Cancer Mendelian Randomization Collaboration (FAMRC), a pan-cancer MR study that seeks to evaluate the causal relevance of fatty acids for risk for most major cancers. The basic principle of our QC approach is to identify meta data errors through comparison of study-specific statistics to external reference datasets (e.g. the NHGRI-EBI GWAS catalog and 1000 genome super populations) and to identify potential analytical issues or summary data errors through comparison of reported to expected genetic effect sizes. Using the pipeline, we collated, harmonised and quality controlled 219,851 genetic associations with 87 cancer types in 160 datasets and 48 separate studies, consortia or biobanks. The size and complexity of the FAMRC makes it an ideal collaboration in which to develop and evaluate QC processes for the detection of errors that can introduce biases into downstream MR analyses.

### Box 1

General overview of Mendelian randomisation studies

The main aim of Mendelian randomisation is to assess the potentially causal nature and direction of associations between exposure and outcome traits. Rather than study the exposure-outcome relationship directly, i.e. using phenotypically measured levels of the exposure, MR uses genetic polymorphisms as instruments or proxies for the exposure. If the genetic instrument for the exposure is associated with the outcome of interest, this can be taken as evidence for a causal effect of the exposure on the outcome, so long as instrumental variable (IV) assumptions are met: i) the instrument is associated with the exposure; ii) the instrument is not associated with confounders of the exposure-outcome association; and iii) the instrument is associated with the outcome exclusively through its effect on the exposure. Although violations of assumptions can be introduced by genomic confounding or horizontal pleiotropy, an increasingly sophisticated range of sensitivity analyses are available that can be used to model the impact of such bias on MR findings.

In the two-sample approach to MR, genetic summary data for the exposure and the outcome are obtained from separate studies. This greatly increases the scope of MR, as it means the method can be applied to any disease case-control collection regardless of whether the exposure has been directly measured or not. The success of genome-wide association studies has greatly increased the number of traits with available genetic association or summary data. In principle, any heritable trait with summary genetic association data can be used to define an exposure or an outcome in a two-sample MR study and thus the scope for what counts as an exposure or an outcome is very broad. Exposure and outcome traits can vary from relatively simple molecular traits, such as expression or protein traits, to highly complex traits, such as human behaviours and disease outcomes.

### Key Messages

- Meta-data errors (e.g. incorrect specification of the effect allele column) and other analytical issues can introduce substantial bias into Mendelian randomization studies but have received relatively little attention in comparison to other sources of bias, such as violations of instrument variable assumptions.
- We found that 13% of the studies in the Fatty Acids in Cancer Mendelian Randomization Collaboration (FAMRC) were subject to meta data errors or analytical issues with the potential to introduce substantial bias into MR analyses (e.g. inferences of causal effect in the wrong direction or bias to the null).
- Previously developed guidelines for conducting MR studies provided insufficient safeguards against such errors.
- We developed additional guidelines and the CheckSumStats R package (https://github.com/MRCIEU/CheckSumStats) that can reliably identify and correct meta-data errors and other analytical issues at the study design stage

## Methods

The FAMRC had four key design components: 1) fatty acid instrument selection strategy; 2) cancer outcome selection strategy; 3) cancer data preparation and harmonisation; and 4) identification of summary and meta data errors and other analytical issues (**Figure 1**).

**Figure 1.**
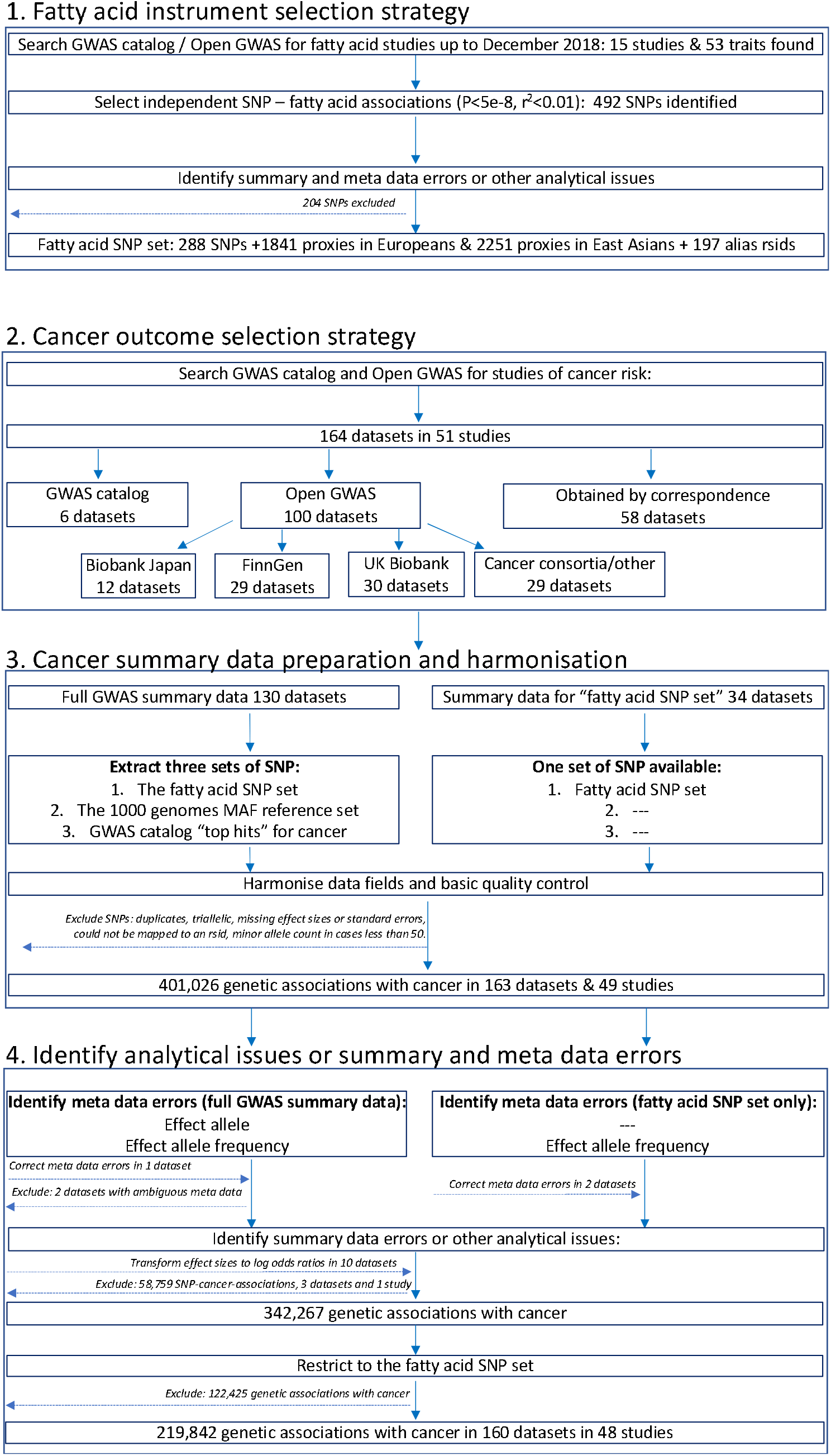
Study design flow chart

### Fatty acid instrument selection strategy

We searched for GWAS of fatty acids published up to December 2018 by searching the NHGRI-EBI GWAS catalog^14^ (https://www.ebi.ac.uk/gwas/) and OpenGWAS^15^ (https://gwas.mrcieu.ac.uk/), using fatty acid related search terms, including: fat, acid, fatty acid, DHA, omega, monounsaturated, mono-unsaturated, polyunsaturated, saturated, omega 3 and omega 6.Fifteen studies were identified by this strategy. When full summary association statistics were available, independent genetic associations with P<5e-8 were identified through linkage disequilibrium (LD) clumping (r^2^ threshold set to 0.01), with LD reference panels based on either the European or East Asian 1000 genome superpopulations (clumping was performed using the ieugwasr package^16^). We also selected all SNP associations reported in the GWAS catalog, with no specified P value threshold. We further identified SNP proxies, defined as SNPs having an r^2^ greater than, or equal to, 0.8 with any one of the fatty acid SNPs in European or East Asian 1000 genomes reference data. We also searched for alias rsids in dbSNP and 1000 genomes reference data, to make allowance for different rsids across different genome builds for the same SNP. We refer to the genetic associations for fatty acids, their r^2^ proxies and alias rsids as the “fatty acid SNP set” (**Figure 1**). To identify meta data errors, summary data errors or other analytical issues, we developed and applied a QC pipeline to the fatty acid summary datasets (described below).

### Cancer outcome selection strategy

We searched for studies of cancer in the GWAS catalog^14^ up to 1^st^ November 2018. Search terms included: cancer, carcinoma, neoplasm, neoplastic, tumor, tumour, adenocarcinoma, glioblastoma, leukemia, lymphoma, melanoma, meningioma, mesothelioma, myeloma, neuroblastoma and sarcoma. When multiple studies of the same cancer outcome were identified, we prioritised the larger study. When not already available via Open GWAS^15^ (https://gwas.mrcieu.ac.uk/) or the GWAS catalog, we invited the identified studies to share summary data for all SNPs in their GWAS analysis (defined as “full GWAS summary data”). If studies were unable to share full summary data, they were invited to share genetic association results for the “fatty acid SNP set”. We further downloaded summary association statistics for cancers from Biobank Japan^17–19^ (http://jenger.riken.jp/en/), FinnGen (data freeze 1 [January 14 2020]) (https://www.finngen.fi/fi) and UK Biobank^20,21^, using the Open GWAS platform^15^ and ieugwasr package^16^. We prioritised studies of cancer incidence and excluded studies of cancer survival, mortality or progression-related phenotypes.

For datasets obtained via correspondence, studies were invited to share summary data up until December 2019, after which data collection was closed. Example data sharing instructions can be found in the supplementary materials. For each SNP, we asked studies to provide a minimum of: effect estimates (log odds ratios and standard errors), the effect allele, non-effect allele and effect allele frequency. We also asked studies to provide metrics of SNP genotype quality, such as P values for Hardy–Weinberg equilibrium and metrics of imputation quality, such as info scores. When the GWAS was a meta-analysis of multiple independent studies, we additionally requested P values for between study heterogeneity.

### Cancer data preparation and harmonisation

For each cancer with full summary data, we extracted the following 3 sets of SNPs:

1. The fatty acid SNP set
2. The 1000 genomes reference set
3. The GWAS catalog “top hits” for cancer

When full summary data were not provided, QC analyses were restricted to the ‘fatty acid SNP set’. We next formatted the cancer summary datasets to have similar tabular formats (e.g. where results were distributed across multiple files we merged these together) and to have consistently named data fields. SNPs without rsids were mapped to an rsid using the reported chromosome and base pair position. We excluded duplicate and triallelic SNPs as well as SNPs with missing effect sizes and standard errors or with a minor allele count less than 50 in either cases or controls. If standard errors were not reported, we attempted to infer this from confidence intervals or P values before excluding the SNP. We asked each study to confirm the identity of the effect allele and effect allele frequency columns in their datasets, unless this was unambiguously specified in the meta data or associated readme file. We manually mapped the cancer name for each dataset to the experimental factor ontology (EFO)^22^.

### Quality control pipeline to identify analytical issues or summary and meta data errors

To identify meta data errors, summary data errors or other analytical issues, we developed a QC pipeline based on the R programming language and associated packages^16,23–33^. We used the pipeline to: 1) confirm the identity of the effect allele column; 2) to confirm the identity of the effect allele frequency column; and 3) to identify analytical issues or potential errors in the summary data (e.g. an unusual number of GWAS hits or unusual distributions in effect sizes). All the functions and tests of the QC pipeline are available to other researchers via the CheckSumStats package (https://github.com/MRCIEU/CheckSumStats).

#### Instrument-specific quality control

To identify potential analytical issues or errors in the genetic instruments for fatty acids, we compared genetic association results identified through LD clumping (r^2^=0.01 and kb=10,000) to associations in the GWAS catalog. We set the significance threshold for LD clumping to the threshold reported in the fatty acid GWAS: 5-e8 in CHARGE^34^, SCHS^35^ and NHAPC/MESA^36^, 1e-8 in the Framingham study^37^, 2.3e-9 in Kettunen et al^38^ and 1.03e-10 in the TwinsUK/KORA study^39^ and used the 1000 genomes European superpopulation as an LD reference panel (except for the SCHS, for which the East Asian superpopulation was used). We searched the GWAS catalog for the lead SNP, identified by our clumping procedure, as well as SNPs within 200,000 base of the lead SNP (associations were retrieved from the GWAS catalog via the gwasrapidd package^33^). Datasets were flagged for further investigation if a substantial number of the lead SNPs were absent from the GWAS catalog. We additionally searched for meta and summary data errors in the fatty acid GWAS results through comparisons of effect alleles and allele frequency with external reference datasets and by comparing reported to expected effect sizes (described below).

#### Confirming the effect allele column

To identify incorrect specification of the effect allele column, we compared summary association statistics in the test dataset (either a fatty acids or cancer dataset) to summary association statistics in the NHGRI-EBI GWAS catalog^14^. The latter is a manually curated database of 251,401 genetic associations from 4961 publications (as of April 2021) and includes information on effect alleles, effect sizes and EFOs. The genetic associations in the manually curated database typically correspond to the statistically significant findings (“top hits”) from published studies (often defined as P<5e-8). In recent years, the GWAS catalog has started to host full GWAS summary statistics. However, for this QC step, we are referring exclusively to the manually curated database of published “top hits”.

In the first step, we searched the GWAS catalog for SNPs associated with the EFO or reported trait of the test dataset. Second, for each SNP associated with the EFO term, we extracted from the GWAS catalog the effect size, standard error, effect allele, effect allele frequency and study ancestry (genetic associations were retrieved via the gwasrapidd package^33^). SNPs missing any of this information, or that were palindromic, were removed. Third, genetic associations for these SNPs were then extracted from the test dataset. Fourth, the effect sizes and effect allele frequencies from the GWAS catalog and test dataset were harmonised to reflect the same effect allele and compared in scatter plots (constructed using the ggplot package^23^). Comparisons were restricted to populations of European or East Asian ancestry.

We then inspected the scatter plots for conflicting directions of association. For example, we declared a conflicting direction of association if the effect allele was associated with higher cancer risk in the GWAS catalog but was associated with lower risk in the test dataset. The level of conflict was further labelled as ‘high’ if the P value for the association was <0.0001 in both the GWAS catalog and the test dataset, and as ‘moderate’ if not, so as to make allowance for chance deviations in effect direction in small studies. If the test dataset and the data in the GWAS catalog corresponded to the same publication, the conflict level was labelled as ‘high’ regardless of the P value strength. For comparisons of allele frequency, we declared a conflict if effect allele frequency was not greater (or less) than 0.5 in both datasets. The level of conflict was further labelled as high if minor allele frequency was ≤0.4 in both datasets, and as moderate if not. The latter step makes allowance for chance deviations in allele frequencies for SNPs with minor allele frequencies close to 0.5. Conflicts were also labelled as high if allele frequency differed by more than 10 points between the test and reference datasets. When interpreting the scatter plots, it is important to take into account the total number of SNPs in the comparison as well as the ancestry of the test and reference datasets. Conflicting associations are more likely to reflect true effect allele coding issues when the conflict is systematic across a large number of SNPs and when the ancestry of the datasets being compared is the same. When a substantial proportion of SNPs displayed effect or allele frequency conflicts, we flagged the test dataset as containing a potential effect allele meta data error.

#### Confirming the effect allele frequency column

To confirm the effect allele frequency column, we compare the test datasets to two types of reference datasets: the 1000 genomes project^40^ and the exposure study. In the case of the present analysis, we used the Cohorts for Heart and Aging Research in Genomic Epidemiology Consortium (CHARGE) and the Singapore Chinese Health Study (SCHS) as representative of exposure (i.e. fatty acid) studies in Europeans and East Asians, respectively. For comparisons with the 1000 genomes project, we created a reference dataset of 2297 SNPs that have the same minor allele across the African, European, East Asian, American, South Asian and Global super populations and that also have a minor allele frequency between 0.1 and 0.35 (this dataset is available to other researchers in the CheckSumStats R package). We refer to the 2297 SNPs as the “1000 genomes reference set”. For comparisons with CHARGE and the SCHS, we created a reference dataset corresponding to the fatty acid SNP set described above (see fatty acid instrument selection strategy). We then compare minor allele frequencies between the test dataset and the reference dataset. The comparison involves the following steps. First, we merge the test and reference dataset. Second, we recode the reported effect allele and reported effect allele frequency in the test dataset to reflect the minor allele in the reference dataset. Third, we compare minor allele frequencies between the datasets in scatter plots^23^ and inspect the plots for conflicting patterns. A conflict is declared for individual SNPs if their allele frequency is greater than 0.5 in the test dataset. If the frequency is also greater than, or equal to, 0.58, the conflict level is upgraded to ‘high’ (to make allowance for chance deviations). Conflicts are also labelled as high if allele frequency differs by more than 10 points between the test and reference datasets. If an inverse correlation is observed across the vast majority of SNPs this indicates that the conflict is systematic and that the reported effect allele frequency actually corresponds to the non-effect allele. When there is a conflict for approximately half the SNPs, this implies that the reported effect allele frequency column actually corresponds to the minor allele and that the minor allele is not consistently the effect allele. In the latter situation, the scatter plot will show two separate groups of SNPs, one with a positive correlation, and the other with an inverse correlation, in allele frequency between the datasets. The strength and linearity of the correlation in allele frequency between the test and reference datasets also provides information on the ancestral background of the participants used to generate the test dataset. An advantage of using our “1000 genomes reference set” is that incorrect specification of effect allele frequency can be identified without knowledge of the ancestral background of the test dataset.

#### Identifying other analytical issues and summary data errors

To identify potential analytical issues or summary data errors, we compare the expected and reported effect sizes. For continuous exposures, such as fatty acid levels, we generate expected betas using the formula:

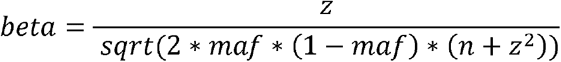

where maf is the minor allele frequency, n is the sample size and Z is the ratio of the effect size to its standard error. The predicted effect size from this transformation can be interpreted as the standard deviation change of the exposure per copy of the effect allele (assuming that z itself was generated in an additive genetic model). When the expected effect size is a log odds ratio, e.g. for cancer status analysed in a logistic regression model, we generate the expected log odds ratio for each SNP using a simulation method that takes into account the SNP’s Z score, minor allele frequency and the number of cases and controls^41^ and assumes an additive genetic model. More details of the method can be found in the supplementary materials.

We then regress the expected on the reported effect size and interpret a slope very different from 1 (which we define as either >1.20 or <0.8) as evidence for potential summary data errors or analytical issues. We also assess the overall shape of the relationship between the expected and reported effect sizes in scatter plots, with the expectation of linearity. Deviations of the slope from one or non-linear patterns could reflect:

1. Errors in the reported effect sizes, sample sizes or allele frequencies.
2. Effect size scale conflicts (e.g. reported effect sizes have not been standardised [for continuous traits] or effect sizes have not been generated in a logistic regression model [for cancer outcomes])
3. The impact of covariate adjustment in the regression model
4. Deviations from Hardy Weinberg Equilibrium (HWE)

If we found that the summary association statistics for cancer were generated in a linear model (e.g. BOLT-LMM^42^), we transformed the effect size to a log odds ratio scale using the following formula:

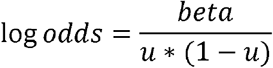

where u is the case prevalence in UK Biobank. The standard error for the log odds ratio can be obtained with the same transformation.

To see whether discrepancies between the reported and expected effect sizes were related to metrics of genotype or imputation quality, we compared discrepancies to reported info or r^2^ imputation scores, P values for Hardy Weinberg Equilibrium and, in the case of meta-analyses, P values for between study heterogeneity and the number of studies. Potential errors in reported effect sizes were also identified by comparing Z_b_ scores (inferred from the reported effect size and standard error) to Z_p_ scores (inferred from the reported P value) in scatter plots (also known as P-Z plots^13^).

## Results

### Fatty acid datasets

We identified 15 GWAS analyses of 53 fatty acid characteristics. Nine of the 15 GWAS analyses were conducted by, or overlapped with, the CHARGE consortium. The median sample size per analysis was 7,811 (min=284; max=17,267), including analyses of 10 monounsaturated fatty acids (MUFAs), 15 saturated fatty acids, 6 omega 3 PUFAs, 11 omega six PUFAs and 11 other characteristics (**Supplementary table 1**). The 15 GWAS analyses corresponded to seven independent studies or consortia and 15 separate publications^35–39,43–52^. An interaction study was the only fatty acid GWAS excluded^53^. We subsequently invited the identified studies to share full summary data with the FAMRC (except for Shin et al^39^ and Kettunen et al^54^, which were already available via Open GWAS). The vast majority of the GWAS analyses were conducted in European ancestry populations (11/15), two were conducted in populations of East Asian ancestry, one in a population of South Asian ancestry and one in a trans-ethnic GWAS of Europeans and East Asians. We collated full summary data from 13 of 15 publications, corresponding to six independent consortia or cohort studies: CHARGE^34,43–45,48,49,51,52^, SCHS^35^, the Framingham study (FHS)^37^, the TwinsUK/KORA study^39^, the NHAPC/MESA-CHI study^36,48^ and Kettunen et al^38^.

To identify meta and summary data errors, we applied a custom QC pipeline to the CHARGE, FHS, SCHS, TwinsUK/KORA, NHAPC/MESA-CHI and Kettunen studies **(Figure 2 and Supplementary figures 1-5**). No allele frequency or effect allele conflicts were observed, indicating that the reported effect allele and effect allele frequency columns were correctly indicated. A strong and positive linear relationship between the expected and reported effect sizes was also observed in the FHS, TwinsUK/KORA, NHAPC/MESA-CHI and Kettunen et al studies, with slopes close to 1, suggesting the absence of major analytical issues in these studies.

**Figure 2.**
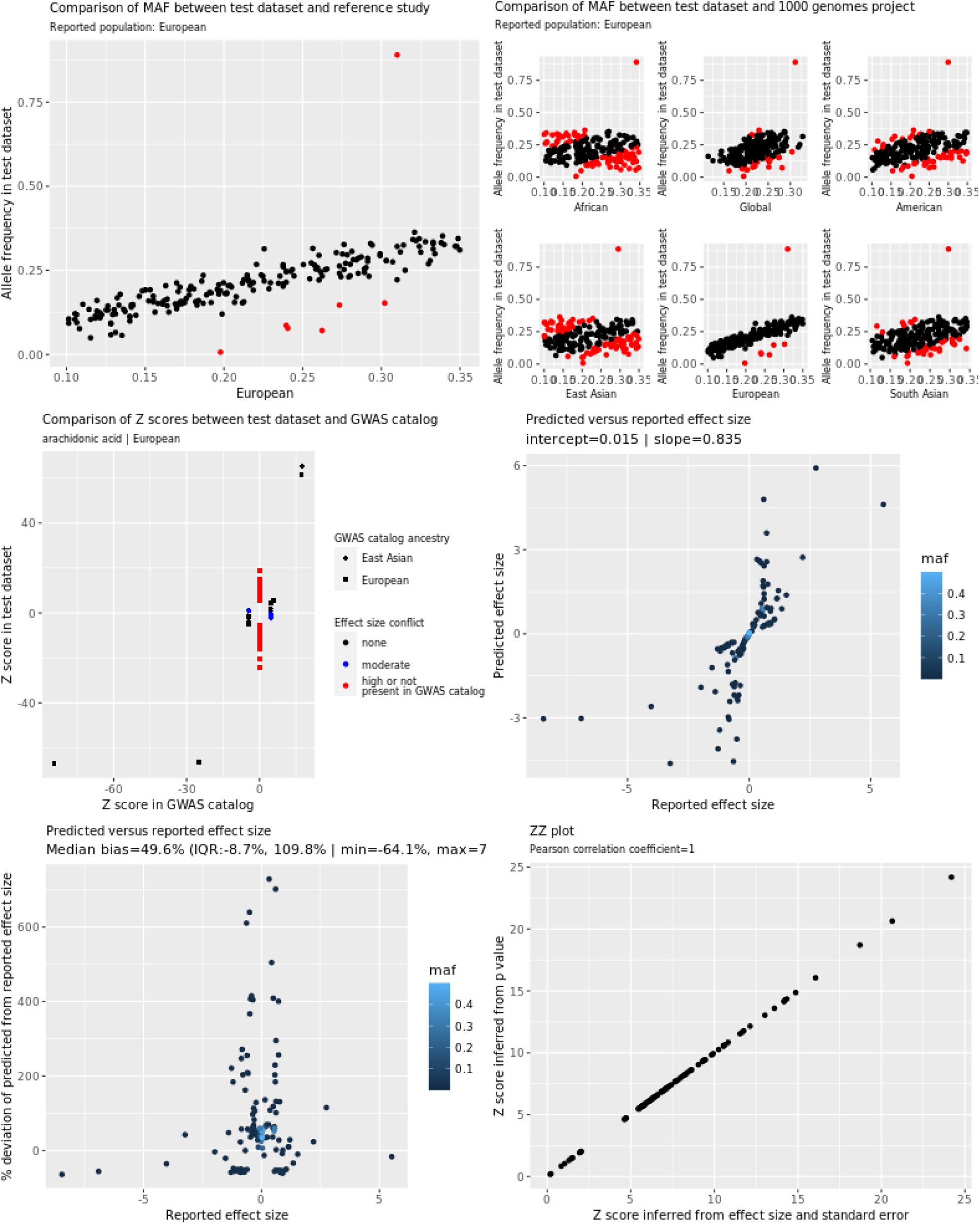
Quality control report for genetic summary data from a genome-wide association of arachidonic acid in CHARGE In the allele frequency subplots, each red data point corresponds to SNPs with high allele frequency conflicts, due to having an allele frequency that is greater than 0.58 (when it is expected to be less than 0.5) or to deviation from the reference allele frequency by more than 10 points

The expected and reported effect sizes for selected fatty acids were not, however, well correlated in the CHARGE (**Figure 2**) and SCHS studies (**Supplementary figure 4)**. We also identified 109 independent GWAS hits for arachidonic acid in CHARGE after LD clumping, of which only four were also reported in the GWAS catalog, compatible with the presence of a large number of false positives. After corresponding with the data provider, we were able to confirm that post-GWAS filtering of low quality variants (defined as SNPs with minor allele frequency <5%, imputation r^2^ < 0.5, or as SNPs that were present in only one study^34^) had not been performed on the dataset posted to the CHARGE website. After excluding these SNPs, following recommendations of the data provider^34^, we observed a strong linear relationship between the reported and expected effect sizes and a slope of 1.02 (**Supplementary figure 6**). The 109 independent GWAS hits also decreased to seven in the cleaned dataset, all of which mapped to the FADS genomic region on chromosome 11 or PDXDC1 on chromosome 16, established GWAS hits for fatty acids (and therefore unlikely to be false positives). In the SCHS, the relationship between the expected and reported effect sizes was skewed by a single outlier SNP (**Supplementary figure 4**). Further investigation revealed that the outlier was due to incorrect specification of the sample size for this SNP.

We also identified two independent GWAS hits for selected fatty acids in the NHAPC/MESA-CHI and SCHS studies that were not present in the GWAS catalog or in the associated publications. We subsequently confirmed that post-GWAS filtering steps for low quality variants had not been applied to the GWAS results files for the NHAPC/MESA-CHI study and that the identified GWAS hit had failed the reported quality control checks (we therefore excluded this variant). In the SCHS, correspondence with the data provider indicated that a file sharing error had occurred and we therefore obtained a new set of GWAS results files (in which conflicts with the GWAS catalog were no longer observed). Conflicts with the GWAS catalog were not observed for the FHS, TwinsUK/KORA and Kettunen et al studies. The false positive GWAS hits identified in the CHARGE and NHAPC/MESA-CHI studies only apply to the results files shared with the FAMRC and do not apply to the reported GWAS findings^34–36,43–45,48,49,51,52^.

After applying the SNP selection strategy and resolving the analytical issues flagged by the QC pipeline, we identified 288 SNPs associated with 53 fatty acid traits (median 6 per trait). We identified a further 1841 SNP proxies using the 1000 genomes European super population, 2251 SNP proxies in the 1000 genomes East Asian super population and 197 alias rsids in dbSNP and 1000 genomes reference data. The total number of SNPs associated with fatty acids, their r^2^ proxies and alias rsids, was 2326 for European studies and 2596 in East Asians (excluding duplicate SNPs that overlapped amongst the fatty acid and proxy sets) (**Supplementary tables 2-3**). We henceforth refer to these SNPs as the “fatty acid SNP set”.

### Cancer datasets

As of January 2020, we had collated 166 summary genetic datasets from 51 cancer studies^17,55–103^. Two datasets did not report information on the effect allele and effect allele frequency and were therefore excluded^97,98^. The 164 retained datasets included 58 obtained via correspondence with study authors, 6 from the GWAS catalog and 100 from the Open GWAS project^104^. Of the 100 Open GWAS datasets, 12 were from Biobank Japan, 29 were from FinnGen, 30 were from UK Biobank, 26 were from GWAS meta-analysis consortia and 3 were from other studies. Further details of the cancer studies can be found in **Table 1 and Supplementary table 5**. Effect allele frequency was available in 155/164 datasets, metrics of imputation quality (r^2^ or info scores) were available in 53/164 datasets and P values for deviations from Hardy Weinberg Equilibrium were available in 6/164 datasets. Of 70 datasets derived from GWAS meta-analyses of multiple studies, P values for between-study heterogeneity were available in 18 and number of studies per SNP was available in 16.

**Table 1.**
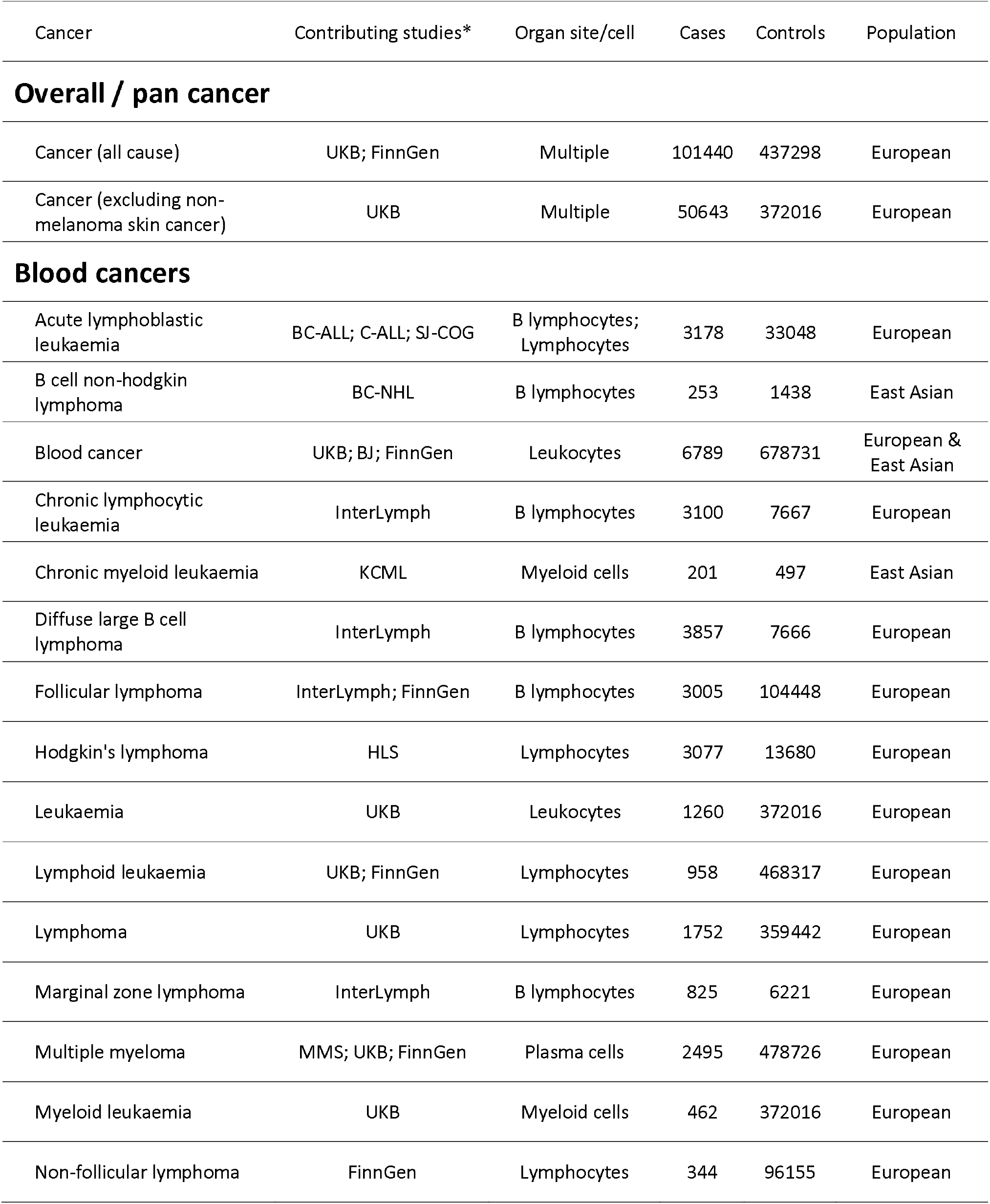
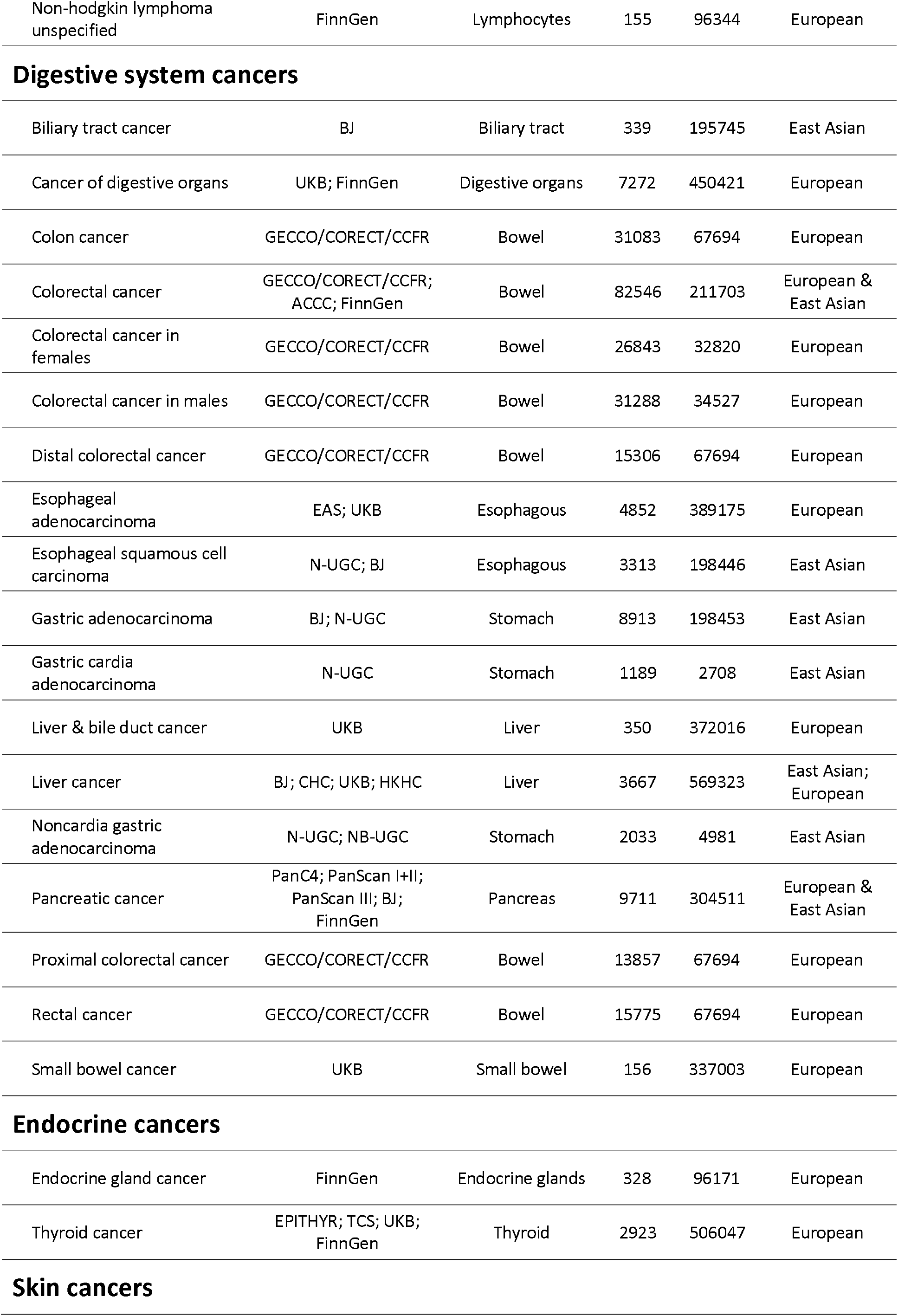
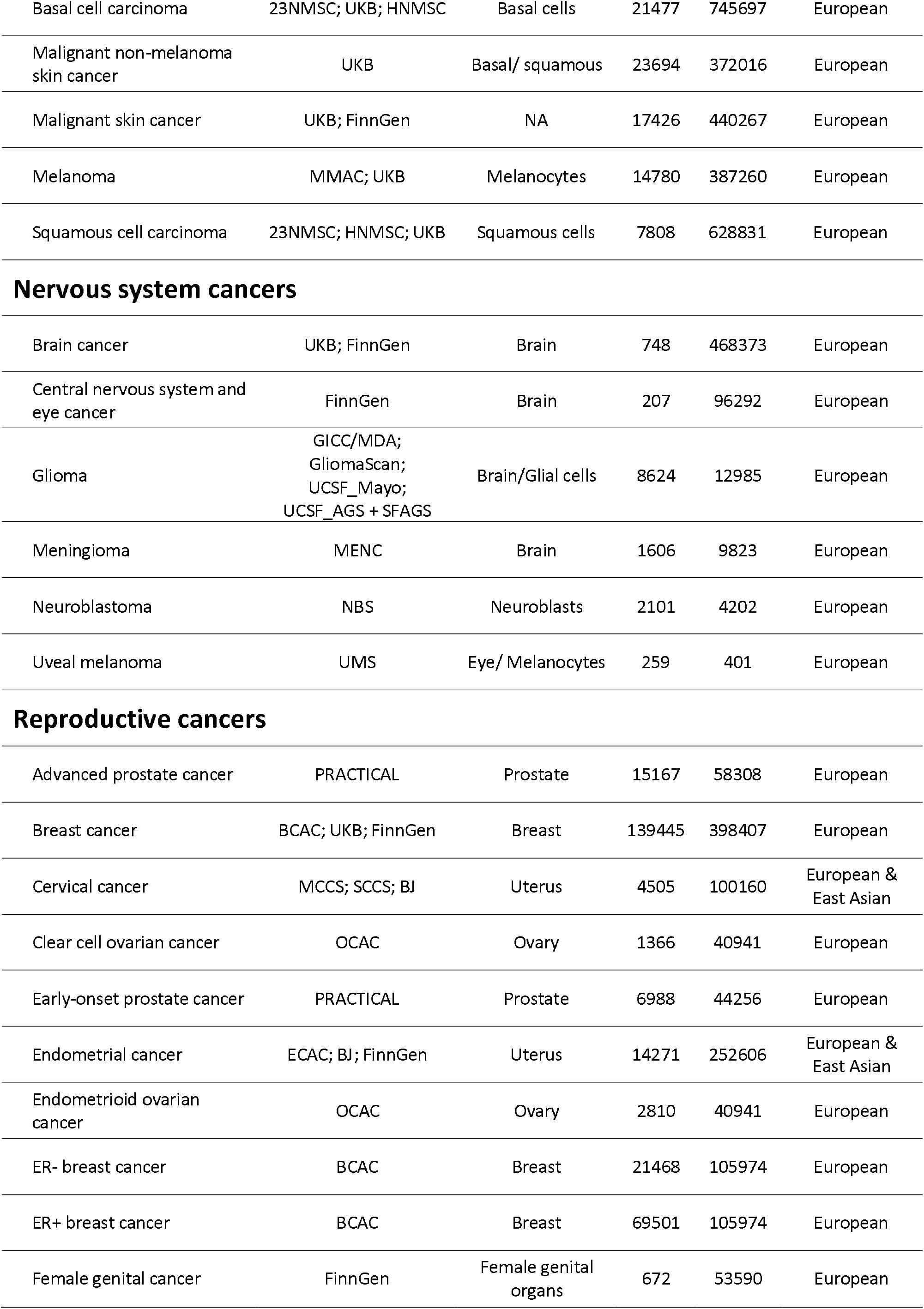
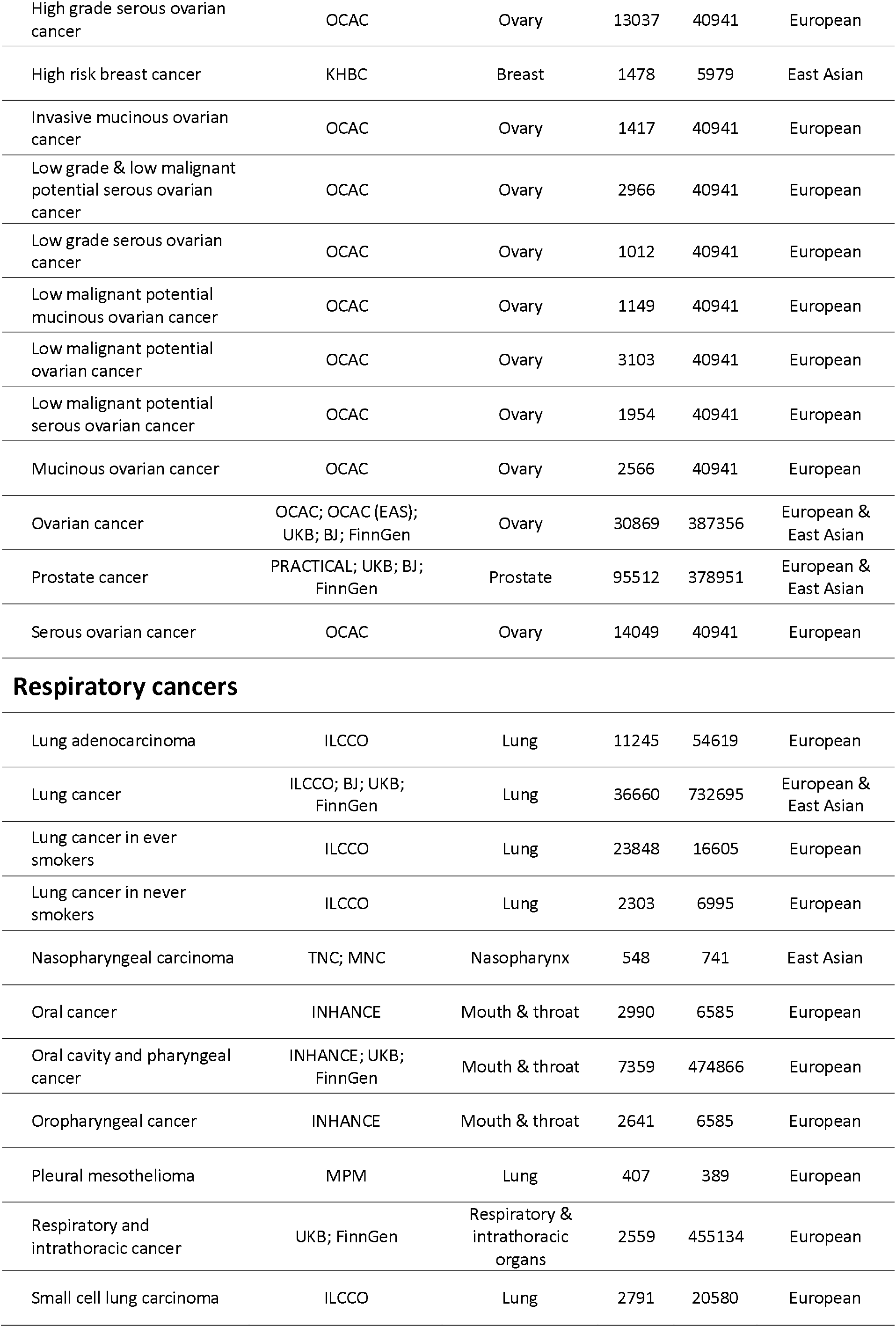
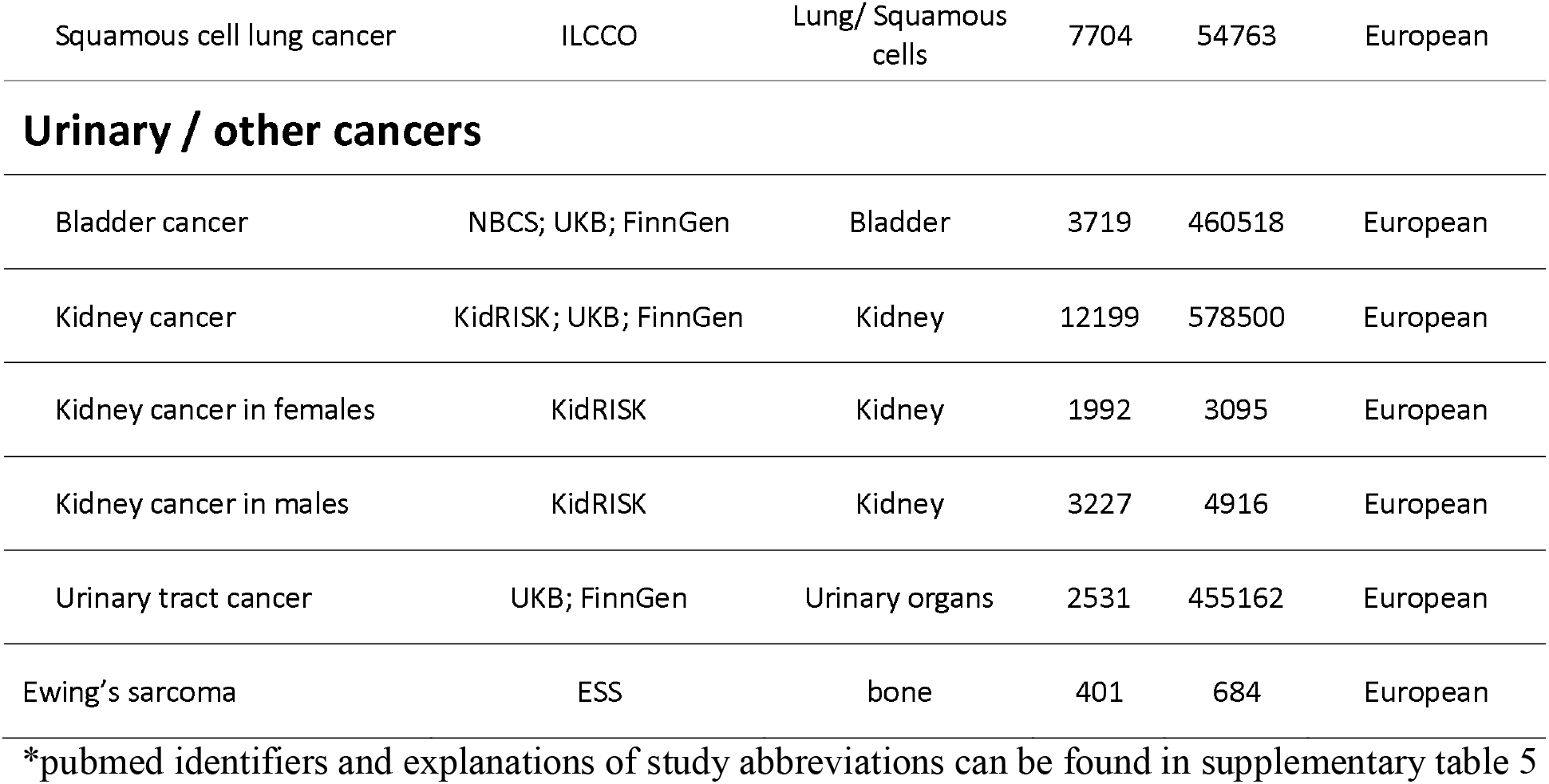
Cancer studies in the Fatty Acids in Cancer Mendelian Randomisation Collaboration

We extracted three sets of genetic associations from each dataset for which full GWAS results were available (130 datasets in total): 1) the fatty acid SNP set, 2) the 1000 genomes reference set and 3) known cancer hits in the GWAS catalog. For 34 datasets, only a subset of GWAS results, corresponding to the fatty acid SNP set, was available. We excluded duplicate and triallelic SNPs, SNPs with missing effect sizes or standard errors, SNPs that could not be mapped to an rsid and SNPs with a minor allele count in cases less than 50. After these exclusions, there were 401,026 genetic associations with cancer across 163 datasets in 49 studies. Of these, 223,970 genetic associations corresponded to the fatty acid SNP set, 93,121 corresponded to the 1000 genomes reference set and 24,860 corresponded to known cancer associations in the GWAS catalog. Three studies providing genetic associations for the fatty acid SNP set provided an additional 40,582 genetic associations for SNPs within 500kb of a fatty acid index SNP (ACCC [ID3], UCSF_AGS + SFAGS [ID133] and UCSF_Mayo [ID134]).

## Results of quality control pipeline applied to the cancer datasets

At least one issue was identified by the QC pipeline in 41 (25%) of 163 total cancer datasets, or in 21 (42.9%) of 49 studies **(Supplementary table 4)**. These included serious meta data errors (defined as incorrect labelling of effect allele or effect allele frequency columns) in 5 (10.2%) of 49 studies. In three datasets, alleles associated with higher cancer risk in the GWAS catalog were associated with lower risk in the test dataset for a substantial proportion of SNPs (**Figures 3-5**), suggesting that the effect allele column actually refers to the non-effect allele. This was clearest for GliomaScan (ID 967) where 19/21 SNPs were discordant with the GWAS catalog but was less clear for the NBS (ID 106) and BC-NHL (ID 5) datasets. In the BC-NHL, although all SNPs were discordant to the GWAS catalog, the number available for comparison was small and Z scores were not highly significant by GWAS standards (Z ≤ 2.5). Therefore, we could not rule out chance deviations from the GWAS catalog for this dataset. In addition, the ancestry of the BC-NHL (East Asian) was different to the ancestry of the reference dataset (European). Therefore, the observed conflict for the BC-NHL dataset could also reflect differences in LD between populations. In the NBS (ID 106), equal numbers of SNPs were highly discordant and highly concordant to the GWAS catalog. Due to the ambiguity of the effect allele we decided to drop the BC-NHL (ID 5) and NBS (ID 106) datasets. Reported effect alleles were compatible with reported cancer hits in the GWAS catalog for other cancer datasets (**Supplementary figure 7)**.

**Figure 3.**
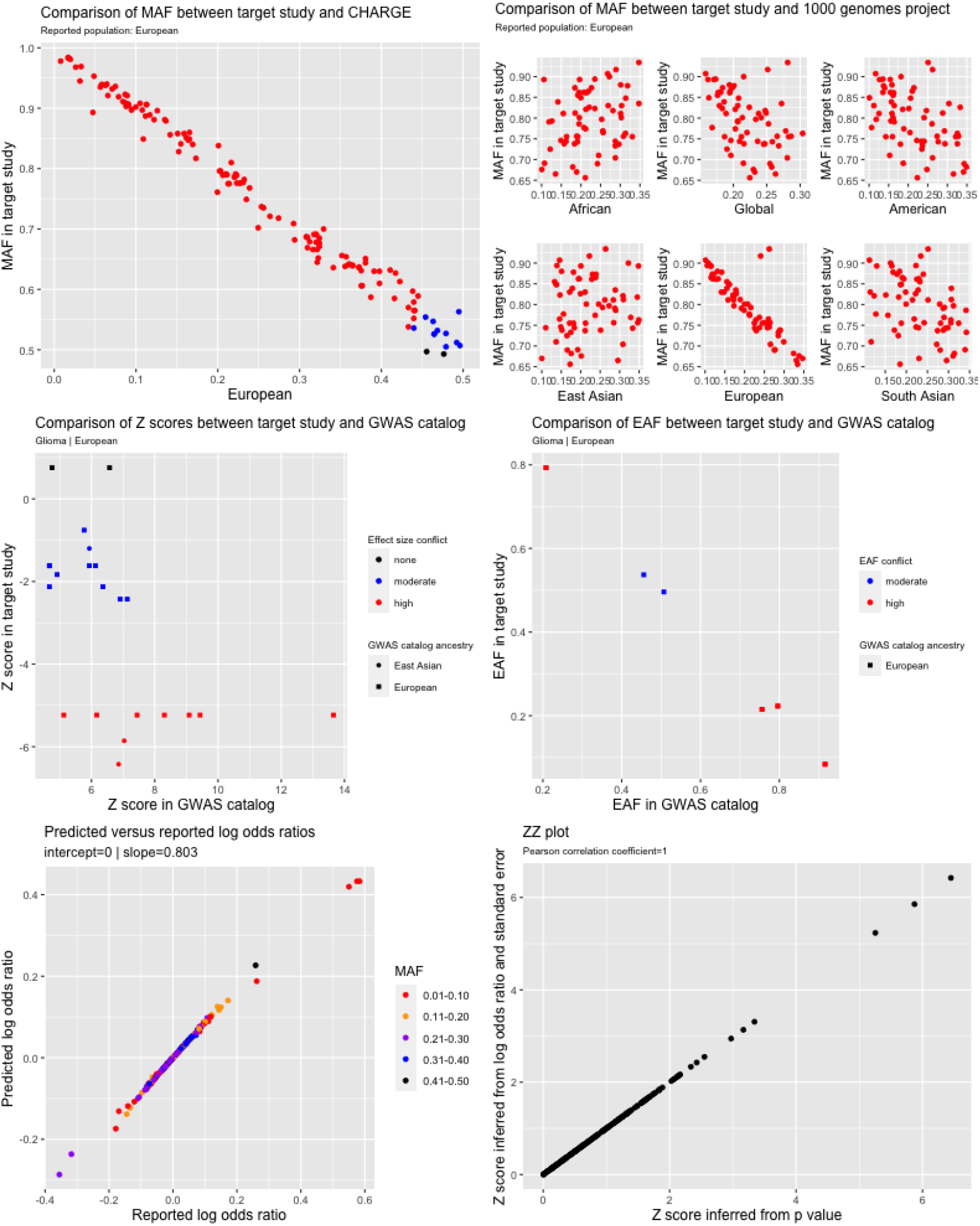
Quality control report for genetic summary data from a genome-wide association of glioma in the GliomaScan dataset (ID 967) In the allele frequency subplots, each red data point corresponds to SNPs with high allele frequency conflicts, due to having an allele frequency that is greater than 0.58 (when it is expected to be less than 0.5) or to deviation from the reference allele frequency by more than 10 points

**Figure 4.**
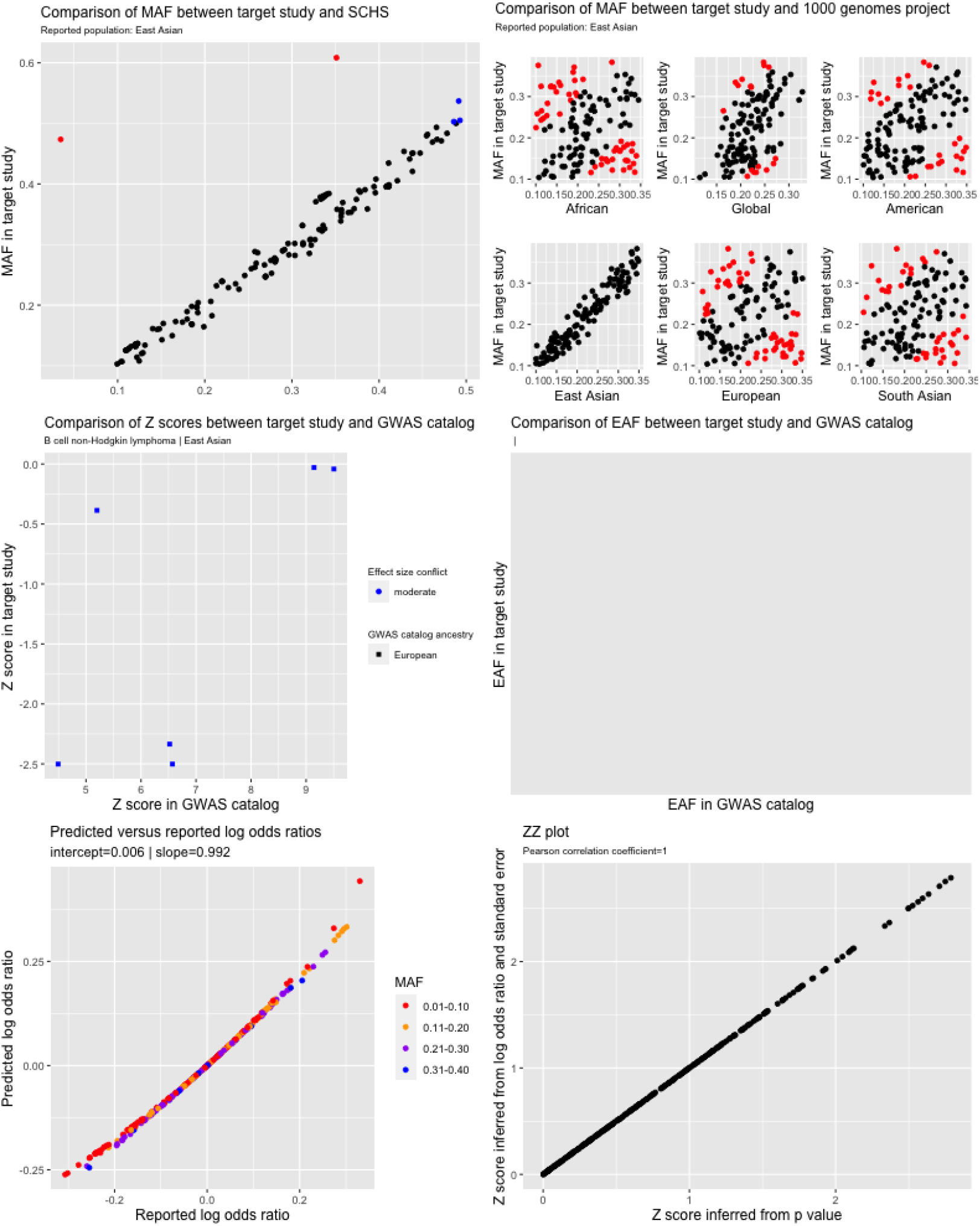
Quality control report for genetic summary data from a genome-wide association B cell non-Hodgkin lymphoma in the BC-NHL study (dataset ID 5) In the allele frequency subplots, each red data point corresponds to SNPs with high allele frequency conflicts, due to having an allele frequency that is greater than 0.58 (when it is expected to be less than 0.5) or to deviation from the reference allele frequency by more than 10 points

**Figure 5.**
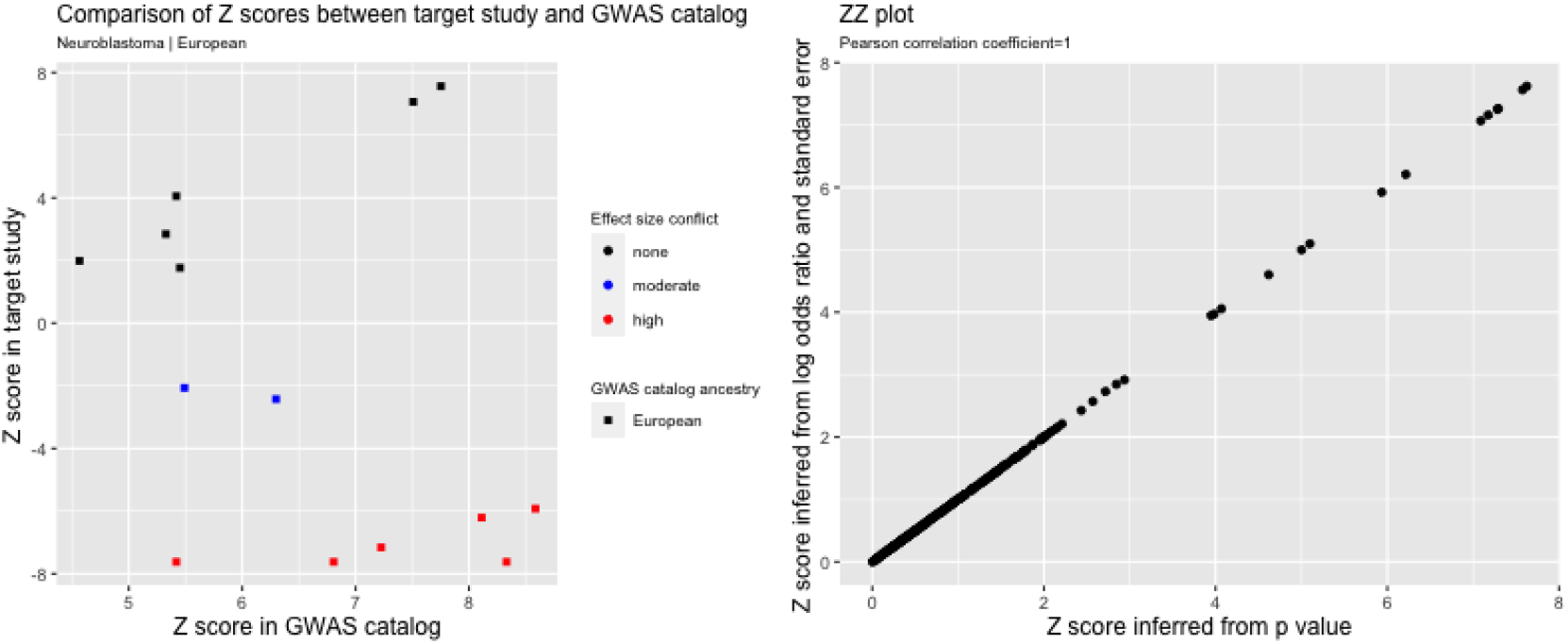
Quality control report for genetic summary data from a genome-wide association of neuroblastoma in the NBS dataset (ID 106)

We identified three datasets where reported allele frequency was inconsistent with allele frequency in reference datasets (**Figures 3, 6 & 7**). This included GliomaScan (ID 967) where allele frequency was inversely correlated with allele frequency across all SNPs in ancestry matched reference datasets, indicating that the reported effect allele frequency corresponded to the non-effect allele (**Figure 3**). In the UCSF_AGS/SFAGS (ID 133) and TNC (ID=132), there were two groups of SNPs showing positive or inverse correlations with allele frequency in ancestry matched datasets (**Figures 6 & 7)**, indicating that the reported effect allele frequency actually corresponds to minor allele frequency and that the minor allele was not consistently the effect allele. For these datasets, we decided to set effect allele frequency to missing. Allele frequency conflicts were not observed for other cancer datasets (**Supplementary figures 8-11**).

**Figure 6.**
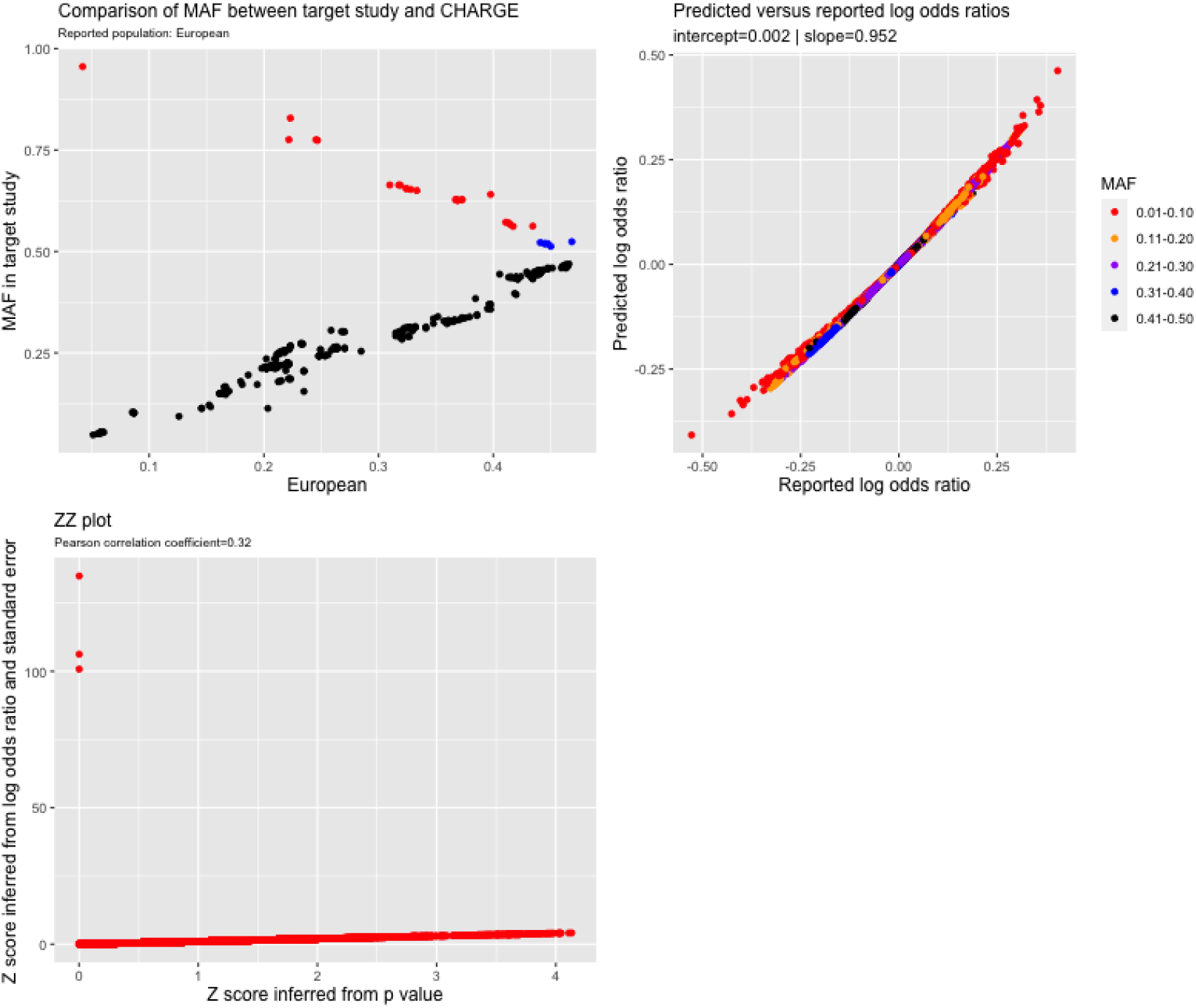
Quality control report for genetic summary data from a genome-wide association of glioma in the UCSF_AGS/SFAGS dataset (ID 133) In the allele frequency subplots, each red data point corresponds to SNPs with high allele frequency conflicts, due to having an allele frequency that is greater than 0.58 (when it is expected to be less than 0.5) or to deviation from the reference allele frequency by more than 10 points

**Figure 7.**
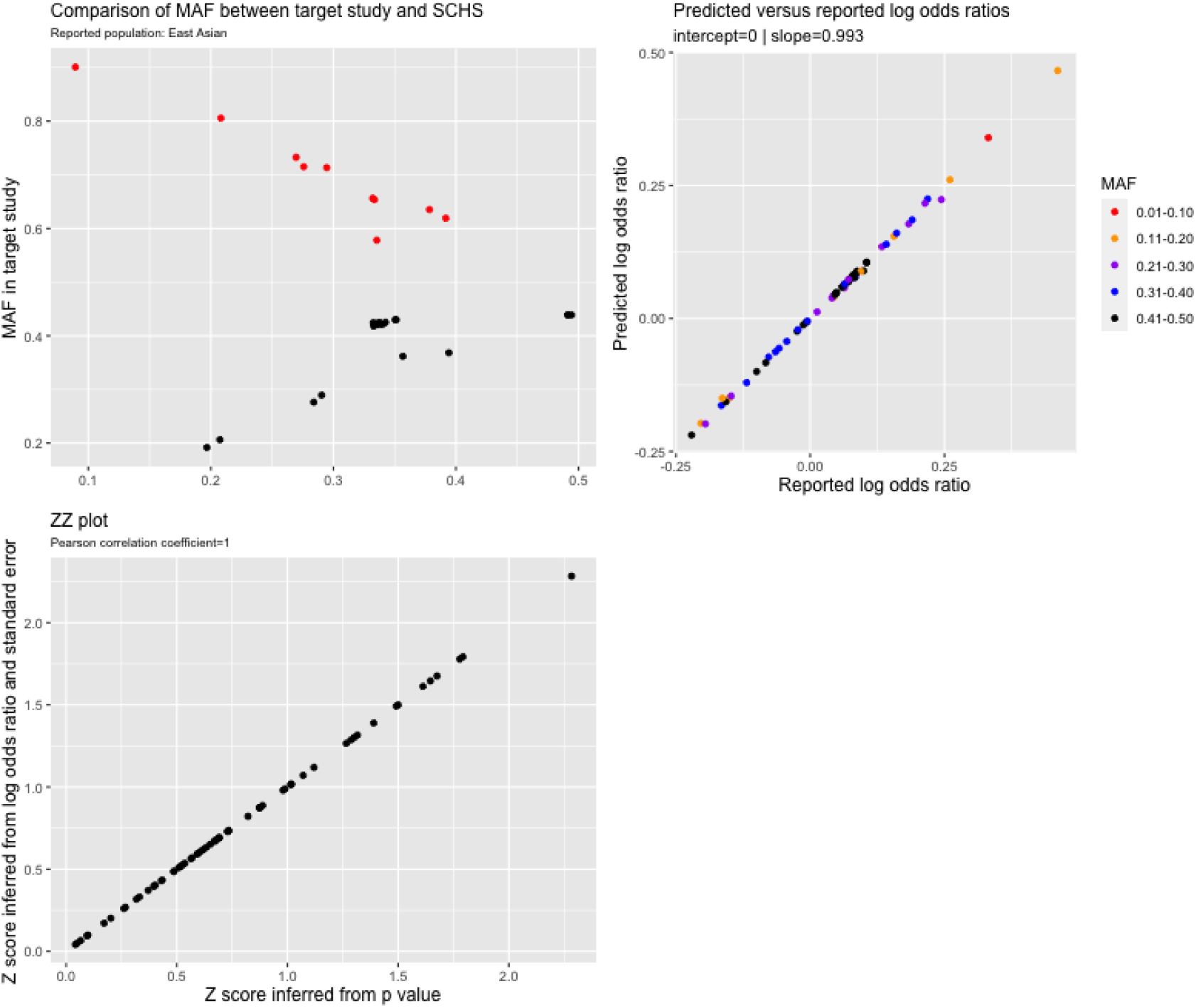
Quality control report for genetic summary data from a genome-wide association study of nasopharyngeal carcinoma in the TNC dataset (ID=132) In the allele frequency subplots, each red data point corresponds to SNPs with high allele frequency conflicts, due to having an allele frequency that is greater than 0.58 (when it is expected to be less than 0.5) or to deviation from the reference allele frequency by more than 10 points

We identified 35 datasets where the slope was >1.2 or <0.8 in models that regressed the expected log odds ratio on the reported effect size (**Supplementary figure 12**). Of these, 10 had slopes greater than 23. Further investigation revealed that the summary data for these 10 datasets had been generated in linear mixed models of cancer in participants from UK Biobank. Effect sizes from such models can be interpreted as the change in absolute risk per copy of the effect allele, explaining the conflict with the expected log odds ratio. We retained these datasets but transformed the reported effect size into a log odds ratio scale.

Of the remaining 25 datasets, all had slopes <0.8 and we confirmed that the reported effect sizes were log odds ratios by consulting the original study publications. For 18 of the 25 datasets, the deviation between the expected and reported log odds ratio was largely attributable to SNP-level sample size reporting errors, low imputation quality or a small number of SNPs with unusually large log odds ratios (>1 or <-1). For example, in the ACCC dataset (ID 3) with results for 20,952 SNPs from a meta-analysis of up-to 15 independent studies, results for a subset of 1562 SNPs had come from 5 or fewer studies (and therefore were analysed in a smaller subset of participants). When restricted to the subset of SNPs that came from ≥14 studies, the slope increased from 0.29 to 0.83 (**Supplementary figure 13**). In the GICC/MDA dataset (a meta-analysis of two independent studies), there were two distinct groups of SNPs, one group where results had come from both studies and a second group where results had come from a single study (**Supplementary figure 14**). When restricted to the dataset contributed to by both studies, the slope increased from 0.67 to 0.81. In the GECCO (IDs 59, 63, 64 and 65) and the HNMSC (IDs 70 & 71) datasets, the deviation of the slopes from one was at least partly driven by a very small number of outlier SNPs that had been derived from a subset of studies (for the HNMSC), or subset of genotyping batches (for GECCO) within each dataset’s respective meta-analysis (**Supplementary figures 15 & 16**). When restricted to the SNPs that had come from the maximum number of studies or genotyping batches, the slopes increased towards one. The slopes improved because the reported sample size was only applicable to those SNPs that were analysed in all studies or genotype batches. We decided to drop SNPs if the number of contributing studies or batches to that SNP was less than the median number of studies or batches. Across 16 datasets where this information was available, there were 9351 SNPs that met this criterion, including 868 in the fatty acid SNP set.

In the TNC (ID 132), NB-UGC (ID 104) and ECAC (ID 25) datasets, the deviation of the slopes from 1 was partly attributable to low imputation quality for some SNPs (**Supplementary figure 17**). We also found that the % deviation of the expected from the reported log odds ratio was strongly and inversely related to metrics of imputation quality (**Supplementary figure 18**) but not P values for deviation from Hardy Weinberg Equilibrium or P values for heterogeneity between studies (**Supplementary figures 19 & 20**). Across datasets where this information was available, there were 46,534 SNPs with imputation quality scores < 0.8, including 1119 in the fatty acid SNP set.

Effect size discrepancies in the OCAC and PRACTICAL datasets were mainly attributable to a very small number of SNPs with unusually large log odds ratios (>1 or < -1) (**Supplementary figure 12**). The number of SNPs across all datasets with log odds ratios greater than 1 or less than -1 was 368, including 5 SNPs in the fatty acid SNP set. Additional effect size discrepancies were identified in two datasets where the correlation was less than 0.99 between Z_p_ scores (Z scores from P values) and Z_b_ scores (Z scores inferred from effect sizes and standard errors) (**Figures 6 and 8**). In one dataset, this was due to three SNPs with very large effect sizes (Z>99) but with P values very close to 1 (>0.9). The second dataset showed a very irregular non-linear relationship between the two sets of Z scores (**Figure 8**). Correlations between the Zb and Zp scores were >0.99 across other cancer datasets **(Supplementary figure 21**).

**Figure 8.**
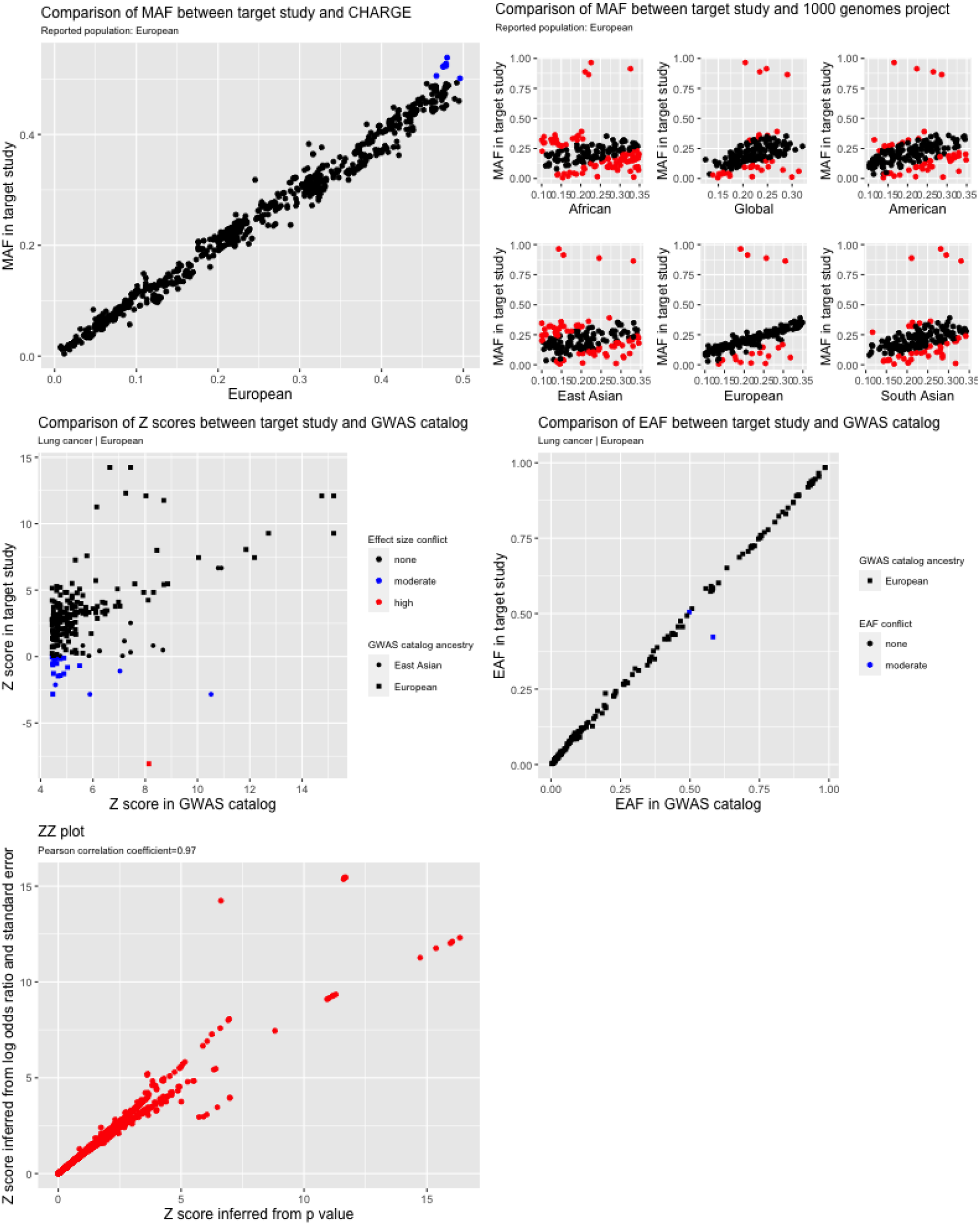
Quality control report for genetic summary data from a genome-wide association study of lung cancer in the ILCCO dataset (ID=74) In the allele frequency subplots, each red data point corresponds to SNPs with high allele frequency conflicts, due to having an allele frequency that is greater than 0.58 (when it is expected to be less than 0.5) or to deviation from the reference allele frequency by more than 10 points

### Final collection of cancer summary datasets

Overall, we identified 69,052 genetic associations (17.2% of the total) across 163 datasets with probable effect allele meta-data errors, low metrics of imputation quality (defined as info or imputation r^2^ score <0.8), large sample size errors, unusually large log odds ratios or effect sizes that didn’t closely correspond to their reported P values. Of these, 5,269 corresponded to the fatty acid SNP set (2.4%). An analytical issue, meta data error or summary data error was identified in 22 of 55 studies (6 fatty acid GWAS and 49 cancer GWAS). A serious error, which we define as having the potential to introduce substantial bias into MR analyses, was identified in 6 of 55 studies (10.9%). These included the effect allele meta data errors identified in the cancer studies and the large number of false positive genetic associations with fatty acids identified in CHARGE. After excluding unreliable associations and restricting the summary data to the fatty acid SNP set, there were 219,842 genetic associations with cancer in 160 datasets. The genetic associations represent analyses of 87 unique cancer types conducted in 609,863 cancer cases and 1,311,961 controls across 51 studies (**Table 1 and Supplementary table 5** ^17,55–96,99–103^. The median number of cases per cancer type was 3,101 (min =155, max= 139,445).

## Discussion

Our pipeline flagged analytical issues or meta and summary data errors in 23 studies (2 fatty acid GWAS and 21 cancer GWAS), including errors in 7 studies with the potential to introduce substantial bias into downstream MR analyses. Issues identified included a large number of false positive genetic associations for fatty acids, incorrect specification of the effect allele and effect allele frequency columns, inconsistent effect size scales amongst cancer studies, reported effect sizes and standard errors that did not correspond to reported P values, large numbers of low quality imputed SNPs, SNPs with incorrect sample sizes and SNPs with unusually large effect sizes.

### Effect allele meta data errors

Of the issues identified, incorrect specification of the effect allele column is the most serious, as it will lead to inferences of causal effect in the wrong direction^105,106^ (when the null hypothesis is false), and was flagged in 3 (6.1%) of the 49 cancer studies. A related, albeit less serious, error is incorrect specification of the effect allele frequency column, which can cause harmonisation problems for palindromic SNPs. Failure to harmonise palindromic SNPs between exposure and outcome studies may lead to increased heterogeneity in MR findings, which could in turn bias results towards the null (assuming the null hypothesis is false and that the palindromic SNPs are valid instruments). A conventional approach for avoiding these meta data errors is to compare allele frequency between the GWAS of interest and an external reference dataset^13^ or to confirm the effect allele through correspondence with study authors (especially when these are ambiguously labelled) or through consultation of readme files. Despite performing all of the latter checks, 5 (10.2%) of the 49 cancer studies were still affected by effect allele meta-data errors. One of the meta data errors was introduced by the MR data analyst whereas others were potentially due to human error by data providers. Our approach of comparing summary associations statistics for known “top hits” between the GWAS of interest and the GWAS catalog offers an additional safeguard against such errors.

### False positive GWAS hits

False positive genetic associations for fatty acids were identified in two of six fatty acid consortia. Failure to account for false positive hits could lead to the inclusion of genetic variants in MR analyses that are not truly associated with the exposure (a violation of instrumental variable assumptions, [see Box 1]), which could have the effect of biasing MR findings towards the null (assuming the null hypothesis is false). The false positives arose because we designed our instruments using the full summary association statistics, downloaded from the consortium website or obtained via correspondence, that had not gone through post-GWAS filtering procedures (e.g. exclusion of low frequency or low imputation quality variants). This instrument design strategy is probably more susceptible to inclusion of false positive genetic associations, compared to using the manually curated findings described in a GWAS publication. The latter are subject to relatively rigorous reporting standards, whereas there is little consensus on the format that GWAS results should take when posted to study-specific websites. Online platforms and databases that aggregate full summary association statistics from different studies may also be susceptible to this kind of error.

### Inconsistent effect size scales

We also found that cancer studies did not consistently express effect sizes as log odds ratios, with a substantial proportion of cancer analyses within UK Biobank expressing effect sizes as absolute changes in disease prevalence. The cancer analyses in question employed BOLT-LMM – a linear mixed model that allows the inclusion of related individuals and is more powerful and efficient than conventional regression procedures^42^ and is a widely used method for analysing binary disease traits in large-scale biobanks^107^. In general, failure to account for effect size scale differences will hamper comparison of findings amongst different diseases and could lead to the misinterpretation of results.

### Summary data errors

Potential summary data errors were flagged by mismatches between expected and reported effect sizes. We found that a substantial proportion of the mismatches were attributable to imputed SNPs, SNPs with incorrect sample sizes and SNPs with unusually large effect sizes. The sample size errors were due to the strategy of using the maximum reported sample size to represent sample size across all SNPs. However, not all samples in a given GWAS necessarily contribute to the analysis of every SNP, which is particularly common in large meta-analysis consortia with many independent studies. Incomplete sample overlap amongst SNPs within a GWAS could introduce bias into methods that assume a constant sample size, such as summary-data methods that rely on an external LD reference panel to model the correlation structure amongst SNPs in a genetic instrument. In the presence of incomplete sample overlap amongst SNPs, the use of an external LD reference panel could lead to the over-estimation of the covariance in SNP effect sizes. For example, in the most extreme case of zero sample overlap, the correlation in effect sizes for two SNPs will be zero even if those two SNPs are in LD^108^.

### General recommendations

When obtaining summary GWAS data via correspondence with study authors, we recommend that researchers should request access to full GWAS summary data, as this allows a far more comprehensive assessment of summary data reliability than is possible with only subsets of data. When full access is not possible, researchers should request summary data for SNPs that are established GWAS hits for their outcome of interest (i.e. not just the SNPs being used to instrument the exposure), which can then be used to confirm the identity of the effect allele through comparisons with the GWAS catalog. In addition, researchers could request summary data corresponding to the SNPs in our 1000 genomes reference set, which contains 2297 SNPs with the same minor allele across all 1000 genomes super populations, and which can be used to identify allele frequency issues. An advantage of using our 1000 genomes reference set is that effect allele frequency conflicts can be identified without knowledge of the ancestral background of the test dataset. Alternatively, a similar quality control check can be achieved by comparing allele frequencies between the exposure and outcome studies of interest (assuming they are closely matched on ancestry). Where possible, researchers should also confirm the identity of effect allele meta data through correspondence with the data providers.

We also recommend that researchers confirm with data providers the nature of all post GWAS filtering procedures that have been applied to the summary data. For example, in our own collaboration we ask each cancer study to confirm that their summary data has been through the same QC procedures described in their GWAS publications. Failure to perform this check could lead to the inclusion of large numbers of low quality and unreliable genetic associations. It is also advisable to confirm effect size scales, to support the correct interpretation of results. These considerations supplement previously developed guidelines for conducting MR studies^4,109,110^.

Our approach of comparing expected to reported effect sizes, and of comparing summary association statistics to external reference datasets, offers an additional safeguard against the aforementioned errors and analytical issues. A limitation of this approach is that not all flagged datasets will necessarily be problematic because other factors, such as covariate adjustment in the original GWAS or deviations from Hardy Weinberg equilibrium for reasons other than measurement error, could also cause deviations between expected and reported effect sizes. Therefore, SNPs flagged by this approach may still be suitable for downstream MR analyses. Another limitation is that our comparative approach may be less effective when there are zero, or few, known genetic associations for the trait of interest. This could happen, for example, when working with understudied or rare characteristics, for which existing GWAS may be underpowered. In such a situation, comparisons with genetic associations for closely related traits could still be informative. We manually mapped the text descriptions for each cancer type to the EFO, which could be inefficient when working with hundreds or thousands of traits. A more efficient approach would be to use the EMBL-EBI Zooma (https://www.ebi.ac.uk/spot/zooma) ontology mapping service, which supports command line access via a REST API.

### Two-sample population assumption

One of the key assumptions made in two-sample MR is that the studies used to define the exposure and the outcome come from the same population. The comparison of allele frequencies between test datasets and reference populations can in principle be used to evaluate this assumption. For example, in our own analyses, allele frequencies in the European origin cancer studies and 1000 genomes European super population were consistently strongly correlated (the same applied to the East Asian origin studies and the 1000 genomes East Asian super population), indicating that the reported study ancestries were broadly accurate. However, our QC procedure was not designed to specifically test for ancestral origins and was restricted to SNPs with a narrow allele frequency range. A more efficient approach would be to select SNPs with a much wider range of variation in minor allele frequency than chosen here. The need to assess the “same population” assumption is becoming more urgent with the growing diversity of GWAS, including a growing number of trans-ethnic and admixed studies.

## Conclusion

We have developed a QC pipeline that can be used to flag meta and summary data errors and a range of analytical issues in GWAS results, which in turn can be used to enhance the integrity of downstream two-sample MR analyses. We applied the pipeline to the FAMRC, identifying issues or errors in 42.9% of the studies. After resolving analytical issues and excluding problematic studies or unreliable genetic associations, we created a dataset of 219,842 genetic associations with 87 cancer types, derived from 48 separate studies, consortia or biobanks. The methods developed here are available to other researchers via the CheckSumStats R package (https://github.com/MRCIEU/CheckSumStats).

## Data Availability

Most of the datasets described here were obtained via Open GWAS, the GWAS catalog or the CHARGE website (see links below). Some datasets were obtained by correspondence with study authors and are available upon request.
https://gwas.mrcieu.ac.uk/
https://www.ebi.ac.uk/gwas/summary-statistics
https://www.chargeconsortium.com/main/results

https://gwas.mrcieu.ac.uk/

https://www.ebi.ac.uk/gwas/summary-statistics

https://www.chargeconsortium.com/main/results

## Acknowledgements

We gratefully acknowledge the participants and investigators of all studies that shared genetic summary data (further details of the studies can be found in **table 1 and supplementary 5**): the 23andMe Non-Melanoma Skin Cancer Study, Asian Colorectal Cancer Consortium, B Cell Childhood Acute Lymphoblastic Leukemia Study, B Cell Non-Hodgkin Lymphoma Study, Biobank Japan, Cohorts for Heart and Aging Research in Genomic Epidemiology Consortium, Childhood Acute Lymphoblastic Leukemia Study China Hepatocellular Carcinoma Study, Endometrial Cancer Association Consortium, EPITHYR, Esophageal Adenocarcinoma Study, Framingham study, FinnGen, Genetics and Epidemiology of Colorectal Cancer Consortium, Colorectal Transdisciplinary study, Colon Cancer Family Registry, Glioma International Case-Control Study, MD Anderson Cancer Center, GliomaScan, Harvard Non-Melanoma Skin Cancer Study, Hodgkin Lymphoma Study, InterLymph, International Head and Neck Cancer Epidemiology Consortium, KidRISK, Korean Chronic Myeloid Leukemia Study, Korean Hereditary Breast Cancer study, Malaysia Nasopharyngeal Carcinoma Study, Malignant Pleural Mesothelioma Study, Melanoma Meta-Analysis Consortium, Meta-analysis of Cervical Cancer Studies, Multiple Myeloma Study, Nanjing+Beijing Upper Gastrointestinal Cancers Study, NCI Upper Gastrointestinal Cancer Study, Neuroblastoma Study, Nijmegen Bladder Cancer Study, Pancreatic Cancer Case-Control Consortium, Pancreatic Cancer Cohort Consortium, Singapore Chinese Health Study, St. Jude Children’s Research Hospital and Children’s Oncology Group, Swedish Cervical Cancer Study, Taiwan Nasopharyngeal Carcinoma Study, The Breast Cancer Association Consortium, The International Lung Cancer Consortium, The Meningioma Consortium, The Ovarian Cancer Association Consortium, The Ovarian Cancer Association Consortium East Asian Subset, The Prostate Cancer Association Group to Investigate Cancer Associated Alterations in the Genome, Thyroid Cancer Study, UCSF Adult Glioma Study / San Francisco Adult Glioma Study, UCSF-Mayo GWAS, UK Biobank and the Uveal Melanoma Study. We gratefully acknowledge the help of Ramiro Magno for their help and advice with the gwasrapidd package.

Harvard cohorts (HPFS, NHS, PHS): The study protocol was approved by the institutional review boards of the Channing Division of Network Medicine, Department of Medicine, Brigham and Women’s Hospital and Harvard T.H. Chan School of Public Health, and those of participating registries as required. We would like to thank the participants and staff of the HPFS, NHS and PHS for their valuable contributions as well as the following state cancer registries for their help: AL, AZ, AR, CA, CO, CT, DE, FL, GA, ID, IL, IN, IA, KY, LA, ME, MD, MA, MI, NE, NH, NJ, NY, NC, ND, OH, OK, OR, PA, RI, SC, TN, TX, VA, WA, WY. The authors assume full responsibility for analyses and interpretation of these data.

Quality Control filtering of the UK Biobank data was conducted by R.Mitchell, G.Hemani, T.Dudding, L.Corbin, S.Harrison, L.Paternoster as described in the published protocol (doi: 10.5523/bris.1ovaau5sxunp2cv8rcy88688v). The MRC IEU UK Biobank GWAS pipeline was developed by B.Elsworth, R.Mitchell, C.Raistrick, L.Paternoster, G.Hemani and T.Gaunt (doi: 10.5523/bris.pnoat8cxo0u52p6ynfaekeigi).

## Disclaimer

Where authors are identified as personnel of the International Agency for Research on Cancer / World Health Organization, the authors alone are responsible for the views expressed in this article and they do not necessarily represent the decisions, policy or views of the International Agency for Research on Cancer / World Health Organization.

## References

1. Zheng J, Baird D, Borges M-C, et al. Recent Developments in Mendelian Randomization Studies. Curr Epidemiol Reports. 2017;4(4):330–345. doi:10.1007/s40471-017-0128-6

2. Burgess S, Small DS, Thompson SG. A review of instrumental variable estimators for Mendelian randomization. Stat Methods Med Res. 2017;26(5):2333–2355. doi:10.1177/0962280215597579

3. Haycock PC, Burgess S, Wade KH, Bowden J, Relton C, Davey Smith G. Best (but oft-forgotten) practices: the design, analysis, and interpretation of Mendelian randomization studies. Am J Clin Nutr. 2016;103(4):965–978. doi:10.3945/ajcn.115.118216

4. Hartwig FP, Davies NM, Hemani G, Smith GD. Counterfactual causation: Avoiding the downsides of a powerful, widely applicable but potentially fallible technique. Int J Epidemiol. 2016;45(6):1717–1726. doi:10.1093/ije/dyx028

5. Lyon MS, Andrews SJ, Elsworth B, Gaunt TR, Hemani G, Marcora E. The variant call format provides efficient and robust storage of GWAS summary statistics. Genome Biol. 2021;22(1):32. doi:10.1186/s13059-020-02248-0

6. Zheng J, Haberland V, Baird D, et al. Phenome-wide Mendelian randomization mapping the influence of the plasma proteome on complex diseases. Nat Genet. 2020;52(10). doi:10.1038/s41588-020-0682-6

7. Kazmi N, Haycock P, Tsilidis K, et al. Appraising causal relationships of dietary, nutritional and physical-activity exposures with overall and aggressive prostate cancer: Two-sample Mendelian-randomization study based on 79 148 prostate-cancer cases and 61 106 controls. Int J Epidemiol. 2020;49(2):587–596. doi:10.1093/IJE/DYZ235

8. Saunders CN, Cornish AJ, Kinnersley B, et al. Searching for causal relationships of glioma: a phenome-wide Mendelian randomisation study. Br J Cancer. 2021;124(2):447–454. doi:10.1038/s41416-020-01083-1

9. Kazmi N, Haycock P, Tsilidis K, et al. Appraising causal relationships of dietary, nutritional and physical-activity exposures with overall and aggressive prostate cancer: two-sample Mendelian-randomization study based on 79[148 prostate-cancer cases and 61[106 controls. Int J Epidemiol. 2020;49(2):587–596. doi:10.1093/ije/dyz235

10. Yuan S, Larsson SC. An atlas on risk factors for type 2 diabetes: a wide-angled Mendelian randomisation study. Diabetologia. 2020;63(11):2359–2371. doi:10.1007/s00125-020-05253-x

11. Haycock PC, Burgess S, Nounu A, et al. Association between telomere length and risk of cancer and non-neoplastic diseases a mendelian randomization study. JAMA Oncol. 2017;3(5):636–651. doi:10.1001/jamaoncol.2016.5945

12. Anderson CA, Pettersson FH, Clarke GM, Cardon LR, Morris AP, Zondervan KT. Data quality control in genetic case-control association studies. Nat Protoc. 2010;5(9):1564–1573. doi:10.1038/nprot.2010.116

13. Winkler TW, Day FR, Croteau-Chonka DC, et al. Quality control and conduct of genome-wide association meta-analyses. Nat Protoc. 2014;9(5):1192–1212. doi:10.1038/nprot.2014.071

14. Buniello A, Macarthur JAL, Cerezo M, et al. The NHGRI-EBI GWAS Catalog of published genome-wide association studies, targeted arrays and summary statistics 2019. Nucleic Acids Res. 2019;47(D1):D1005–D1012. doi:10.1093/nar/gky1120

15. Hemani G, Zheng J, Elsworth B, et al. The MR-Base platform supports systematic causal inference across the human phenome. Elife. 2018;7. doi:10.7554/eLife.34408

16. Hemani G. ieugwasr: R interface to the IEU GWAS database API. 2021. https://github.com/mrcieu/ieugwasr.

17. Ishigaki K, Akiyama M, Kanai M, et al. Large-scale genome-wide association study in a Japanese population identifies novel susceptibility loci across different diseases. Nat Genet. 2020;52(7):669–679. doi:10.1038/s41588-020-0640-3

18. Tanikawa C, Kamatani Y, Takahashi A, et al. GWAS identifies two novel colorectal cancer loci at 16q24.1 and 20q13.12. Carcinogenesis. 2018;39(5):652–660. doi:10.1093/carcin/bgy026

19. Nagai A, Hirata M, Kamatani Y, et al. Overview of the BioBank Japan Project: Study design and profile. J Epidemiol. 2017;27(3):S2–S8. doi:10.1016/j.je.2016.12.005

20. Mitchell Ruth, Hemani Gibran, Dudding Tom, Corbin Laura, Harrison Sean PL. UK Biobank Genetic Data: MRC-IEU Quality Control, version 2 - Datasets - data.bris. https://data.bris.ac.uk/data/dataset/1ovaau5sxunp2cv8rcy88688v. Published 2019. Accessed June 10, 2021.

21. Ruth, Mitchell; Elsworth, BL; Raistrick, CA; Paternoster, L; Hemani, G; Gaunt T. MRC IEU UK Biobank GWAS pipeline version 2 - Datasets - data.bris. https://data.bris.ac.uk/data/dataset/pnoat8cxo0u52p6ynfaekeigi. Published 2019. Accessed June 10, 2021.

22. Malone J, Holloway E, Adamusiak T, et al. Modeling sample variables with an Experimental Factor Ontology. Bioinformatics. 2010;26(8):1112–1118. doi:10.1093/bioinformatics/btq099

23. Wickham H. Ggplot2: Elegant Graphics for Data Analysis. New York: Springer-Verlag; 2016.

24. Durinck S, Moreau Y, Kasprzyk A, et al. BioMart and Bioconductor: a powerful link between biological databases and microarray data analysis. Bioinformatics. 2005;21(16):3439–3440. doi:10.1093/bioinformatics/bti525

25. Durinck S, Spellman PT, Birney E, Huber W. Mapping identifiers for the integration of genomic datasets with the R/Bioconductor package biomaRt. Nat Protoc. 2009;4(8):1184–1191. doi:10.1038/nprot.2009.97

26. Auguie B. gridExtra: Miscellaneous Functions for “Grid” Graphics. R package version 2.3.

27. Wilke CO. cowplot: Streamlined Plot Theme and Plot Annotations for “ggplot2.”2020. https://cran.r-project.org/package=cowplot.

28. Auguie B. gridExtra: Miscellaneous Functions for “Grid” Graphics. 2017. https://cran.r-project.org/package=gridExtra.

29. Henry L, Wickham H. purrr: Functional Programming Tools. 2020. https://cran.r-project.org/package=purrr.

30. Wickham H, François R, Henry L, Müller K. dplyr: A Grammar of Data Manipulation. 2021. https://cran.r-project.org/package=dplyr.

31. Müller K, Wickham H. tibble: Simple Data Frames. 2021. https://cran.r-project.org/package=tibble.

32. Bache SM, Wickham H. magrittr: A Forward-Pipe Operator for R. 2020. https://cran.r-project.org/package=magrittr.

33. Magno R, Maia A-TT. gwasrapidd: an R package to query, download and wrangle GWAS Catalog data. Wren J, ed. Bioinformatics. 2019;36(2):1–2. doi:10.1093/bioinformatics/btz605

34. Guan W, Steffen BT, Lemaitre RN, et al. Genome-Wide association study of plasma n6 polyunsaturated fatty acids within the cohorts for heart and aging research in genomic epidemiology consortium. Circ Cardiovasc Genet. 2014;7(3):321–331. doi:10.1161/CIRCGENETICS.113.000208

35. Dorajoo R, Sun Y, Han Y, et al. A genome-wide association study of n-3 and n-6 plasma fatty acids in a Singaporean Chinese population. Genes Nutr. 2015;10(6):53. doi:10.1007/s12263-015-0502-2

36. Zhu J, Manichaikul A, Hu Y, et al. Meta-analysis of genome-wide association studies identifies three novel loci for saturated fatty acids in East Asians. Eur J Nutr. 2016. doi:10.1007/s00394-016-1193-1

37. Tintle NL, Pottala J V., Lacey S, et al. A genome-wide association study of saturated, mono-and polyunsaturated red blood cell fatty acids in the Framingham Heart Offspring Study. Prostaglandins Leukot Essent Fat Acids. 2015;94:65–72. doi:10.1016/j.plefa.2014.11.007

38. Kettunen J, Demirkan A, Würtz P, et al. Genome-wide study for circulating metabolites identifies 62 loci and reveals novel systemic effects of LPA. Nat Commun. 2016;7(1):11122. doi:10.1038/ncomms11122

39. Shin S-Y, Fauman EB, Petersen A-K, et al. An atlas of genetic influences on human blood metabolites. Nat Genet. 2014;46(6):543–550. doi:10.1038/ng.2982

40. Auton A, Abecasis GR, Altshuler DM, et al. A global reference for human genetic variation. Nature. 2015;526(7571):68–74. doi:10.1038/nature15393

41. Harrison S. Estimating an Odds Ratio from a GWAS only reporting the P value – Sean Harrison: Blog. https://seanharrisonblog.com/2020/04/11/estimating-an-odds-ratio-from-a-gwas-only-reporting-the-p-value/. Published 2020. Accessed December 1, 2020.

42. Loh PR, Kichaev G, Gazal S, Schoech AP, Price AL. Mixed-model association for biobank-scale datasets. Nat Genet. 2018;50(7):906–908. doi:10.1038/s41588-018-0144-6

43. Lemaitre RN, King IB, Kabagambe EK, et al. Genetic loci associated with circulating levels of very long-chain saturated fatty acids. J Lipid Res. 2015;56(1):176–184. doi:10.1194/jlr.M052456

44. de Oliveira Otto MC, Lemaitre RN, Sun Q, et al. Genome-wide association meta-analysis of circulating odd-numbered chain saturated fatty acids: Results from the CHARGE Consortium. Loor JJ, ed. PLoS One. 2018;13(5):e0196951. doi:10.1371/journal.pone.0196951

45. Tanaka T, Shen J, Abecasis GR, et al. Genome-wide association study of plasma polyunsaturated fatty acids in the InCHIANTI Study. Georges M, ed. PLoS Genet. 2009;5(1):e1000338. doi:10.1371/journal.pgen.1000338

46. Gieger C, Geistlinger L, Altmaier E, et al. Genetics meets metabolomics: a genome-wide association study of metabolite profiles in human serum. Gibson G, ed. PLoS Genet. 2008;4(11):e1000282. doi:10.1371/journal.pgen.1000282

47. Mychaleckyj JC, Nayak U, Colgate ER, et al. Multiplex genomewide association analysis of breast milk fatty acid composition extends the phenotypic association and potential selection of FADS1 variants to arachidonic acid, a critical infant micronutrient. J Med Genet. 2018;55(7):459–468. doi:10.1136/jmedgenet-2017-105134

48. Hu Y, Tanaka T, Zhu J, et al. Discovery and fine-mapping of loci associated with MUFAs through trans-ethnic meta-analysis in Chinese and European populations. J Lipid Res. 2017;58(5):974–981. doi:10.1194/jlr.P071860

49. Mozaffarian D, Kabagambe EK, Johnson CO, et al. Genetic loci associated with circulating phospholipid trans fatty acids: a meta-analysis of genome-wide association studies from the CHARGE Consortium. Am J Clin Nutr. 2015;101(2):398–406. doi:10.3945/ajcn.114.094557

50. Guan W, Steffen BT, Lemaitre RN, et al. Genome-wide association study of plasma N6 polyunsaturated fatty acids within the cohorts for heart and aging research in genomic epidemiology consortium. Circ Cardiovasc Genet. 2014;7(3):321–331. doi:10.1161/CIRCGENETICS.113.000208

51. Lemaitre RN, Tanaka T, Tang W, et al. Genetic loci associated with plasma phospholipid n-3 fatty acids: a meta-analysis of genome-wide association studies from the CHARGE Consortium. McCarthy MI, ed. PLoS Genet. 2011;7(7):e1002193. doi:10.1371/journal.pgen.1002193

52. Wu JHY, Lemaitre RN, Manichaikul A, et al. Genome-wide association study identifies novel loci associated with concentrations of four plasma phospholipid fatty acids in the de novo lipogenesis pathway: results from the Cohorts for Heart and Aging Research in Genomic Epidemiology (CHARGE) consortium. Circ Cardiovasc Genet. 2013;6(2):171–183. doi:10.1161/CIRCGENETICS.112.964619

53. Veenstra J, Kalsbeek A, Westra J, Disselkoen C, E. Smith C, Tintle N. Genome-Wide Interaction Study of Omega-3 PUFAs and Other Fatty Acids on Inflammatory Biomarkers of Cardiovascular Health in the Framingham Heart Study. Nutrients. 2017;9(8):900. doi:10.3390/nu9080900

54. Kettunen J, Tukiainen T, Sarin A-P, et al. Genome-wide association study identifies multiple loci influencing human serum metabolite levels. Nat Genet. 2012;44(3):269–276. doi:10.1038/ng.1073

55. Chahal HS, Wu W, Ransohoff KJ, et al. Genome-wide association study identifies 14 novel risk alleles associated with basal cell carcinoma. Nat Commun. 2016;7:12510. doi:10.1038/ncomms12510

56. Huyghe JR, Bien SA, Harrison TA, et al. Discovery of common and rare genetic risk variants for colorectal cancer. Nat Genet. 2019;51(1):76–87. doi:10.1038/s41588-018-0286-6

57. Rajaraman P, Melin BS, Wang Z, et al. Genome-wide association study of glioma and meta-analysis. Hum Genet. 2012;131(12):1877–1888. doi:10.1007/s00439-012-1212-0

58. Sud A, Thomsen H, Law PJ, et al. Genome-wide association study of classical Hodgkin lymphoma identifies key regulators of disease susceptibility. Nat Commun. 2017;8(1):1892. doi:10.1038/s41467-017-00320-1

59. Zhang M, Song F, Liang L, et al. Genome-wide association studies identify several new loci associated with pigmentation traits and skin cancer risk in European Americans. Hum Mol Genet. 2013;22(14):2948–2959. doi:10.1093/hmg/ddt142

60. McKay JD, Hung RJ, Han Y, et al. Large-scale association analysis identifies new lung cancer susceptibility loci and heterogeneity in genetic susceptibility across histological subtypes. Nat Genet. 2017;49(7):1126–1132. doi:10.1038/ng.3892

61. Lesseur C, Diergaarde B, Olshan AF, et al. Genome-wide association analyses identify new susceptibility loci for oral cavity and pharyngeal cancer. Nat Genet. 2016;48(12):1544–1550. doi:10.1038/ng.3685

62. Berndt SI, Camp NJ, Skibola CF, et al. Meta-analysis of genome-wide association studies discovers multiple loci for chronic lymphocytic leukemia. Nat Commun. 2016;7:10933. doi:10.1038/ncomms10933

63. Cerhan JR, Berndt SI, Vijai J, et al. Genome-wide association study identifies multiple susceptibility loci for diffuse large B cell lymphoma. Nat Genet. 2014;46(11):1233–1238. doi:10.1038/ng.3105

64. Skibola CF, Berndt SI, Vijai J, et al. Genome-wide association study identifies five susceptibility loci for follicular lymphoma outside the HLA region. Am J Hum Genet. 2014;95(4):462–471. doi:10.1016/j.ajhg.2014.09.004

65. Vijai J, Wang Z, Berndt SI, et al. A genome-wide association study of marginal zone lymphoma shows association to the HLA region. Nat Commun. 2015;6:5751. doi:10.1038/ncomms6751

66. Chahal HS, Lin Y, Ransohoff KJ, et al. Genome-wide association study identifies novel susceptibility loci for cutaneous squamous cell carcinoma. Nat Commun. 2016;7:12048. doi:10.1038/ncomms12048

67. Lee J-Y, Kim J, Kim S-W, et al. BRCA1/2-negative, high-risk breast cancers (BRCAX) for Asian women: genetic susceptibility loci and their potential impacts. Sci Rep. 2018;8(1):15263. doi:10.1038/s41598-018-31859-8

68. Scelo G, Purdue MP, Brown KM, et al. Genome-wide association study identifies multiple risk loci for renal cell carcinoma. Nat Commun. 2017;8:15724. doi:10.1038/ncomms15724

69. Leo PJ, Madeleine MM, Wang S, et al. Defining the genetic susceptibility to cervical neoplasia-A genome-wide association study. PLoS Genet. 2017;13(8):e1006866. doi:10.1371/journal.pgen.1006866

70. Claus EB, Cornish AJ, Broderick P, et al. Genome-wide association analysis identifies a meningioma risk locus at 11p15.5. Neuro Oncol. 2018;20(11):1485–1493. doi:10.1093/neuonc/noy077

71. Swaminathan B, Thorleifsson G, Jöud M, et al. Variants in ELL2 influencing immunoglobulin levels associate with multiple myeloma. Nat Commun. 2015;6:7213. doi:10.1038/ncomms8213

72. Chin Y-M, Tan LP, Abdul Aziz N, et al. Integrated pathway analysis of nasopharyngeal carcinoma implicates the axonemal dynein complex in the Malaysian cohort. Int J cancer. 2016;139(8):1731–1739. doi:10.1002/ijc.30207

73. Matullo G, Guarrera S, Betti M, et al. Genetic variants associated with increased risk of malignant pleural mesothelioma: a genome-wide association study. PLoS One. 2013;8(4):e61253. doi:10.1371/journal.pone.0061253

74. Wu C, Wang Z, Song X, et al. Joint analysis of three genome-wide association studies of esophageal squamous cell carcinoma in Chinese populations. Nat Genet. 2014;46(9):1001–1006. doi:10.1038/ng.3064

75. Hu N, Wang Z, Song X, et al. Genome-wide association study of gastric adenocarcinoma in Asia: a comparison of associations between cardia and non-cardia tumours. Gut. 2016;65(10):1611–1618. doi:10.1136/gutjnl-2015-309340

76. Wang Z, Dai J, Hu N, et al. Identification of new susceptibility loci for gastric non-cardia adenocarcinoma: pooled results from two Chinese genome-wide association studies. Gut. 2017;66(4):581–587. doi:10.1136/gutjnl-2015-310612

77. Lu Y, Kweon S-S, Cai Q, et al. Identification of Novel Loci and New Risk Variant in Known Loci for Colorectal Cancer Risk in East Asians. Cancer Epidemiol biomarkers Prev a Publ Am Assoc Cancer Res cosponsored by Am Soc Prev Oncol. 2020;29(2):477–486. doi:10.1158/1055-9965.EPI-19-0755

78. Rafnar T, Vermeulen SH, Sulem P, et al. European genome-wide association study identifies SLC14A1 as a new urinary bladder cancer susceptibility gene. Hum Mol Genet. 2011;20(21):4268–4281. doi:10.1093/hmg/ddr303

79. McDaniel LD, Conkrite KL, Chang X, et al. Common variants upstream of MLF1 at 3q25 and within CPZ at 4p16 associated with neuroblastoma. PLoS Genet. 2017;13(5):e1006787. doi:10.1371/journal.pgen.1006787

80. Phelan CM, Kuchenbaecker KB, Tyrer JP, et al. Identification of 12 new susceptibility loci for different histotypes of epithelial ovarian cancer. Nat Genet. 2017;49(5):680–691. doi:10.1038/ng.3826

81. Lawrenson K, Song F, Hazelett DJ, et al. Genome-wide association studies identify susceptibility loci for epithelial ovarian cancer in east Asian women. Gynecol Oncol. 2019;153(2):343–355. doi:10.1016/j.ygyno.2019.02.023

82. Klein AP, Wolpin BM, Risch HA, et al. Genome-wide meta-analysis identifies five new susceptibility loci for pancreatic cancer. Nat Commun. 2018;9(1):556. doi:10.1038/s41467-018-02942-5

83. Schumacher FR, Al Olama AA, Berndt SI, et al. Association analyses of more than 140,000 men identify 63 new prostate cancer susceptibility loci. Nat Genet. 2018;50(7):928–936. doi:10.1038/s41588-018-0142-8

84. Chen D, Juko-Pecirep I, Hammer J, et al. Genome-wide association study of susceptibility loci for cervical cancer. J Natl Cancer Inst. 2013;105(9):624–633. doi:10.1093/jnci/djt051

85. Treviño LR, Yang W, French D, et al. Germline genomic variants associated with childhood acute lymphoblastic leukemia. Nat Genet. 2009;41(9):1001–1005. doi:10.1038/ng.432

86. Tse K-PP, Su W-HH, Chang K-PP, et al. Genome-wide Association Study Reveals Multiple Nasopharyngeal Carcinoma-Associated Loci within the HLA Region at Chromosome 6p21.3. Am J Hum Genet. 2009;85(2):194–203. doi:10.1016/j.ajhg.2009.07.007

87. Melin BS, Barnholtz-Sloan JS, Wrensch MR, et al. Genome-wide association study of glioma subtypes identifies specific differences in genetic susceptibility to glioblastoma and non-glioblastoma tumors. Nat Genet. 2017;49(5):789–794. doi:10.1038/ng.3823

88. Vijayakrishnan J, Studd J, Broderick P, et al. Genome-wide association study identifies susceptibility loci for B-cell childhood acute lymphoblastic leukemia. Nat Commun. 2018;9(1):1340. doi:10.1038/s41467-018-03178-z

89. Zhou W, Nielsen JB, Fritsche LG, et al. Efficiently controlling for case-control imbalance and sample relatedness in large-scale genetic association studies. Nat Genet. 2018;50(9):1335–1341. doi:10.1038/s41588-018-0184-y

90. Mobuchon L, Battistella A, Bardel C, et al. A GWAS in uveal melanoma identifies risk polymorphisms in the CLPTM1L locus. NPJ genomic Med. 2017;2. doi:10.1038/s41525-017-0008-5

91. Mitchell JA, Warner TD. COX isoforms in the cardiovascular system: Understanding the activities of non-steroidal anti-inflammatory drugs. Nat Rev Drug Discov. 2006;5(1):75–86. doi:10.1038/nrd1929

92. Köhler A, Chen B, Gemignani F, et al. Genome-wide association study on differentiated thyroid cancer. J Clin Endocrinol Metab. 2013;98(10):E1674–81. doi:10.1210/jc.2013-1941

93. Tan DEK, Foo JN, Bei J-X, et al. Genome-wide association study of B cell non-Hodgkin lymphoma identifies 3q27 as a susceptibility locus in the Chinese population. Nat Genet. 2013;45(7):804–807. doi:10.1038/ng.2666

94. Kim DHD, Lee S-T, Won H-H, et al. A genome-wide association study identifies novel loci associated with susceptibility to chronic myeloid leukemia. Blood. 2011;117(25):6906–6911. doi:10.1182/blood-2011-01-329797

95. Law MH, Bishop DT, Lee JE, et al. Genome-wide meta-analysis identifies five new susceptibility loci for cutaneous malignant melanoma. Nat Genet. 2015;47(9):987–995. doi:10.1038/ng.3373

96. Truong T, Lesueur F, Sugier PE, et al. Multiethnic genome-wide association study of differentiated thyroid cancer in the EPITHYR consortium. Int J Cancer. 2021;148(12):2935–2946. doi:10.1002/ijc.33488

97. Li WQ, Hu N, Hyland PL, et al. Genetic variants in DNA repair pathway genes and risk of esophageal squamous cell carcinoma and gastric adenocarcinoma in a Chinese population. Carcinogenesis. 2013;34(7):1536–1542. doi:10.1093/carcin/bgt094

98. Ciampa J, Yeager M, Amundadottir L, et al. Large-scale exploration of gene-gene interactions in prostate cancer using a multistage genome-wide association study. Cancer Res. 2011;71(9):3287–3295. doi:10.1158/0008-5472.CAN-10-2646

99. Michailidou K, Lindström S, Dennis J, et al. Association analysis identifies 65 new breast cancer risk loci. Nature. 2017;551(7678):92–94. doi:10.1038/nature24284

100. Ellinghaus E, Stanulla M, Richter G, et al. Identification of germline susceptibility loci in ETV6-RUNX1-rearranged childhood acute lymphoblastic leukemia. Leukemia. 2012;26(5):902–909. doi:10.1038/leu.2011.302

101. Li S, Qian J, Yang Y, et al. GWAS identifies novel susceptibility loci on 6p21.32 and 21q21.3 for hepatocellular carcinoma in chronic hepatitis B virus carriers. PLoS Genet. 2012;8(7):e1002791. doi:10.1371/journal.pgen.1002791

102. Gharahkhani P, Fitzgerald RC, Vaughan TL, et al. Genome-wide association studies in oesophageal adenocarcinoma and Barrett’s oesophagus: a large-scale meta-analysis. Lancet Oncol. 2016;17(10):1363–1373. doi:10.1016/S1470-2045(16)30240-6

103. O’Mara TA, Glubb DM, Amant F, et al. Identification of nine new susceptibility loci for endometrial cancer. Nat Commun. 2018;9(1):3166. doi:10.1038/s41467-018-05427-7

104. Elsworth B, Lyon M, Alexander T, et al. The MRC IEU OpenGWAS data infrastructure. bioRxiv. August 2020:2020.08.10.244293. doi:10.1101/2020.08.10.244293

105. Inoshita M, Numata S, Tajima A, et al. A significant causal association between C-reactive protein levels and schizophrenia. Sci Rep. 2016;6. doi:10.1038/srep26105

106. Prins BP, Abbasi A, Wong A, et al. Investigating the Causal Relationship of C-Reactive Protein with 32 Complex Somatic and Psychiatric Outcomes: A Large-Scale Cross-Consortium Mendelian Randomization Study. PLoS Med. 2016;13(6):34. doi:10.1371/journal.pmed.1001976

107. Masuda T, Low SK, Akiyama M, et al. GWAS of five gynecologic diseases and cross-trait analysis in Japanese. Eur J Hum Genet. 2020;28(1):95–107. doi:10.1038/s41431-019-0495-1

108. Rüeger S, McDaid A, Kutalik Z. Evaluation and application of summary statistic imputation to discover new height-associated loci. PLoS Genet. 2018;14(5). doi:10.1371/journal.pgen.1007371

109. Davey Smith G, Davies N, Dimou N, et al. STROBE-MR: Guidelines for strengthening the reporting of Mendelian randomization studies. 2019. doi:10.7287/peerj.preprints.27857

110. Burgess S, Davey Smith G, Davies NM, et al. Guidelines for performing Mendelian randomization investigations. Wellcome Open Res. 2020;4. doi:10.12688/wellcomeopenres.15555.2

