## Supplementary Figures for "Design and quality control of large-scale two-sample Mendelian randomisation studies"

Supplementary figure 1. Quality control report for genetic summary data from a genome-wide association of arachidonic acid in the Framingham heart study

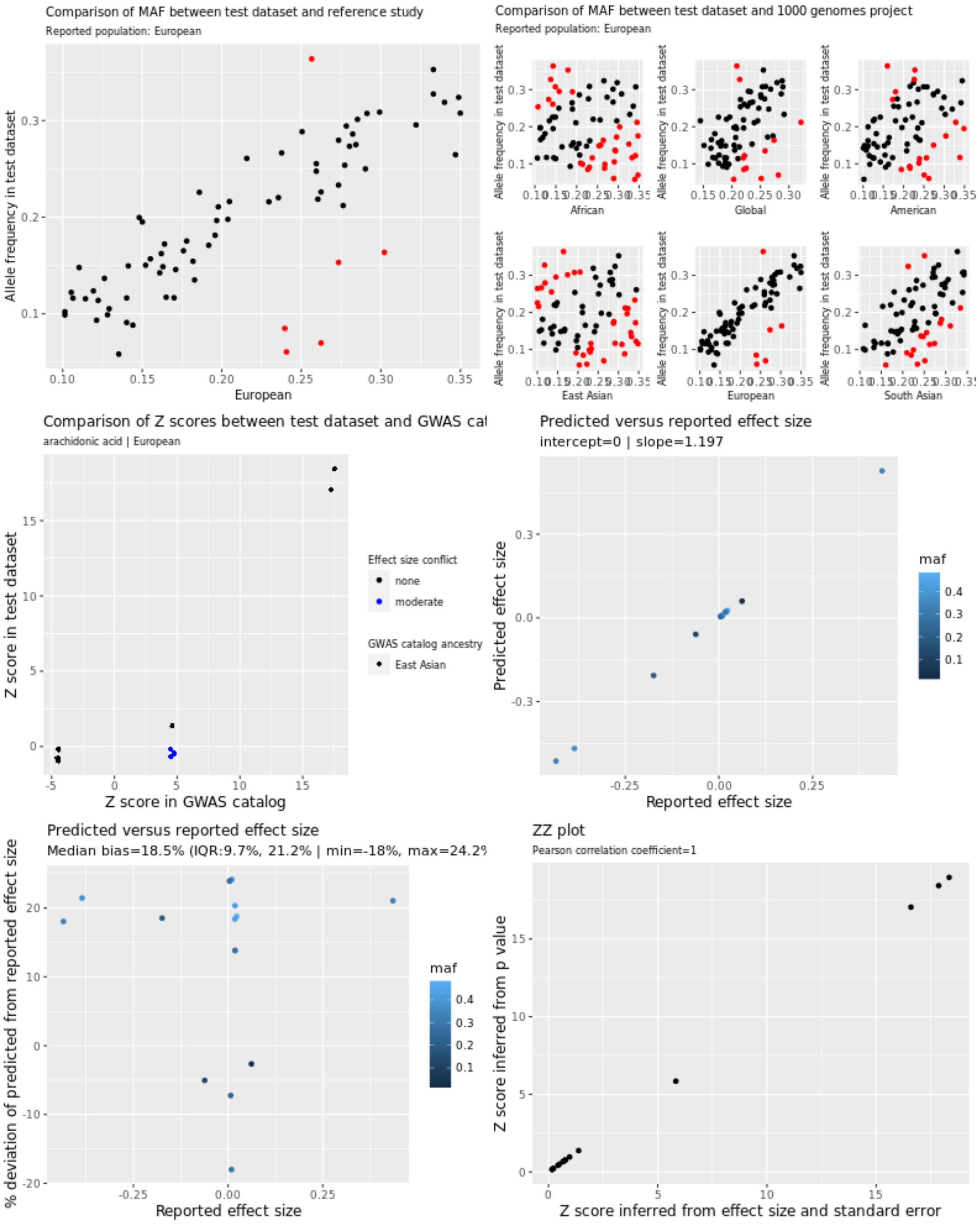


In the allele frequency plots, each red data point corresponds to SNPs with high allele frequency conflicts, due to deviation from the reference allele frequency by more than 10 points.

Supplementary figure 2. Quality control report for genetic summary data from a genome-wide association of linoleic acid the Kettunen study

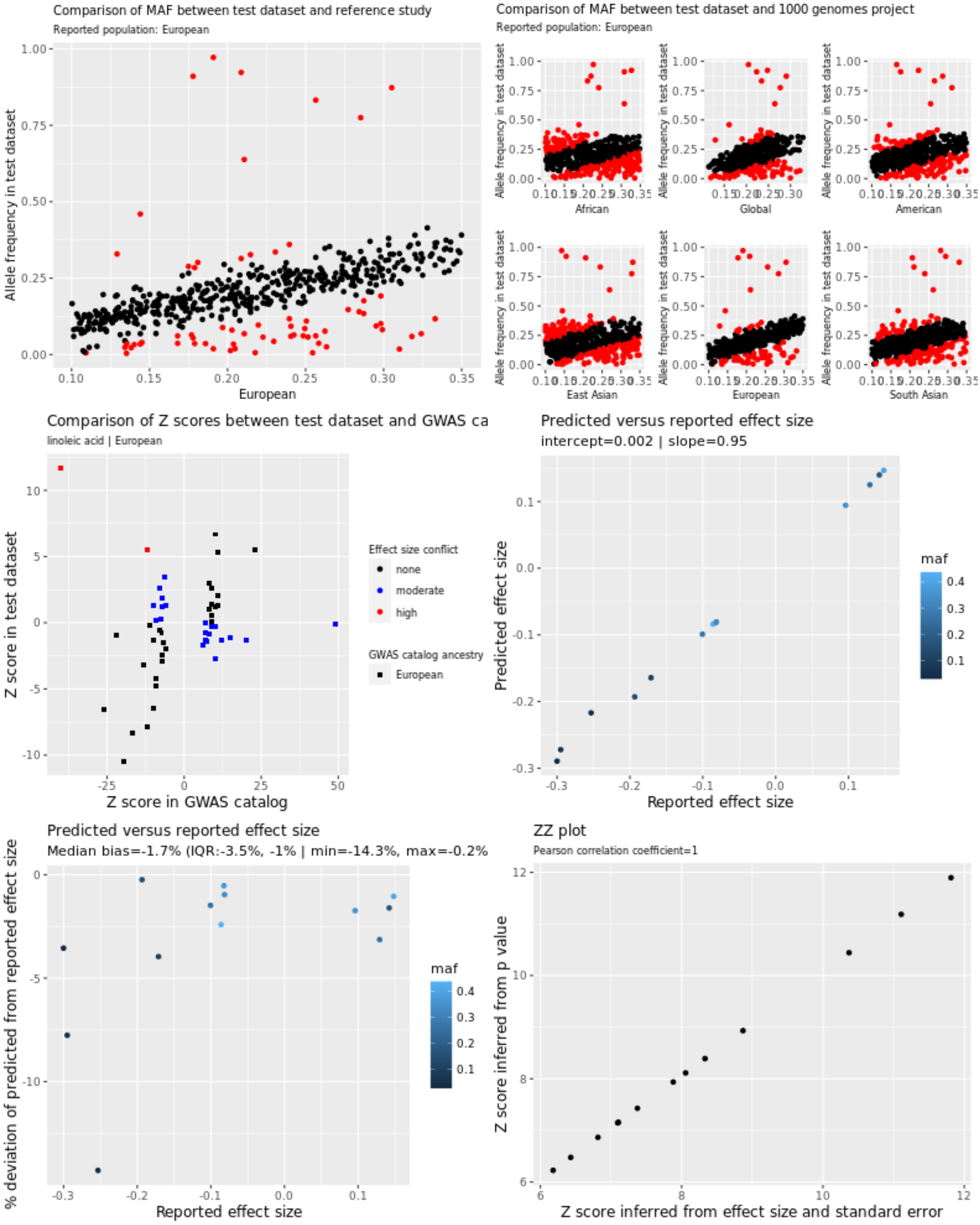


In the allele frequency plots, each red data point corresponds to SNPs with high allele frequency conflicts, due to having an allele frequency that is greater than 0.58 (when it is expected to be less than 0.5) or to deviation from the reference allele frequency by more than 10 points.

Supplementary figure 3. Quality control report for genetic summary data from a genome-wide association of arachidonic acid in the TwinsUK/KORA study

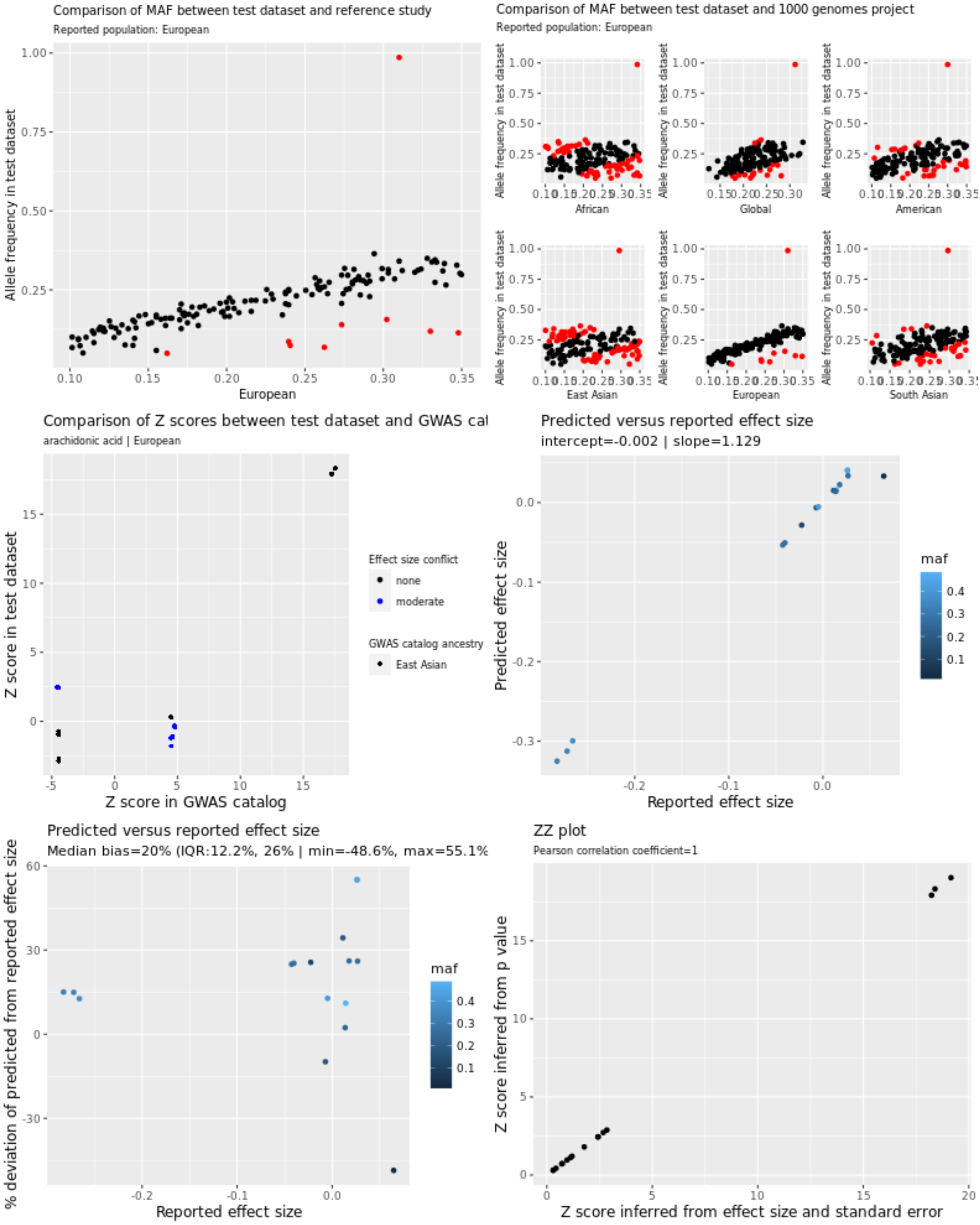


In the allele frequency plots, each red data point corresponds to SNPs with high allele frequency conflicts, due to having an allele frequency that is greater than 0.58 (when it is expected to be less than 0.5) or to deviation from the reference allele frequency by more than 10 points.

Supplementary figure 4. Quality control report for genetic summary data from a genome-wide association of arachidonic acid in the SCHS

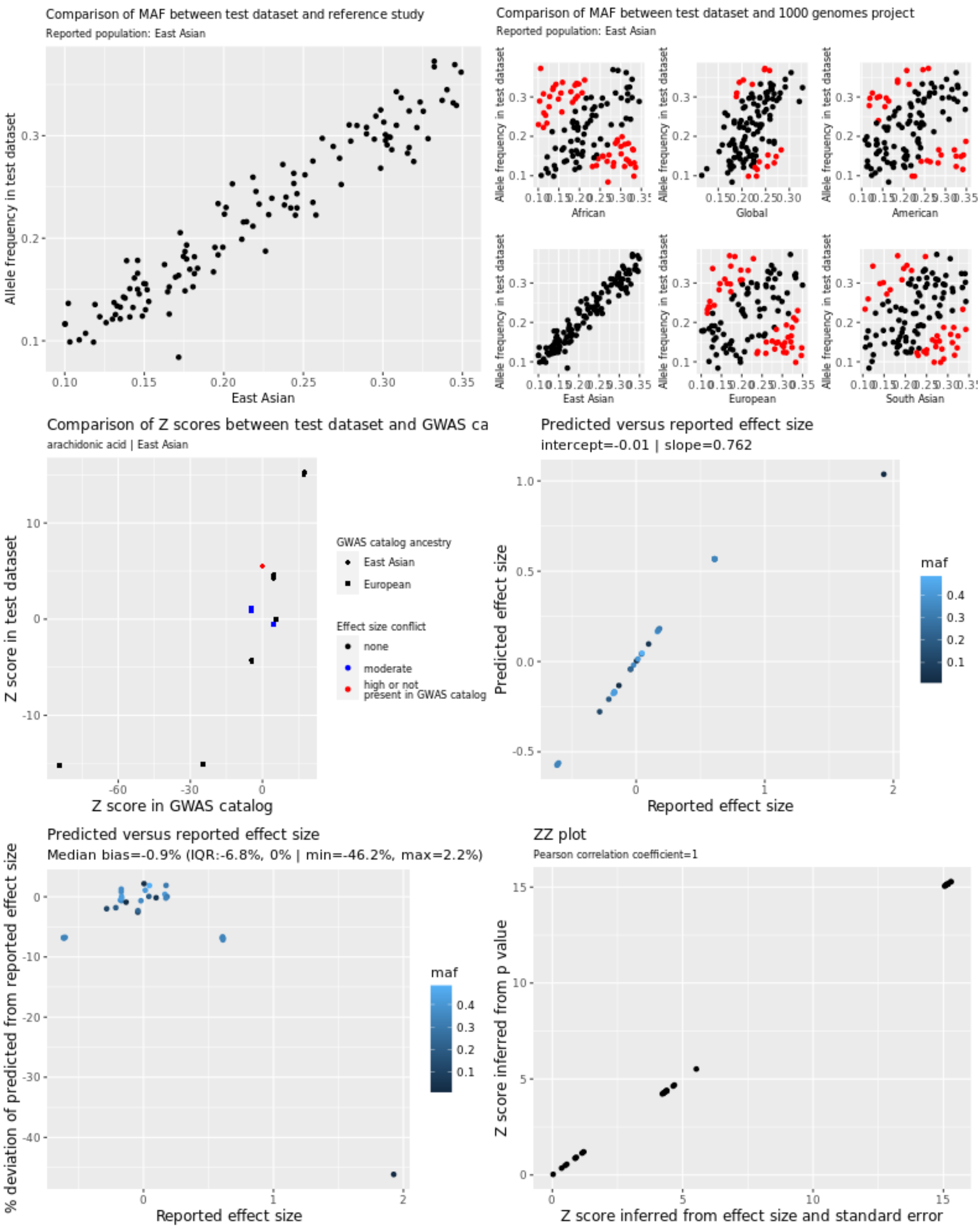


In the allele frequency plots, each red data point corresponds to SNPs with high allele frequency conflicts, due to deviation from the reference allele frequency by more than 10 points.

Supplementary figure 5. Quality control report for genetic summary data from a genome-wide association of stearic acid in the NHAPC/MESA-CHI study

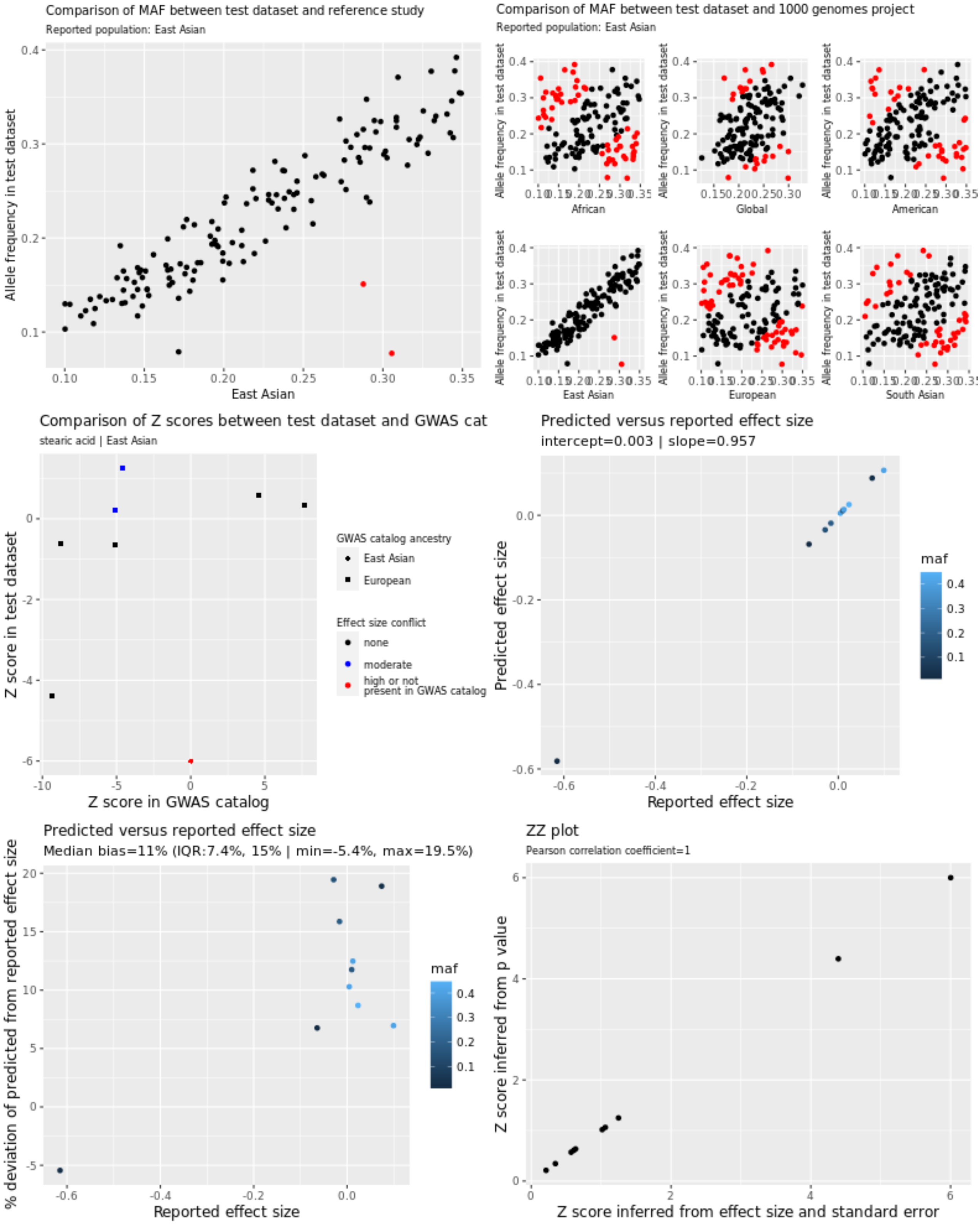


In the allele frequency plots, each red data point corresponds to SNPs with high allele frequency conflicts, due to deviation from the reference allele frequency by more than 10 points.

Supplementary figure 6. Relationship between reported and expected effect sizes for SNPs associated with arachidonic acid in CHARGE, before and after filtering out low quality SNPs

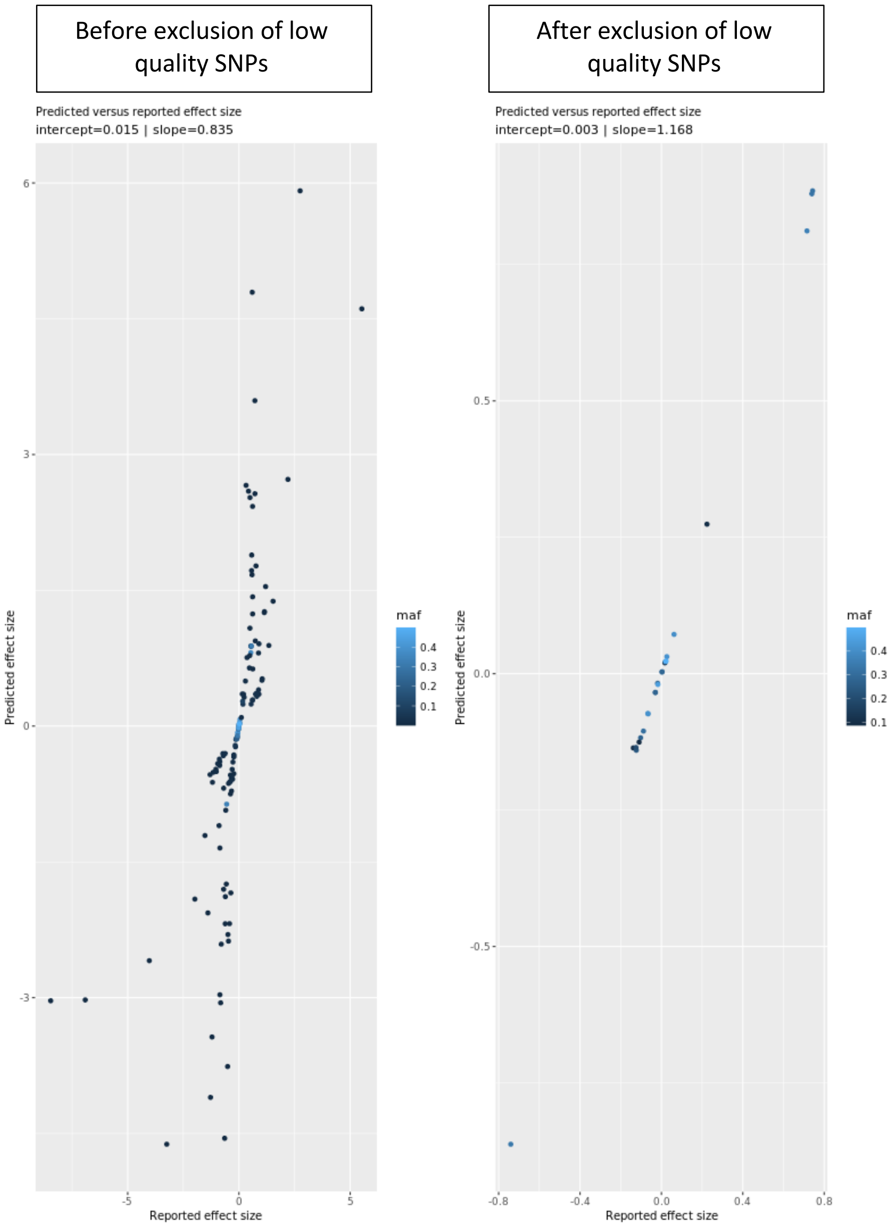

Supplementary Figure 7a. Comparison of effect sizes between cancer datasets (ID1-ID83) and the NHGRI-EBI GWAS catalog

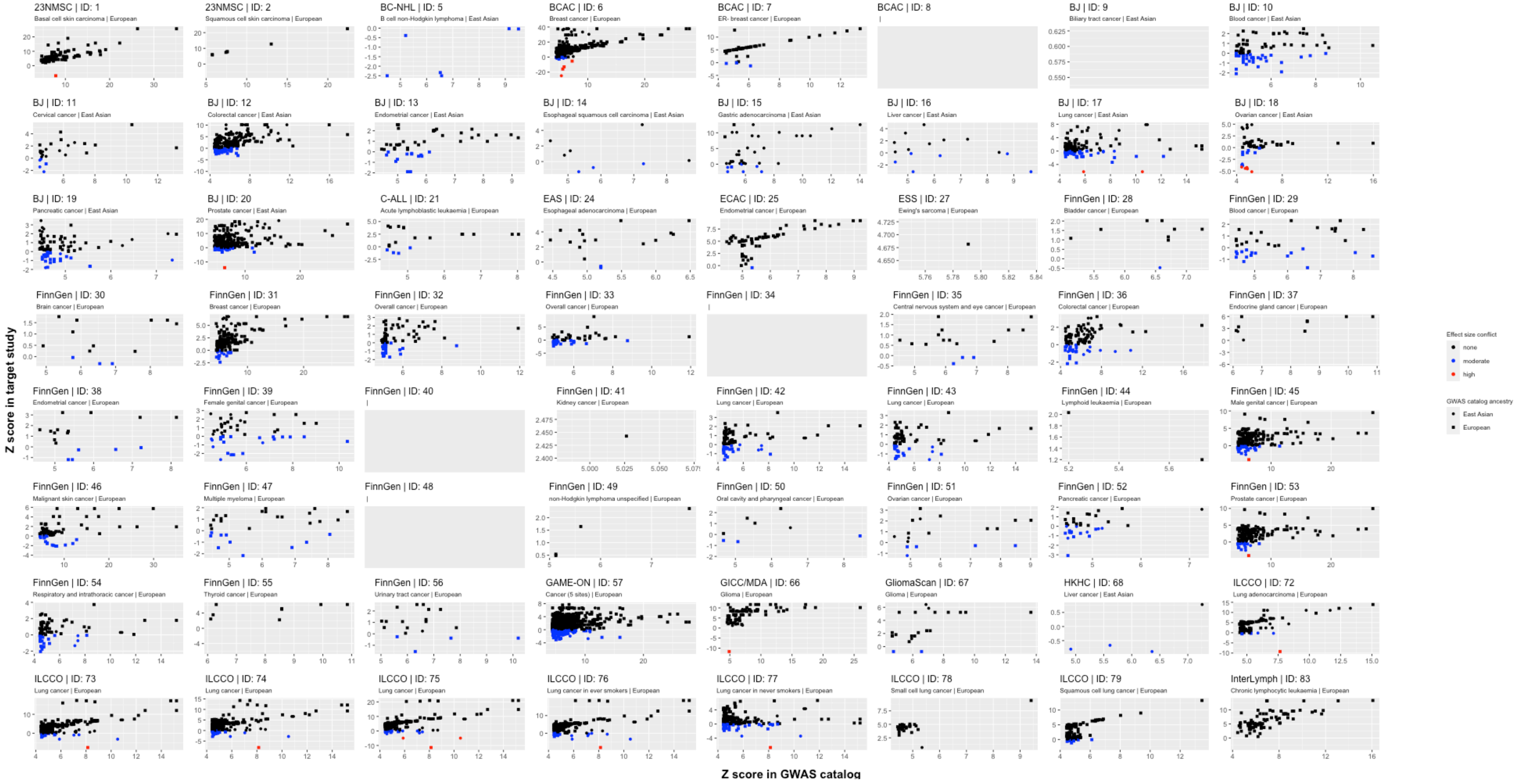

Supplementary Figure 7b. Comparison of effect sizes between cancer datasets (ID84-ID1499) and the NHGRI-EBI GWAS catalog

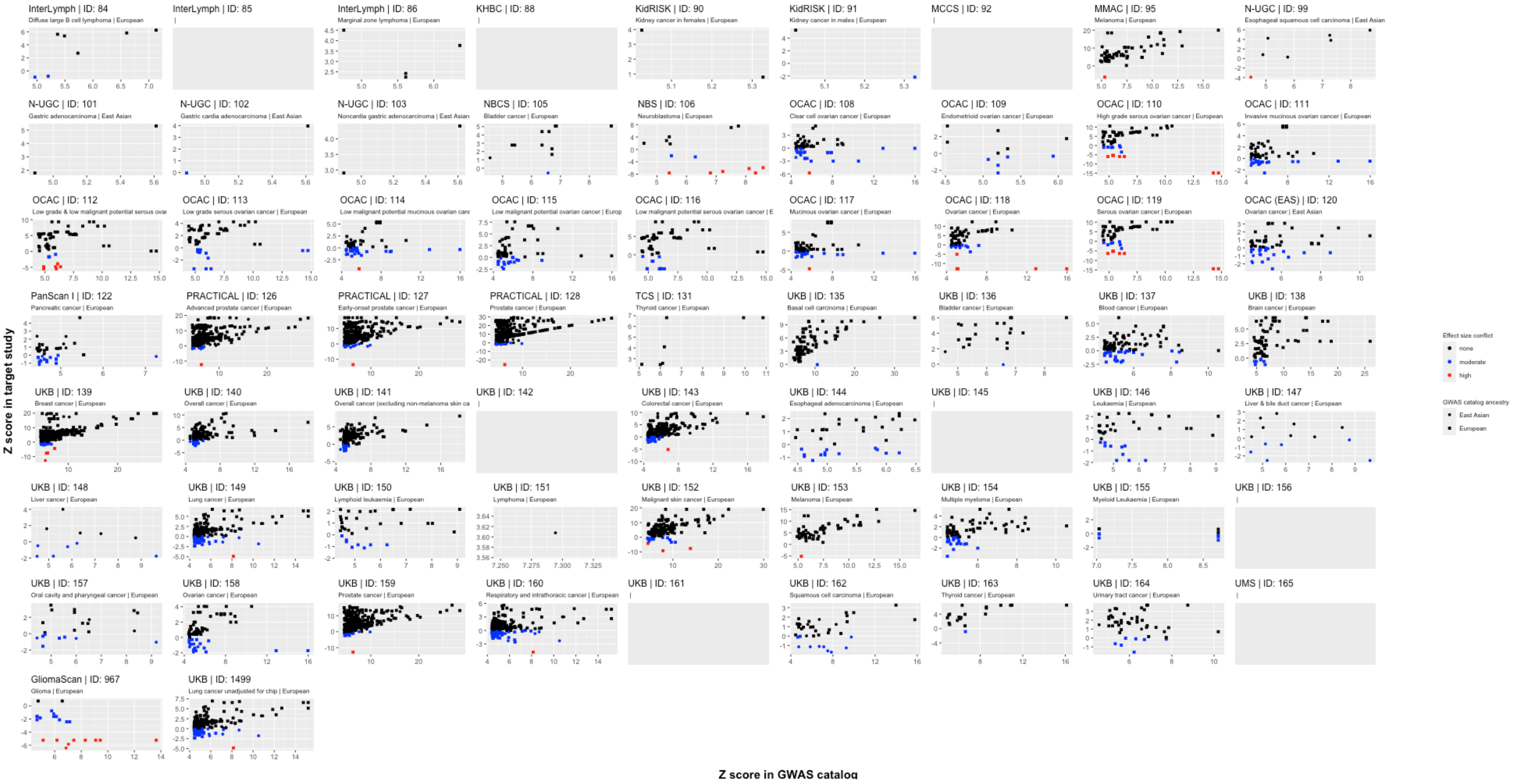

Supplementary Figure 8. Comparison of minor allele frequency between cancer datasets from European studies and the Cohorts for Heart and Aging Research in Genomic Epidemiology Consortium

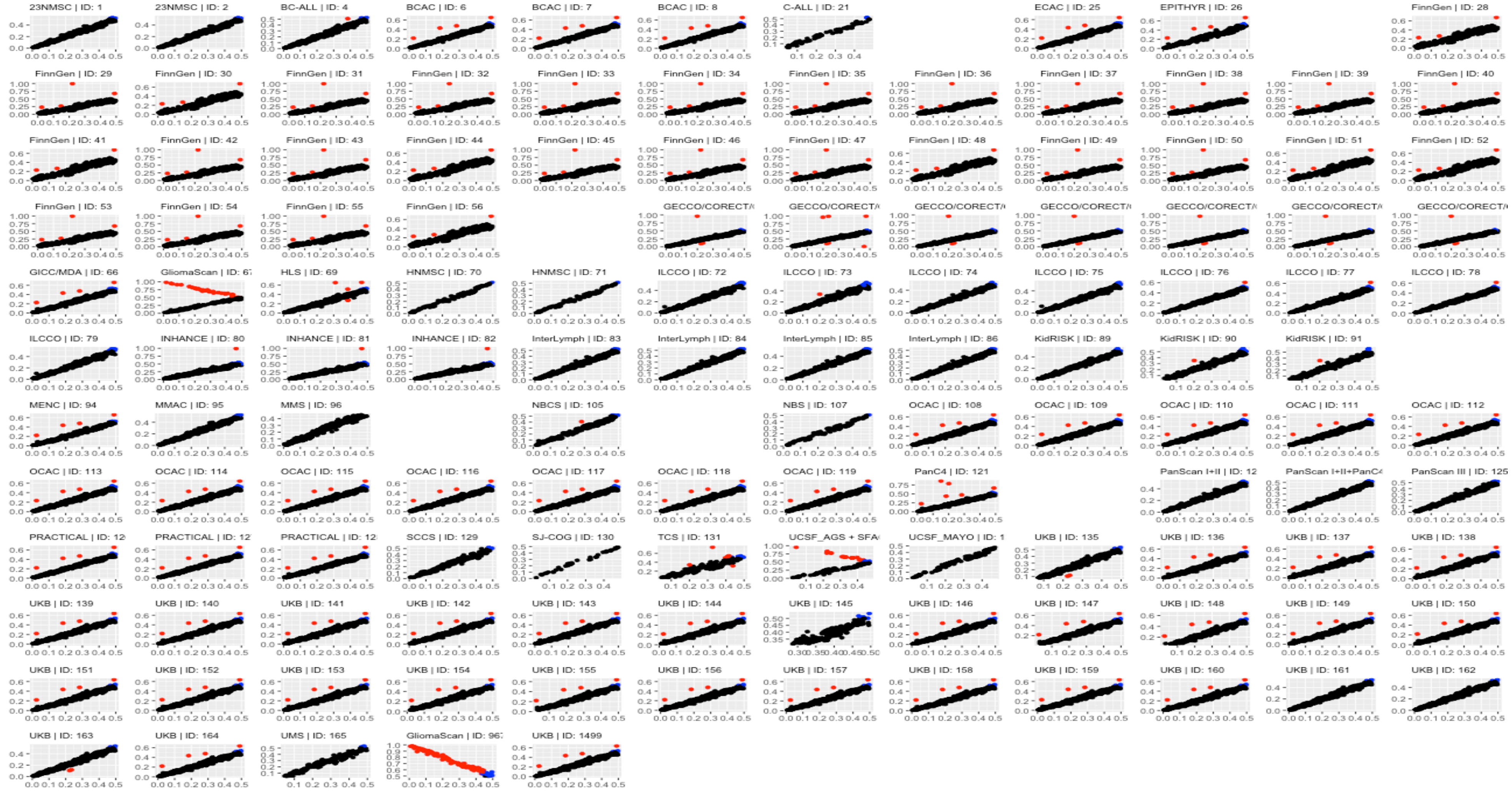


Red data points correspond to SNPs with high allele frequency conflicts, due to having an allele frequency that is greater than 0.58 (when it is expected to be less than 0.5) or to deviation from the reference allele frequency by more than 10 points. Study acronyms are explained in supplementary table 5.

Supplementary Figure 9. Comparison of minor allele frequency between cancer datasets from East Asian studies and the Singapore Chinese Health Study

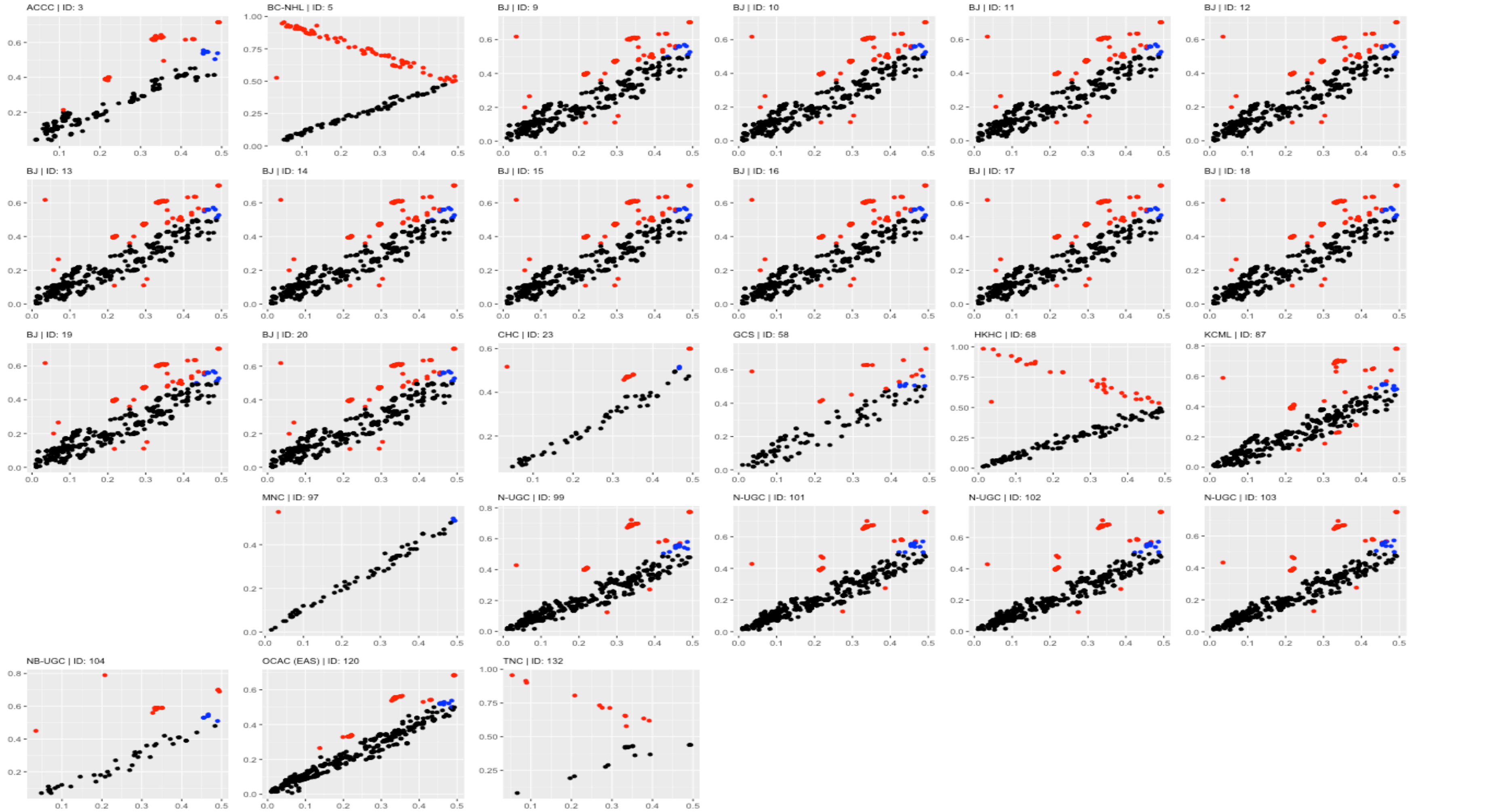


Red data points correspond to SNPs with high allele frequency conflicts, due to having an allele frequency that is greater than 0.58 (when it is expected to be less than 0.5) or to deviation from the reference allele frequency by more than 10 points. Study acronyms are explained in supplementary table 5.

Supplementary Figure 10. Comparison of minor allele frequency between cancer datasets from East Asian studies and super populations from the 1000 genomes project

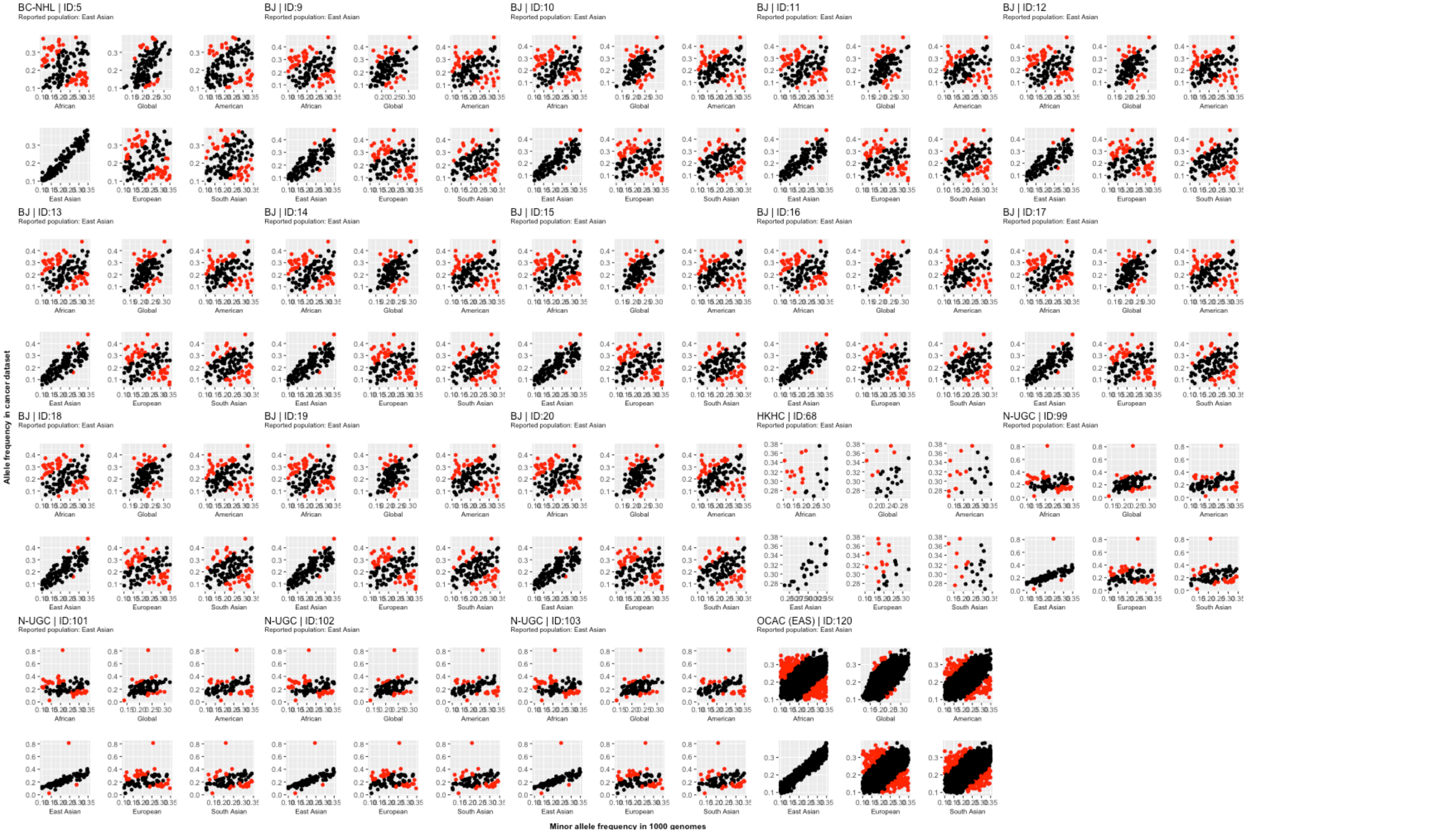


Each data point represents a single SNP. Red data points correspond to SNPs with high allele frequency conflicts, due to having an allele frequency that is greater than 0.58 (when it is expected to be less than 0.5) or to deviation from the reference allele frequency by more than 10 points. Study acronyms are explained in supplementary table 5.

Supplementary Figure 11a. Comparison of minor allele frequency between cancer datasets [ID1 to ID45] from European studies and super populations from the 1000 genomes project

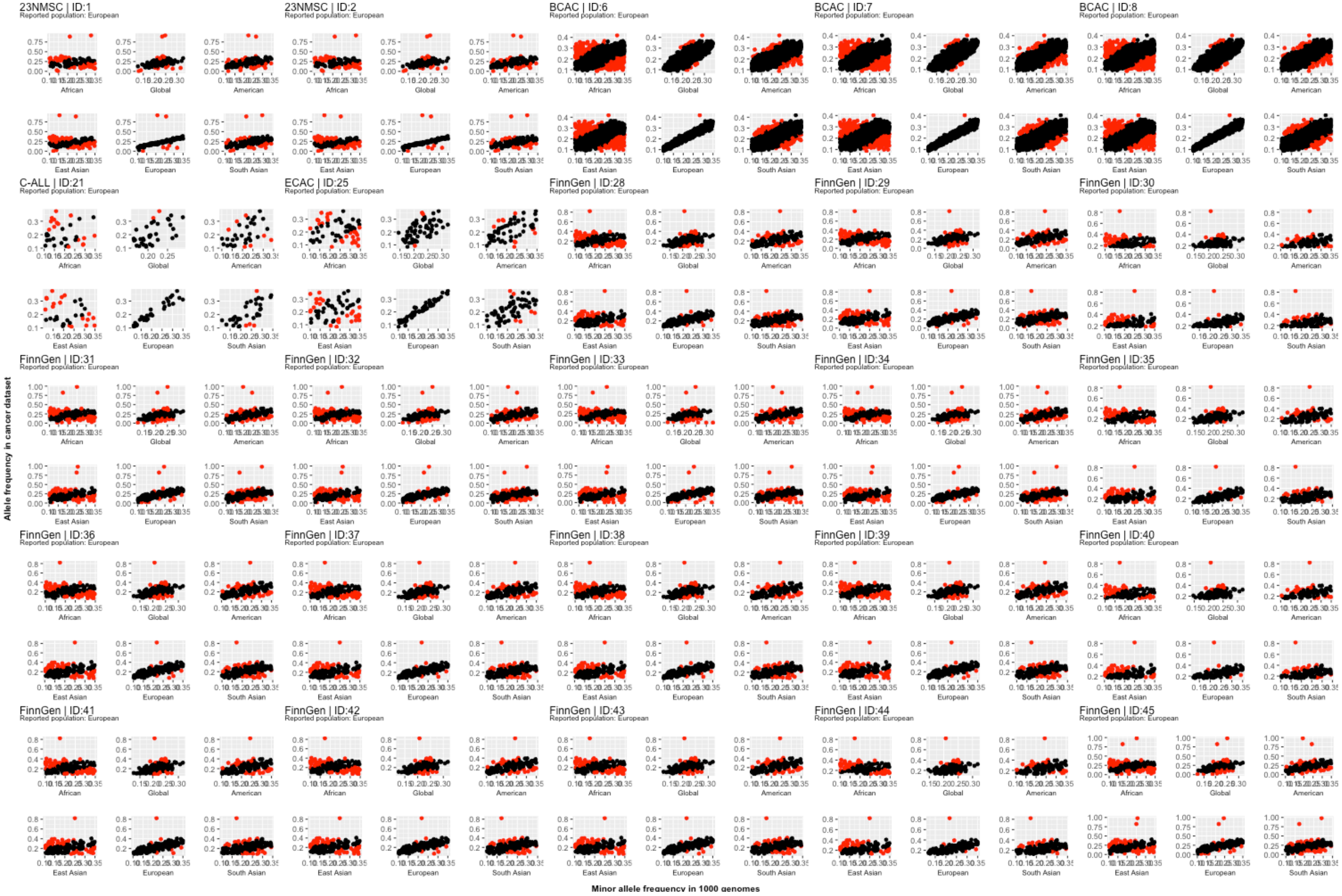


Each data point represents a single SNP. Red data points correspond to SNPs with high allele frequency conflicts, due to having an allele frequency that is greater than 0.58 (when it is expected to be less than 0.5) or to deviation from the reference allele frequency by more than 10 points. Study acronyms are explained in supplementary table 5.

Supplementary Figure 11b. Comparison of minor allele frequency between cancer datasets [ID46 to ID86] from European studies and super populations from the 1000 genomes project

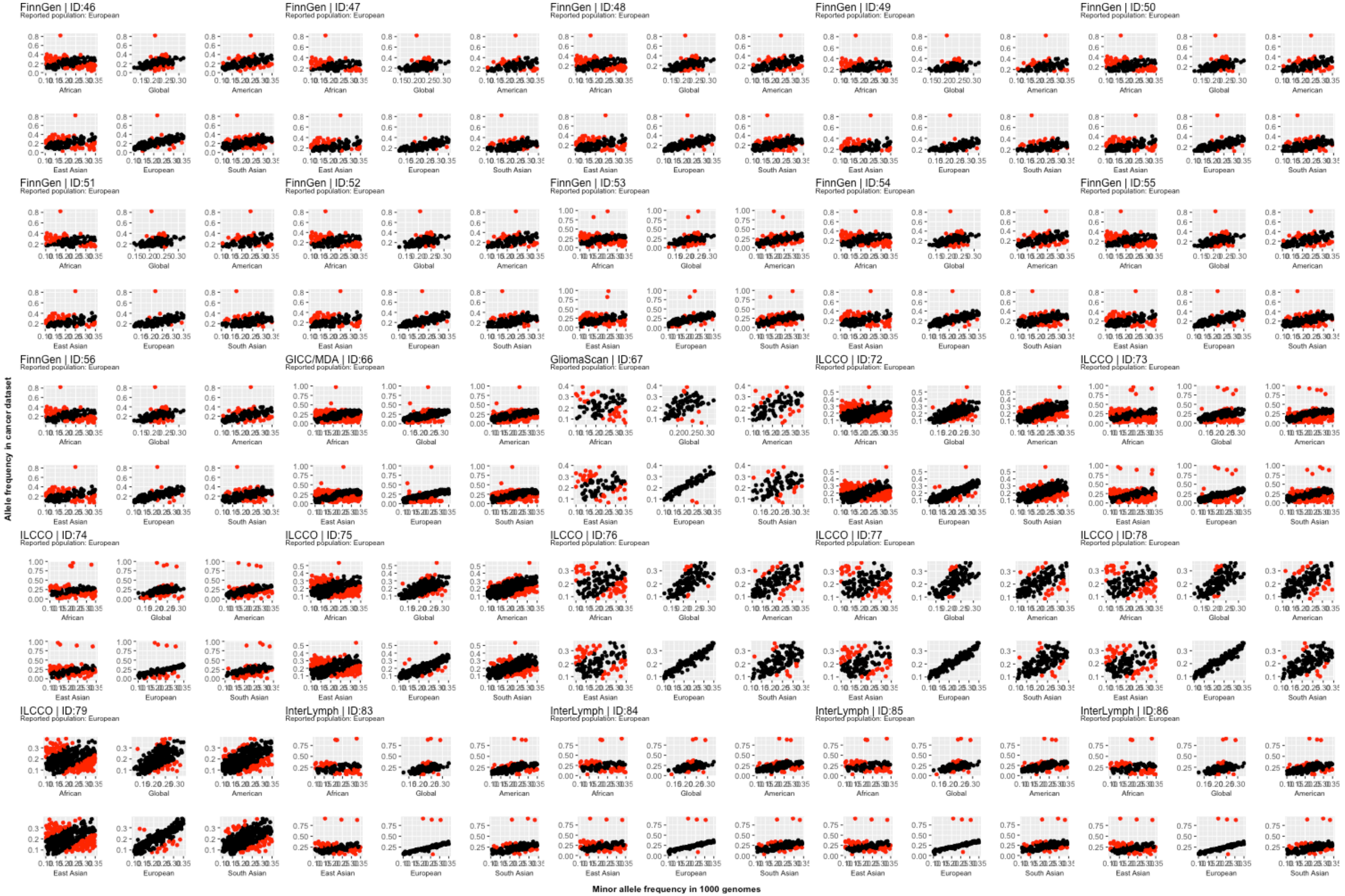


Each data point represents a single SNP. Red data points correspond to SNPs with high allele frequency conflicts, due to having an allele frequency that is greater than 0.58 (when it is expected to be less than 0.5) or to deviation from the reference allele frequency by more than 10 points. Study acronyms are explained in supplementary table 5.

Supplementary Figure 11c. Comparison of minor allele frequency between cancer datasets [ID90 to ID139] from European studies and super populations from the 1000 genomes project

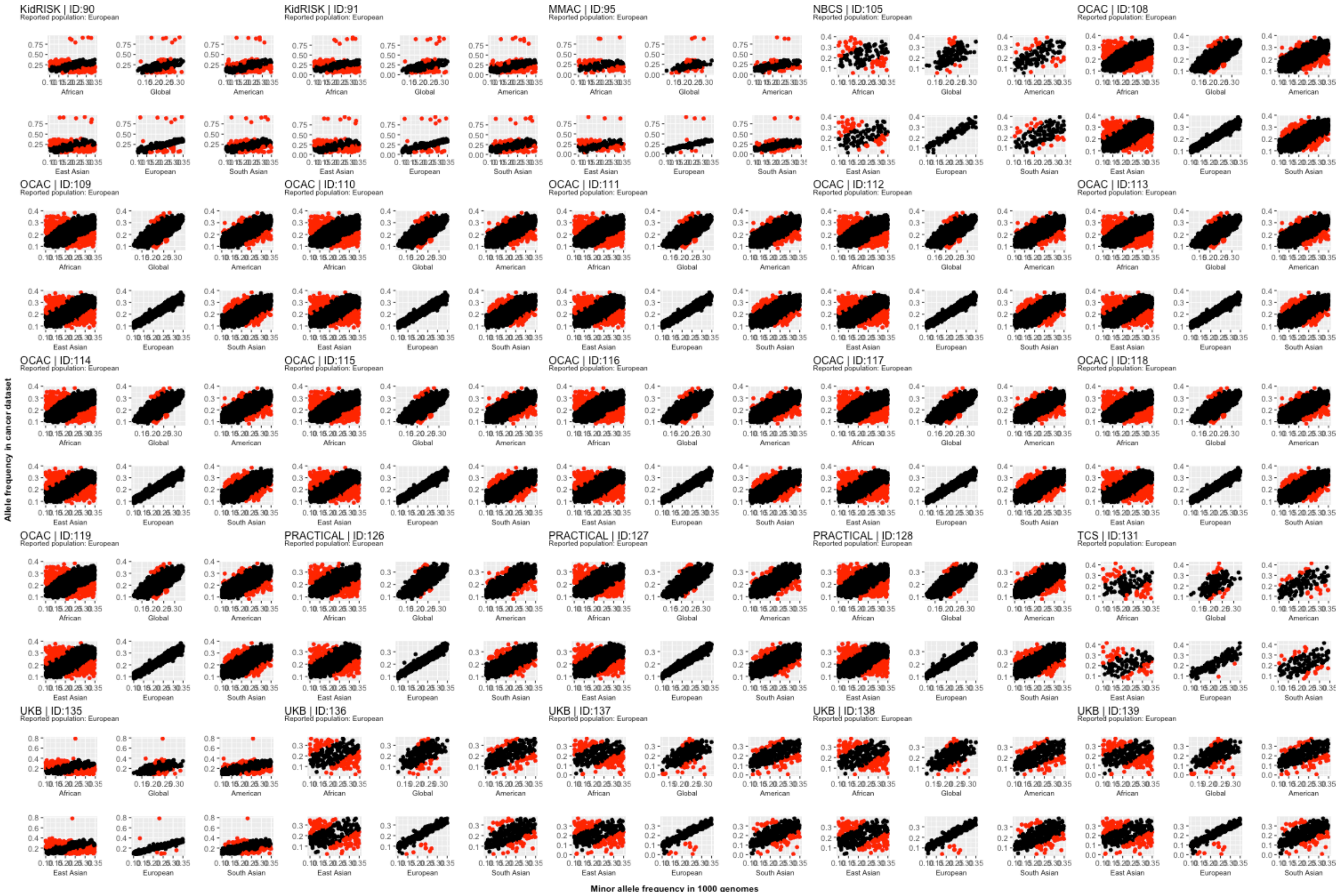


Each data point represents a single SNP. Red data points correspond to SNPs with high allele frequency conflicts, due to having an allele frequency that is greater than 0.58 (when it is expected to be less than 0.5) or to deviation from the reference allele frequency by more than 10 points. Study acronyms are explained in supplementary table 5.

Supplementary Figure 11d. Comparison of minor allele frequency between cancer datasets [ID140 to ID1499] from European studies and super populations from the 1000 genomes project

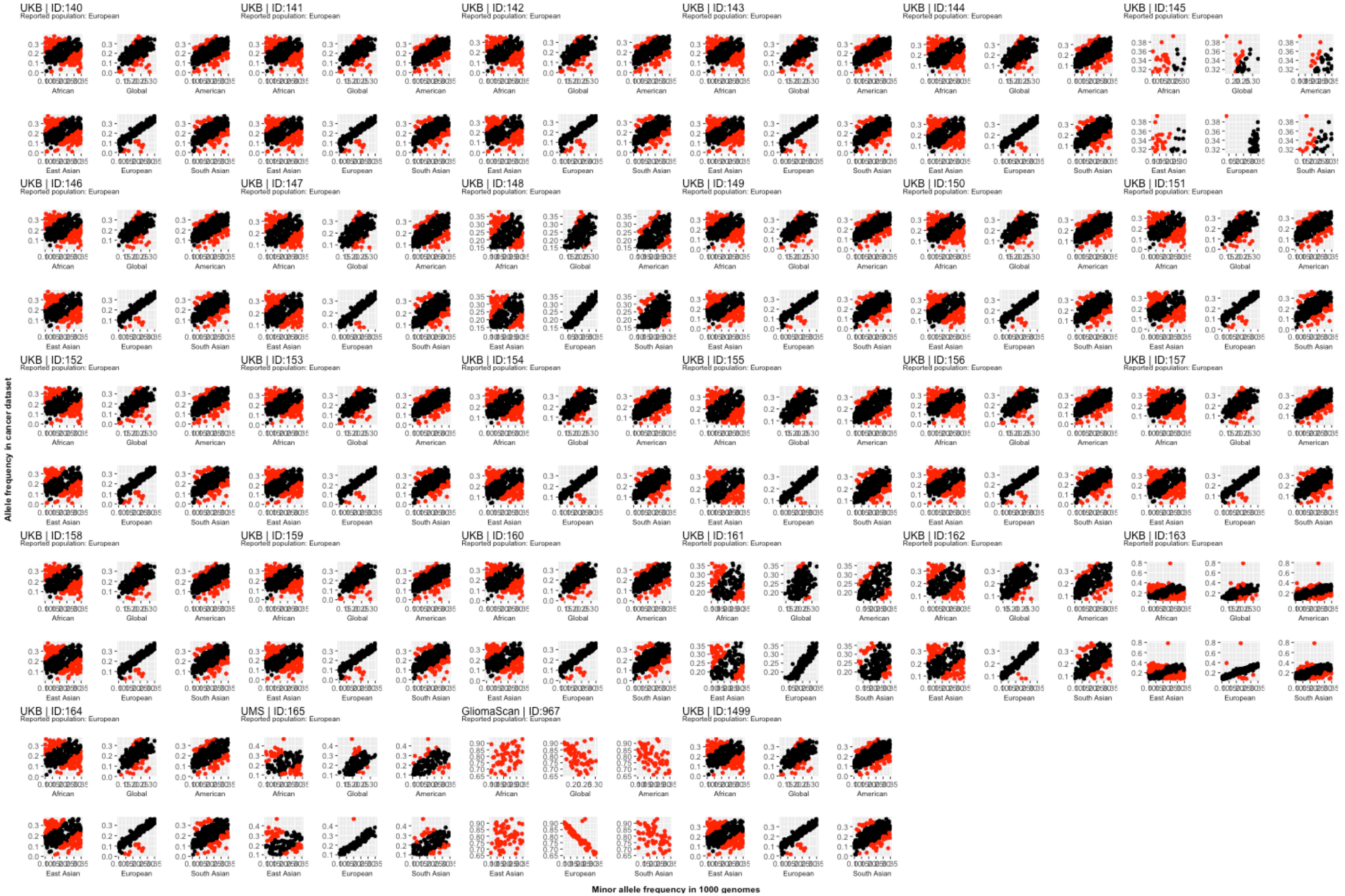


Each data point represents a single SNP. Red data points correspond to SNPs with high allele frequency conflicts, due to having an allele frequency that is greater than 0.58 (when it is expected to be less than 0.5) or to deviation from the reference allele frequency by more than 10 points. Study acronyms are explained in supplementary table 5.

### Supplementary Figure 12. Datasets with discrepancies between the expected log odds ratio and reported effect sizes

The plotted datasets correspond to slopes > 1.2 or <0.8 from models of the expected log odds ratio regressed on the reported effect size.

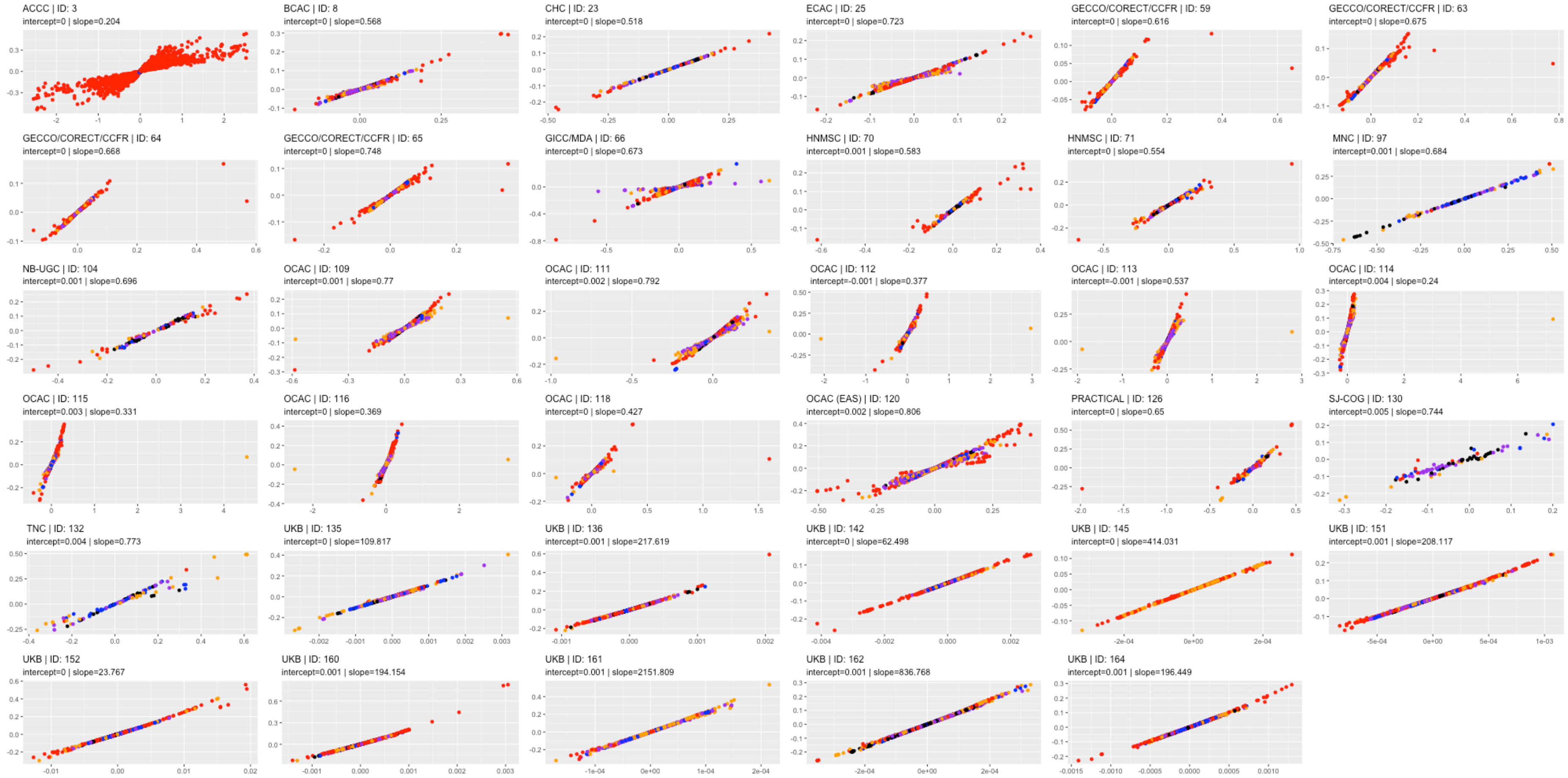

Supplementary figure 13. Comparison of reported and expected log odds ratios in the ACCC dataset

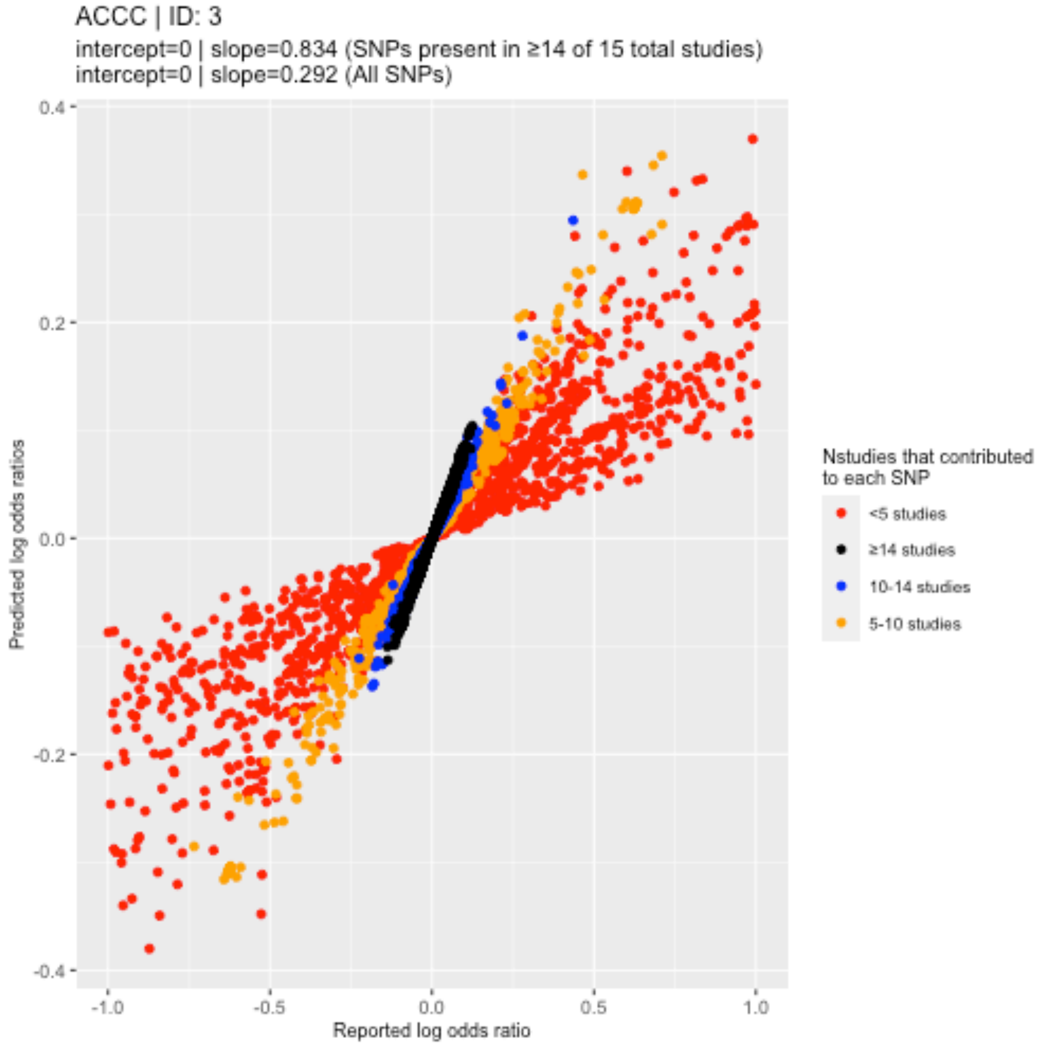


Slopes and intercepts were generated from models regressing the reported log odds ratio on the expected log odds ratio

Supplementary Figure 14. Comparison of reported and expected log odds ratios in the GICC/MDA dataset

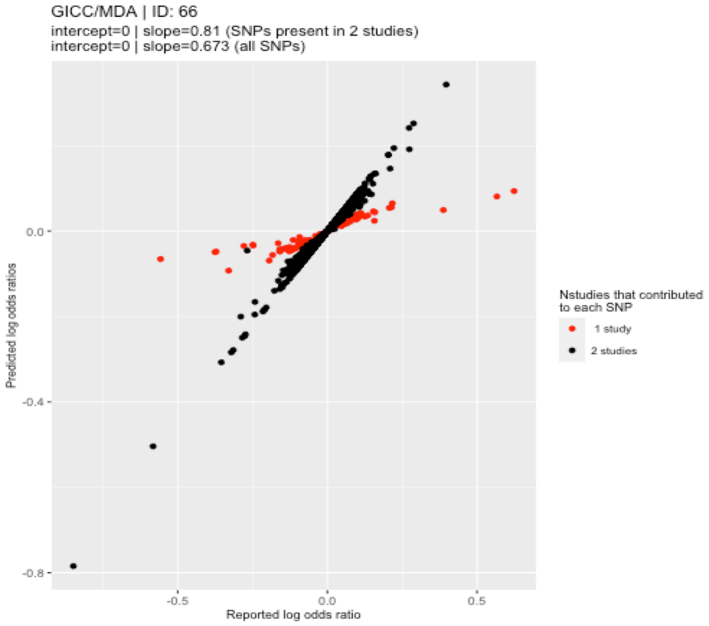


Slopes and intercepts were generated from models regressing the reported log odds ratio on the expected log odds ratio

Supplementary figure 15. Comparison of reported and expected log odds ratios in the GECCO datasets

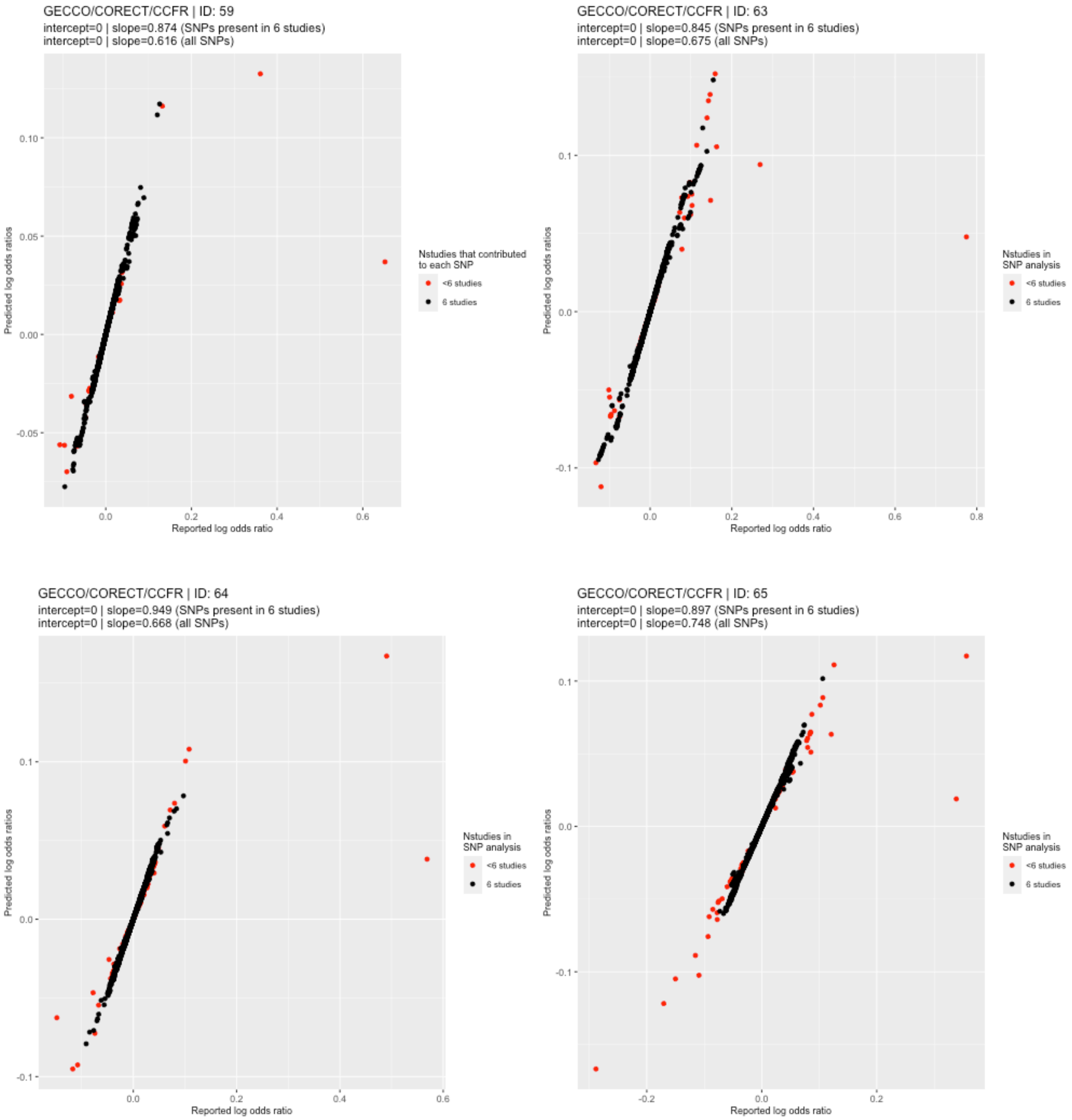


Slopes and intercepts were generated from models regressing the reported log odds ratio on the expected log odds ratio

Supplementary figure 16. Comparison of reported and expected log odds ratios in the HNMSC datasets

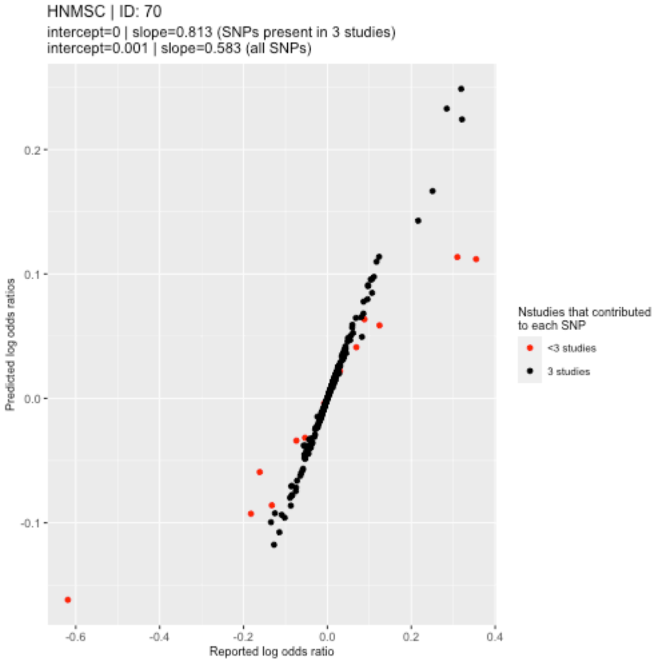


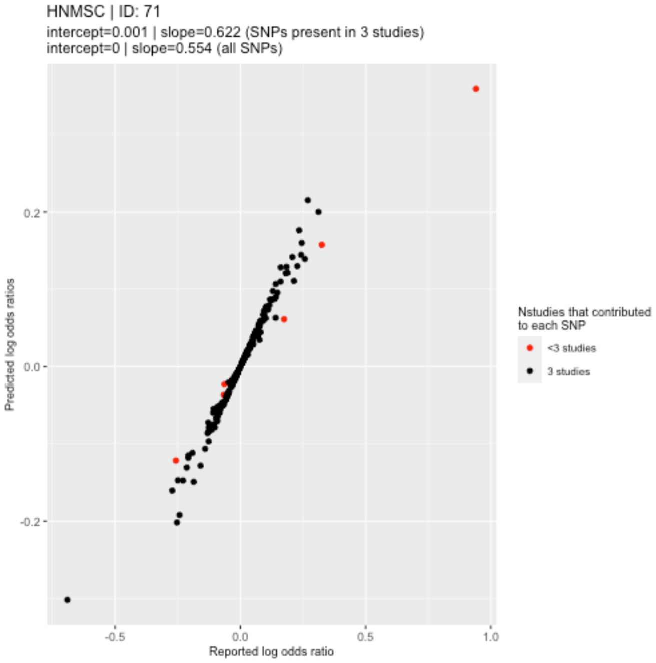

Supplementary figure 17. Deviations in reported from expected log odds ratios attributable to low imputation quality

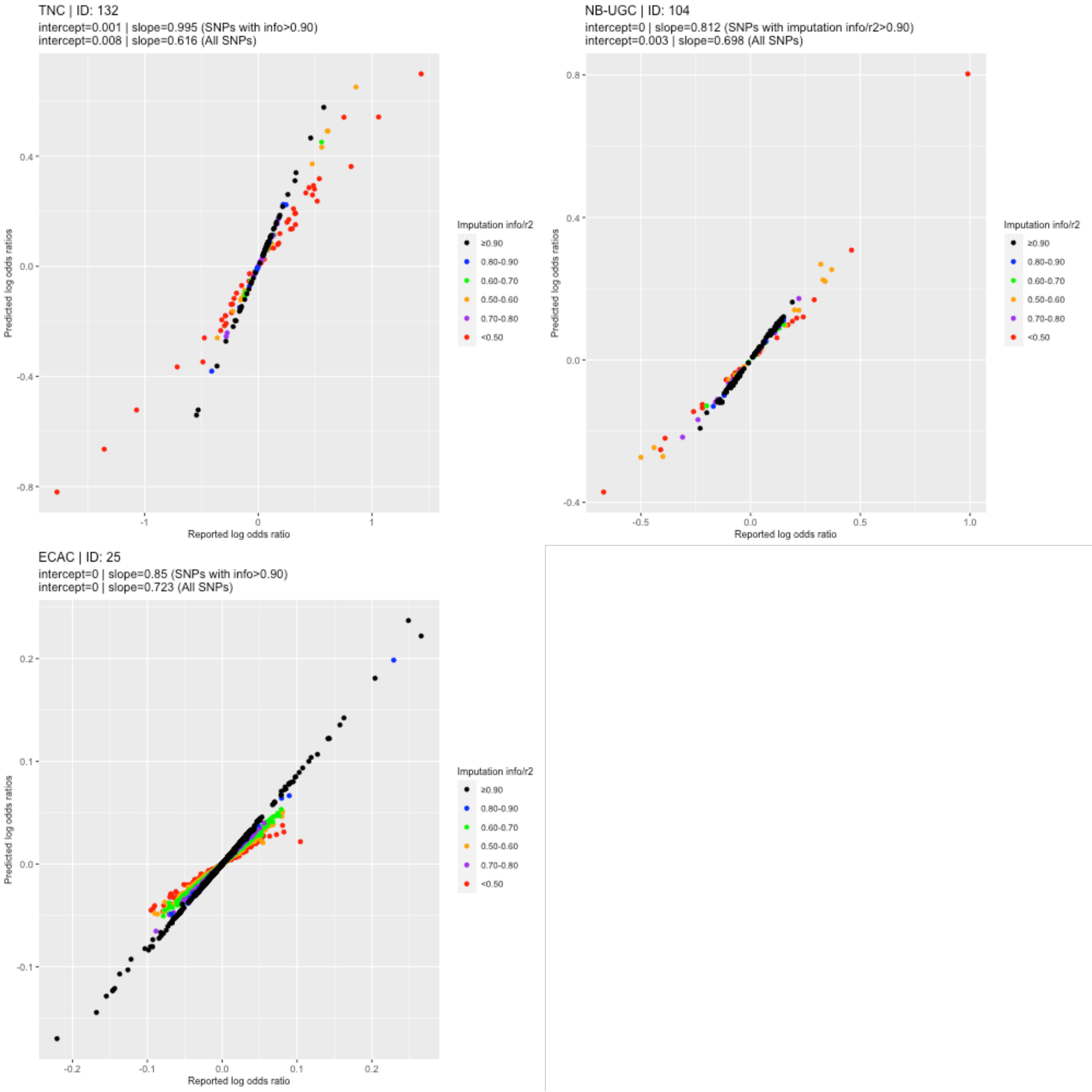

**Supplementary figure 18.** Deviation between reported and expected log odds ratio and imputation quality

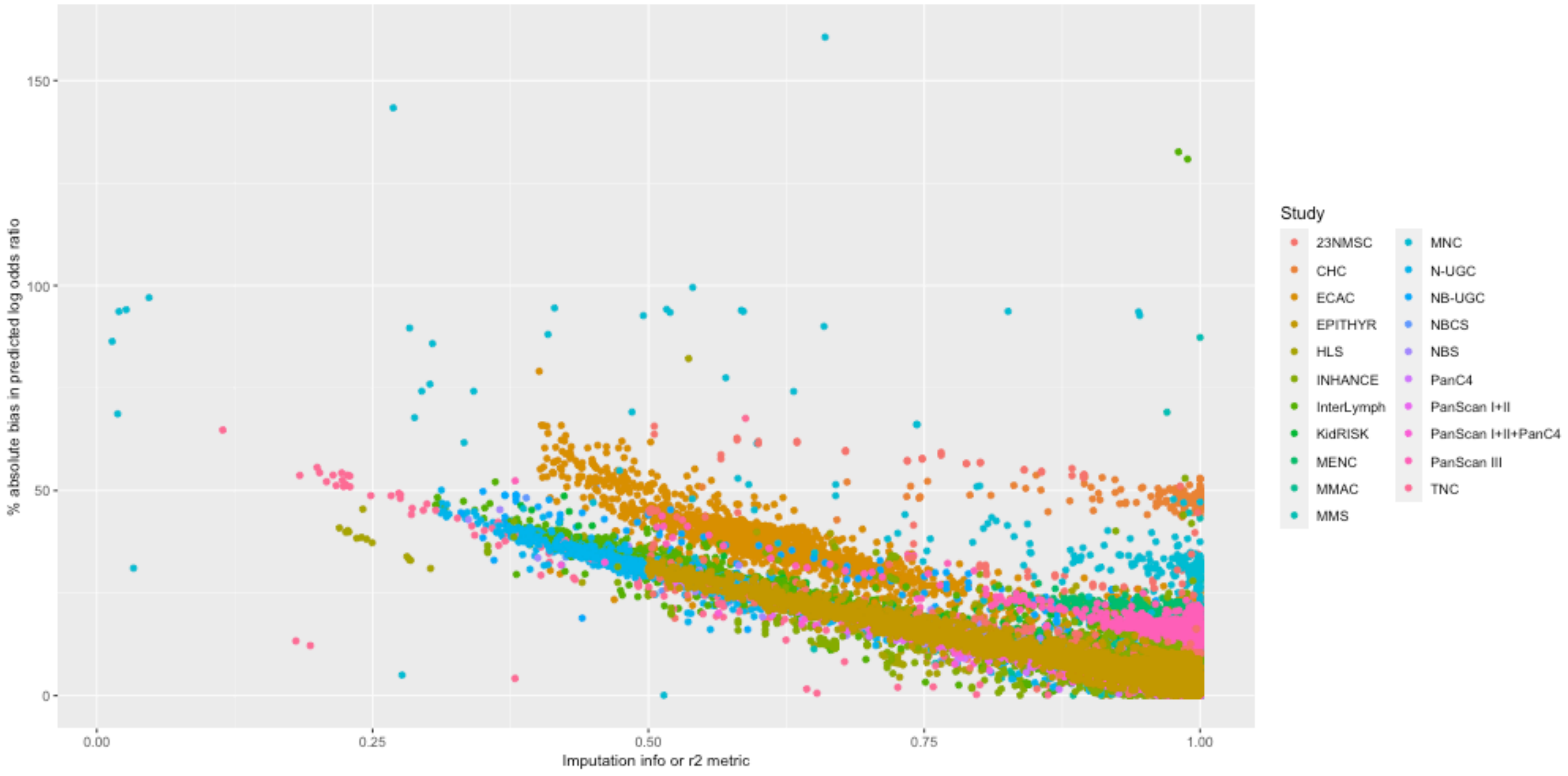

**Supplementary figure 19.** Deviation between reported and expected log odds ratio and P value for deviations from Hardy Weinberg Equilibrium

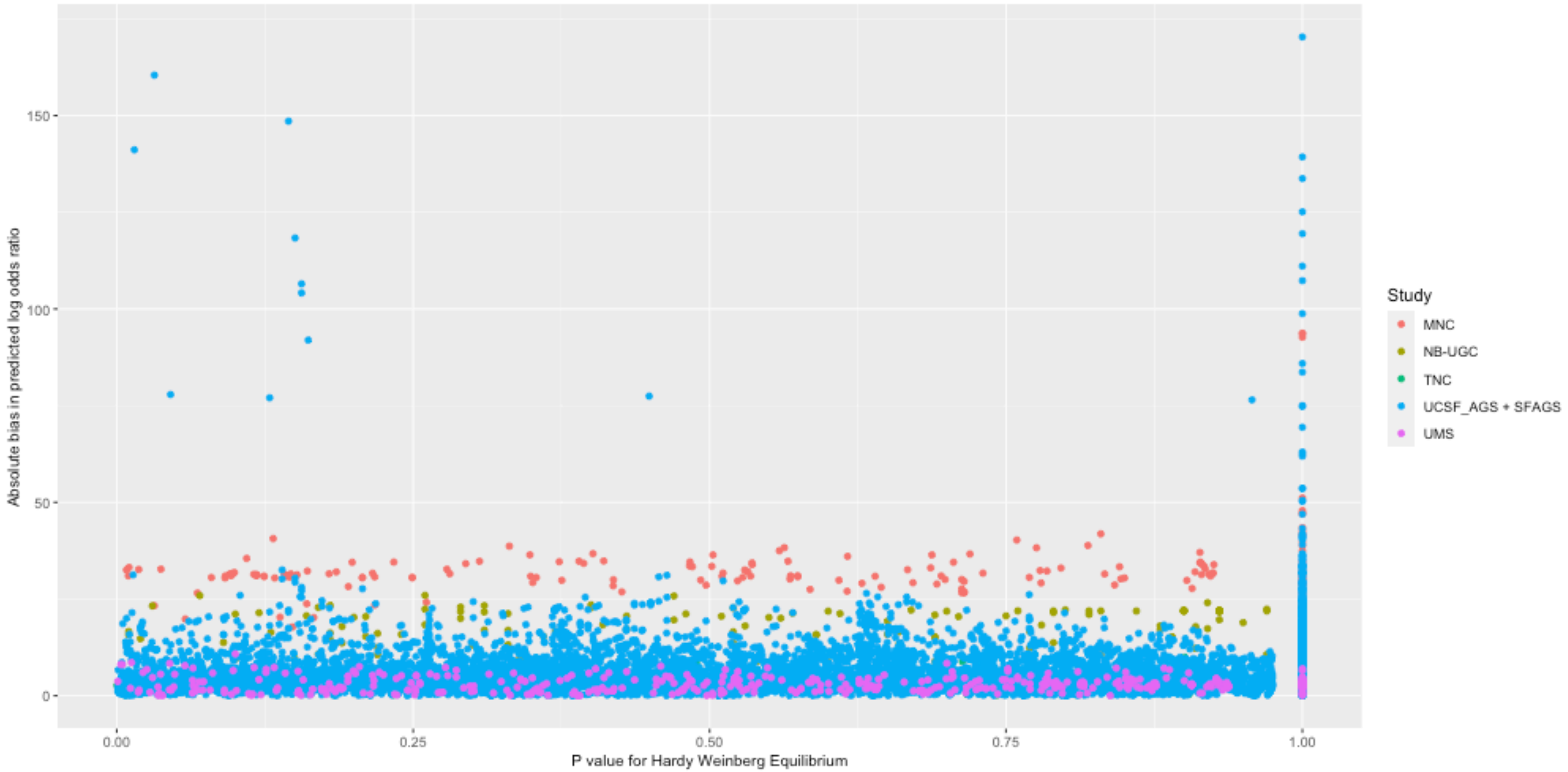

**Supplementary figure 20.** Deviation between reported and expected log odds ratio and P value for between study heterogeneity

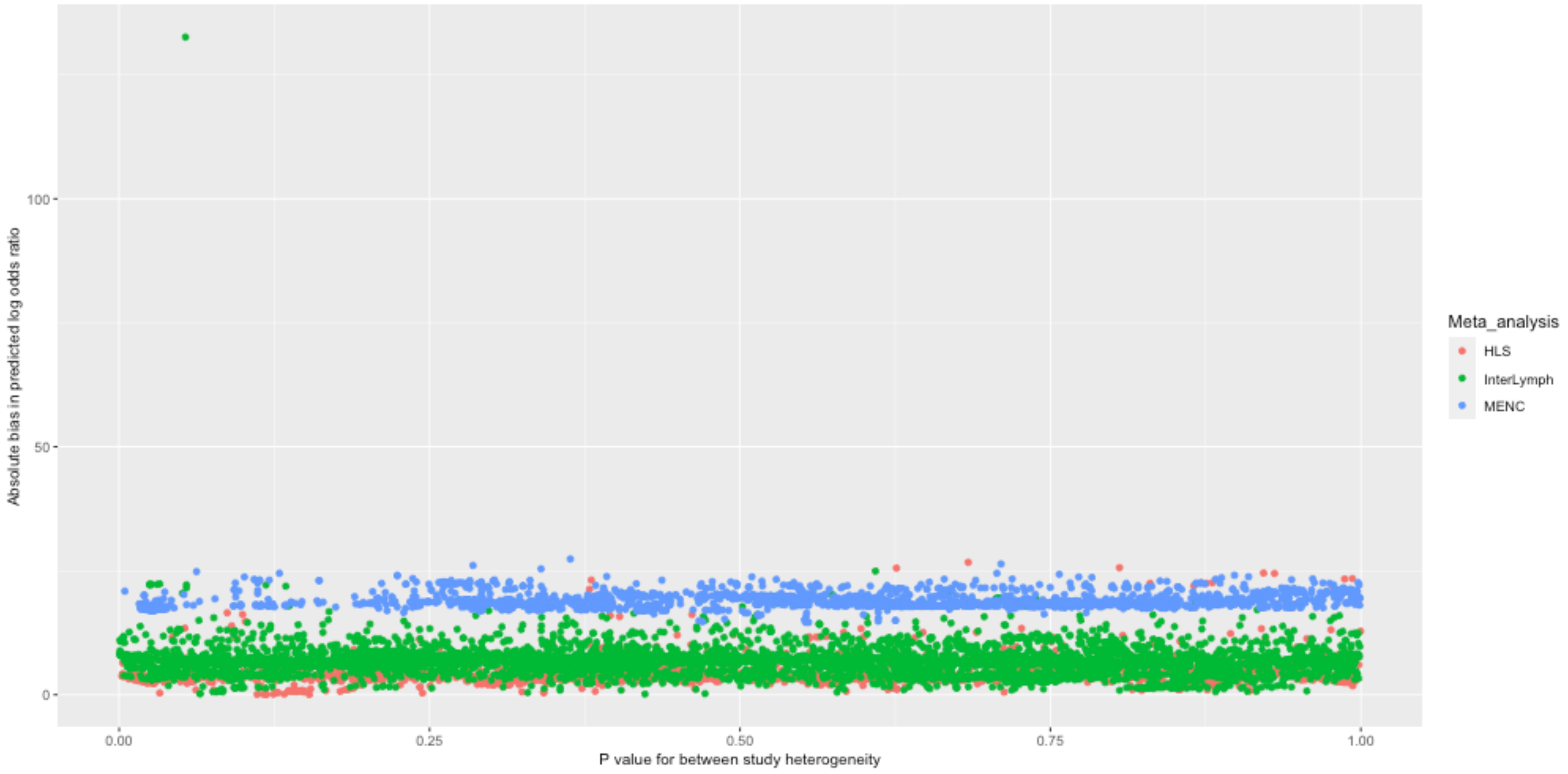

#### Supplementary Figure 21. Correlation between $Z_p$ and $Z_b$ scores within cancer datasets

$Z_p$ = Z scores inferred from reported P values; $Z_b$ = Z scores inferred from reported effect sizes and standard errors. Correlations less than 0.99 are highlighted in red

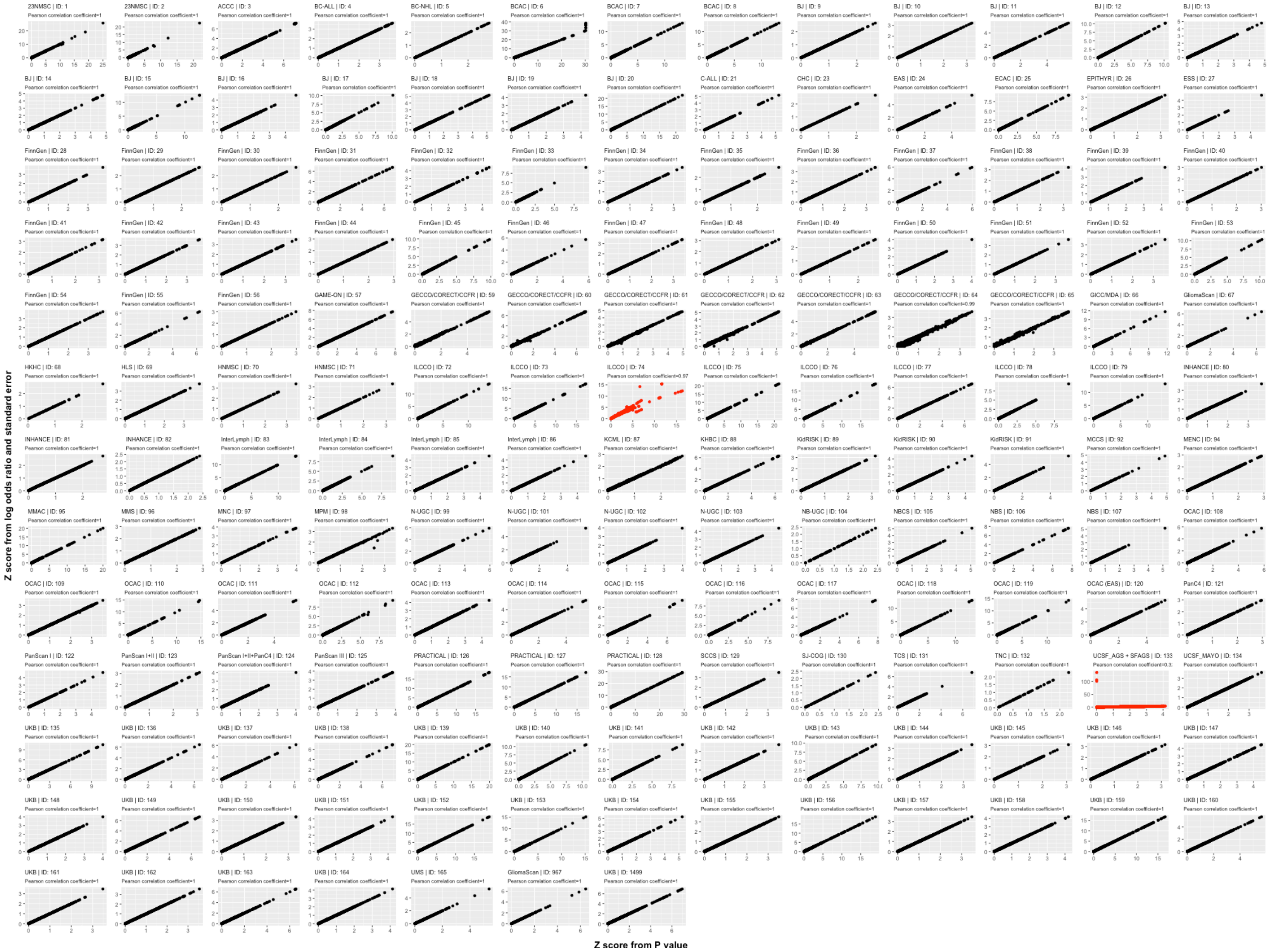
